# Systematic review finds keratoconus AI unproven in fellow eyes with normal topography and tomography

**DOI:** 10.64898/2026.09.29.26364068

**Authors:** Hanyu Jiang, Fei Gao, Ying Jie, Liner Yang, Ying Li, Yang Jiang

## Abstract

Published artificial intelligence for keratoconus is unproven in fellow eyes that topography and tomography call normal, the eyes where surgical decisions are made, although it reports near-perfect accuracy for early disease. In a registered systematic review of 573 studies (PROSPERO CRD420261441197), none of 2,035 claim-by-stratum units from 212 adjudicated studies established discrimination in these eyes. Most units (90.0%) answered a different question, never constructed the target population or certified its normality; external validation did not test label generation, and five systematic reviews did not assess it. In a public cohort, a frozen score yielded an area under the receiver operating characteristic curve of 0.996 against a same-eye index label and 0.549 against a contralateral proxy; its inputs reconstructed the label-generating index (*r* = 0.991), so accuracy there measured agreement with the index. Anchor-gap evaluation measures accuracy in these eyes against the contralateral clinical diagnosis.

## Introduction

In keratoconus screening, the clinically consequential cross-sectional claim is discrimination in very asymmetric ectasia with normal topography and tomography (VAE-NTT); prediction of progression, management benefit and the separation of manifest keratoconus from healthy eyes are distinct tasks, the last of them already established. The VAE-NTT claim has a direct test: the target eye, the eye being classified, must be normal on prespecified topographic and tomographic criteria, while the contralateral eye carries a clinical diagnosis of keratoconus [1]. Positive-class identity comes from the contralateral diagnosis, whereas the prespecified topographic and tomographic criteria establish that the screening rules in clinical use return a negative result in the target eye. The normality requirement matters just as much: the weaker VAE-NT stratum (very asymmetric ectasia, normal topography) certifies the target eye on topography alone and still admits eyes that the tomographic instrument already flags [2].

Both strata occur among fellow eyes of clinically unilateral keratoconus, a population that can appear normal on clinical examination [3,4] and for which clinical decisions include refractive-surgery eligibility [5] and the timing of cross-linking [6,7,8]. The 2015 global consensus holds that true unilateral keratoconus does not exist [7]; by that definition, a fellow eye that passes clinical examination is affected at a stage the instruments have not reported. This is the population that early-detection studies target when they claim to detect disease in eyes that routine instruments do not yet flag [9,1,10]. VAE-NTT makes that cross-sectional claim falsifiable.

In one study design, a model reads the target eye’s tomography and the positive label is calculated from a vendor index derived from those same measurements, a dependence between index test and reference standard that inflates apparent accuracy [11,12,13], so high discrimination can arise from recovery of the index rule rather than from detection of disease. In a second design, the target eye must be normal on prespecified topographic and tomographic criteria and positive-class membership comes from the clinical diagnosis of the contralateral eye, so success requires information beyond what the target eye’s reference measurements already flag.

Target–measurement separation comprises two design features: the source of positive-class identity and the depth of target-eye normality certification relative to the measurements available to the model. The first feature has four levels of separation from the model inputs. At the nearest separation, the target eye’s own device indices determine the label; at the next, the label is derived from the contralateral eye’s device output [14]. Farther out, membership rests on a clinical diagnosis of the contralateral eye, so the label is not computed from the target eye’s measurements. At the greatest separation, positive-class membership is defined by a later clinical outcome [3].

The second feature is the depth of normality certification in the target eye. With topography-only certification, the target stratum can still contain eyes with tomographic signal that the instrument already flags; topography-plus-tomography certification (dual certification) excludes those eyes, leaving only eyes in which both screening rules return a negative result. Together, the two features define the prespecified qualitative design classes used in this review (Fig. 1a). The classes are partially ordered. Among cross-sectional designs, VAE-NTT places both features at their strictest settings, with a clinically anchored positive-class identity and a target eye meeting dual certification.

**Fig. 1.**
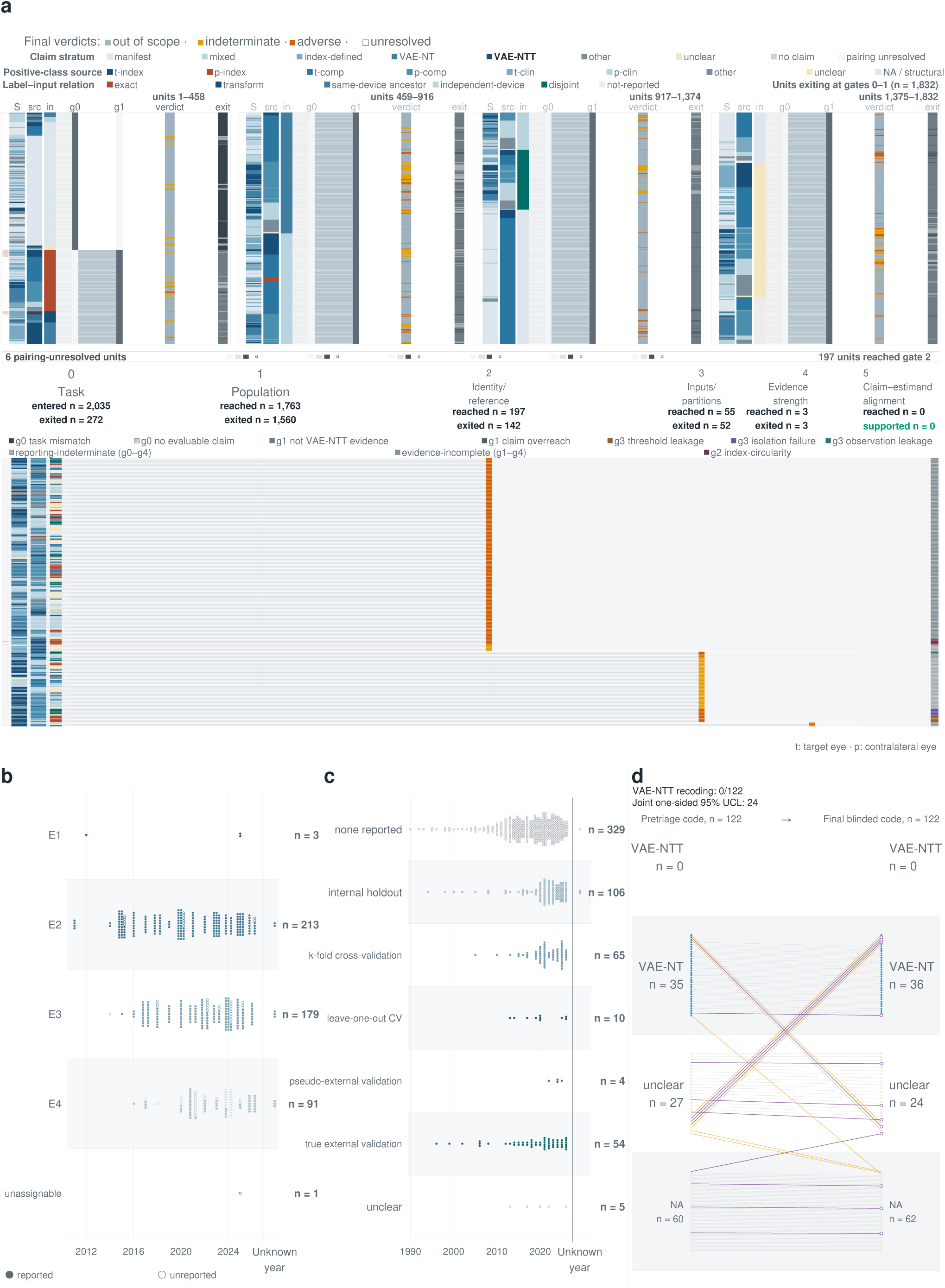
Claim-by-stratum audit of 2,035 claim units through six gates. **a**, Display rows show 2,035 claim units from 212 adjudicated studies, annotated by claim stratum (S), positive-class source (src), label–input relation (in), gate-0/1 exit code (g0, g1, exit) and final verdict. Claim strata include VAE-NTT (very asymmetric ectasia with normal topography and tomography; 210 units) and the weaker VAE-NT (normal topography only; 310), alongside manifest (642), mixed (395), index-defined (359), other (108), unclear (4), no-claim (1) and pairing-unresolved (6) units. Positive-class sources combine the eye of origin (t, target; p, contralateral) with the identity type (index, composite or clinical); NA marks units with no positive-class source. The 1,832 units (90.0%) exiting at gates 0–1 form four equal display blocks (units 1–458, 459–916, 917–1,374 and 1,375–1,832); the 197 units that reached gate 2 and the 6 pairing-unresolved units are shown below. The lower track follows advanced units through gates 2–5 (identity/reference; inputs/partitions; evidence strength; claim–estimand alignment): 197 reached gate 2, 55 reached gate 3, 3 reached gate 4, none reached gate 5 and none was supported. Final verdicts were 1,540 out of scope, 234 indeterminate, 255 adverse and 6 unresolved (exit reasons by gate, Supplementary Table S3). Red marks flag positive-class index circularity, assigned only when both conditions hold jointly: positive-class grouping is defined by an index, and the label-generating index is retained in the model inputs exactly or through a deterministic transformation (24 of 2,035 units). **b**, Normality-certification evidence class by publication year for the 487 normal-eye-stratum units: E1, per-eye certified thresholds; E2, paper-stated numeric criteria or joint category counts; E3, aggregate distributions only; E4, narrative description only. Open circles mark the 75 units (15.4%) without an operational normality criterion. **c**, Validation category by publication year for all 573 studies. **d**, Blinded target-stratum recoding of 122 studies, pretriage code versus final blinded code; outlined endpoints mark the 12 third-rater adjudications. No study was recoded to VAE-NTT, and the joint one-sided 95% upper confidence limit (UCL) for VAE-NTT studies among those still unchecked is 24.

The same index can enter a study in two roles. On the exclusion side, it records that the deployed rule returned a negative result in the target eye and thereby defines the question; on the positive side, it can define the label the model is rewarded for recovering and thereby answer the question in advance. Reference-pathway leakage occurs when an index defines the positive class and the model inputs retain the measurement pathway needed to reconstruct that index. Unlike train–test leakage, it requires no case overlap across data partitions, because the overlap lies between the label-generating rule and the input measurements, and the same pathway persists when a public dataset transmits the inherited label to downstream studies. At the design level, the label-generating reference is functionally incorporated into the inputs, and that overlap persists across sites, cohorts and operators, so external validation can move a model to a new site without breaking the pathway.

Existing appraisal frameworks can flag this risk but do not connect the flag to the reported accuracy. The Quality Assessment of Diagnostic Accuracy Studies 2 (QUADAS-2) and Prediction model Risk Of Bias ASsessment Tool (PROBAST) instruments assess the reference standard [15,16]; the Standards for Reporting of Diagnostic Accuracy Studies (STARD 2015) ask whether index-test results were available to those who read the reference standard [17], the Transparent Reporting of a multivariable prediction model for Individual Prognosis Or Diagnosis + Artificial Intelligence (TRIPOD+AI) statement asks for any blinding of outcome assessment [18], and the Checklist for Artificial Intelligence in Medical Imaging (CLAIM 2024) asks how reference-standard annotations were obtained [19]. None quantifies how much measured performance survives as the target moves away from the measurement pathway, or whether performance measured in a broader stratum can support a claim in a stricter one. A study can therefore be externally validated and still leave the VAE-NTT target untested.

We assessed whether published accuracy estimates establish discrimination in VAE-NTT and, where they do not, what sustains high reported accuracy. In a 573-study corpus, we adjudicated claim-by-stratum units from 212 studies against six ordered gates, and the remaining 361 formed a target-stratum verification pool assessed by stratified blinded sampling. We traced the provenance of the composite indices that anchor the field’s reference labels, characterized how accuracy and its uncertainty were reported, and used partial identification to bound strict-stratum performance from published broad-stratum data. We then evaluated a frozen score as its target moved away from the label-generating pathway in a public cohort and assessed age- and sex-only discrimination in an independent cohort. Together, the analyses show that published keratoconus AI has not established the capability for which it is cited and that the anchor gap, the gap between the clinical target named in a claim and the evidence supplied for that target, runs through the whole evidence chain, from target construction and label generation to reference indices, public datasets and systematic reviews. They also yield anchor-gap evaluation, a proposed reporting standard that links each screening claim to the evidence for its target.

## Results

### VAE-NTT discrimination was not established in the corpus

None of the 2,035 claim-by-stratum units from 212 adjudicated studies met the gate criteria for supported discrimination in VAE-NTT (Fig. 1a). Each unit paired one performance claim with one target stratum. The 573-study corpus (identification and eligibility accounting in Supplementary Table S1; study characteristics in Supplementary Table S2) comprised these 212 studies with unit-level adjudication and 361 studies in the target-stratum verification pool; gate-specific results for all 2,035 units are given in Supplementary Table S3. No study in the 122-study stratified verification sample, drawn from the non-adjudicated pool as it stood at sampling, was reclassified as VAE-NTT (Fig. 1d). The one-sided joint 95% upper confidence limit for the number of VAE-NTT studies still unchecked in that sampling frame was 24 (Supplementary Table S4).

Gates 0 and 1, which asked only whether a relevant claim existed and whether the target population was constructed with documented normality, removed 1,832 of the 2,035 units (90.0%): these units answered a different question, never constructed the target population, or never certified normality. Of the remaining 203 units, 6 had unresolved claim–stratum pairings and 197 advanced to gates 2–5, and none of those 197 passed all four gates. Among 487 units invoking a normal-eye stratum, 75 (15.4%) reported no operational normality criterion (Fig. 1b), so in these units “normal” functioned as a group label with no documented measurement behind it.

### Validation was abundant but addressed other targets

All 92 deep-learning studies reported at least one validation procedure, and 212 studies evaluated fixed, named indices against established diagnoses (Fig. 1c; Supplementary Table S2). The 212 fixed-index studies formed a separate set from the 212 adjudicated studies. Only 29 of 573 studies (5.1%) met all three adverse conditions of fitting parameters, claiming an early or subclinical target and reporting no validation of any kind. The adverse classification applied to early and subclinical claims; manifest-disease tasks formed a separate target class. Within subclinical claims, 11 studies used a purely index-defined positive class and 43 used mixed index–clinical definitions. Nine of the 11 index-only studies also retained a shared measurement pathway and therefore met the study-level definition of reference-pathway leakage, whereas the other two had index-defined labels without measurement-pathway overlap. Median cohort size across these 11 studies was 122 eyes, and the largest (4,894 eyes) was a preprint that had not undergone peer review.

Among 54 studies with true external validation, 17 were extracted as constructing VAE-NTT or a stricter target, and 6 of those 17 were extracted as sourcing positive-class identity from a clinical diagnosis outside the index. On cell-level review of those six, three constructed the stratum in full and three conditionally; for positive-class identity, three met the criterion in full, two met it conditionally, and one classified target eyes within the measurement pathway (Table 1). Pathway orthogonality, defined here as a detector outside the exclusion/label measurement family, was demonstrated in none of the six: five shared the pathway and one did not report it (Table 1). These nested counts split external validation into four distinct dimensions: transport to data outside development, construction of the intended target, independence of positive-class identity from the index, and orthogonality of the measurement pathway. Steinberg et al. formally stratified eyes into VAE-E (the ectatic eye), VAE-NT and VAE-NTT, yet 17 of the study’s 19 units (89.5%) exited at gate 1 when the audit required per-eye certification [1]. Almeida et al. labeled their groups VAE-NTT, but per-eye normality on both topographic and tomographic criteria could not be verified from the report [20]. Existing studies supplied the individual components of a clinically anchored design, but no supported result combined them (Table 1).

**Table 1.** Feasibility map. Strict target stratum, clinically sourced positive-class identity, and per-eye exclusion rulers use the Table 3 states (Met, Conditional, Not demonstrated, Not reported) followed by the deciding evidence; detector–reference pathway uses Orthogonal, Shared, and Not reported. Abbreviations: BAD-D, Belin/Ambrósio Enhanced Ectasia Display deviation index; FF, forme fruste; FFKC, forme fruste keratoconus; I-S, inferior–superior asymmetry value; IHD, index of height decentration; KCI, keratoconus index; KISA%, keratoconus percentage index; Kmax, maximum keratometry; KSI, keratoconus severity index; LASIK, laser-assisted in situ keratomileusis; SS, combined forme fruste and suspect keratoconus group; UZG, Ghent University Hospital; VAE-NT, very asymmetric ectasia with normal topography; VAE-NTT, very asymmetric ectasia with normal topography and tomography. Herber et al., 2022 is shown as the single-site architecture example and is excluded from the six-candidate count.

| Study | True external transport | Strict target stratum | Clinically sourced positive-class identity | Per-eye exclusion rulers | Detector–reference pathway | What the design establishes — and what it does not |
| --- | --- | --- | --- | --- | --- | --- |
| Karimi et al., 2018 [45] (FT126) | Yes — independent test data reported | Conditional — a suspect stratum is named but defined by persistent diagnostic uncertainty rather than by stated eligibility criteria | Conditional — three ophthalmologists assigned identity, but Corvis and Pentacam also entered the identification pathway | Not reported — a complete ophthalmic examination is described, but no named exclusion ruler or per-eye result | Shared | Demonstrates Corvis-based modeling across healthy, suspect, and keratoconic eyes. The models use the same mechanical-property values across healthy and keratoconic eyes. |
| Issarti et al., 2020 [46] (FT512) | Yes — UZG external-center data excluded from training | Met — five strata are explicit; the suspect target eye is asymptomatic with no clinical or tomographic signs while the contralateral eye has clinical keratoconus | Met — partner-eye clinical keratoconus anchors the suspect stratum | Not demonstrated — target-eye normality is narrative; no named numeric exclusion ruler in the suspect definition | Shared | Adds suspect-keratoconus detection and an objective severity score to five-stratum classification. Healthy and suspect cases still overlap modestly. |
| Wahba et al., 2018 [47] (FT666) | Yes — a separate prospective validation sample was collected | Met — FFKC explicitly required no clinical, curvature, elevation, or pachymetry abnormality plus fellow-eye keratoconus | Met — evident keratoconus in the fellow eye while the target eye is normal across the stated domains | Conditional — clinical, curvature, elevation, and pachymetry domains were applied on the exclusion side, but numeric cut-offs were not reported | Shared | Establishes strong I-S–IHD correlation. It does not establish strict-target diagnostic performance, and right-eye selection differed for scarce FFKC cases. |
| Saad & Gatinel, 2012 [48] (FT686) | Yes — validation eyes were consecutive cases excluded from score creation | Conditional — normal versus at-risk populations were predefined, but the at-risk stratum was assigned by one surgeon's subjective reading of the maps | Not demonstrated — target-eye classification from Placido and elevation maps within the measurement pathway, without an outside-pathway clinical or partner-eye anchor | Not demonstrated — target eyes were subjectively classified from maps rather than certified by reported per-eye rulers | Shared | Shows reasonable concordance between objective and subjective classifications for early subclinical keratoconus detection. The score remains tied to Orbscan topographers. |
| Ruiz Hidalgo et al., 2017 [49] (FT687) | Yes — KA (Keratoconus Assistant) was applied to a cohort not used in development | Conditional — clinical groups are explicit, but the SS group combines forme fruste and suspect eyes under center-specific workflows | Conditional — the forme fruste component has a partner-eye anchor, but the combined SS group also contains suspect eyes | Not reported — center standards are named, but the full ruler set and per-eye measurements are not reported for the combined FF/SS target | Not reported | Supports cross-sectional identification of forme fruste keratoconus framed as at risk of ectasia after refractive surgery. It does not establish prediction of post-LASIK ectasia; equipment and forme fruste definitions differed between centers. |
| Chan et al., 2015 [50] (FT688) | Yes — masked independent testing in a new all-Asian cohort | Met — target-eye normality defined by clinical and topographic examination plus KCI/KSI, keratometry, posterior-elevation, and axial-map criteria, with frank keratoconus in the contralateral eye | Met — contralateral keratoconus reconfirmed by a corneal subspecialist through clinical examination and topography | Met — KCI/KSI plus curvature, elevation, and axial-map criteria were applied to target-eye normality | Shared | Supports cross-sectional identification of FFKC framed as at risk of post-LASIK keratectasia. It does not establish prediction of post-LASIK ectasia or detect every FFKC case. |
| Herber et al., 2022 [33] (FT378) — architecture example, not one of the six true-external candidates | No — fixed indices evaluated in one prospective cohort; no development-to-validation transport | Met — VAE-NT and VAE-NTT separated explicitly with named topographic and tomographic thresholds | Met — partner eye had clinical keratoconus | Met — KISA%, I-S, Kmax, and BAD-D thresholds applied to the target eye | Orthogonal | Demonstrates tomographic exclusion with an orthogonal biomechanical detector. Generalizability is limited by the White study population and small keratoconus sample. |

Large studies showed the same anchor gap. A JAMA Ophthalmology study of 2,802 eyes contributed nine claim units, six of which exited at gate 1, and none was ultimately supported [9]. An American Journal of Ophthalmology study of 1,035 eyes performed true external validation [21], yet 13 of its 15 units exited at gate 1, which required the target population to be constructed with documented normality.

### Index use was common, and identifiable derivations addressed other targets

Qualifying use of an index was common; use as ground truth was rare. In 217 of 573 studies (37.9%), at least one of seven composite indices was either evaluated or used as a reference standard, label source or eligibility criterion, and 231 studies mentioned at least one index (Fig. 2a). Six met the prespecified usage threshold to count as load-bearing, and the seventh, the second version of the Tomographic and Biomechanical Index (TBI; TBIv2), fell below it. BAD-D, the composite deviation index of the Belin/Ambrósio Enhanced Ectasia Display, was mentioned in 150 studies and had a qualifying use in 140 (Fig. 2a). Of the 140 studies with a qualifying BAD-D use, 27 (19.3%) used it as a reference standard, label source or eligibility criterion, and 7 of those 27 (5.0% of 140) used it to anchor ground truth (index-provenance audit in the Zenodo archive). Qualifying use therefore exceeded use as ground truth 20-fold.

**Fig. 2.**
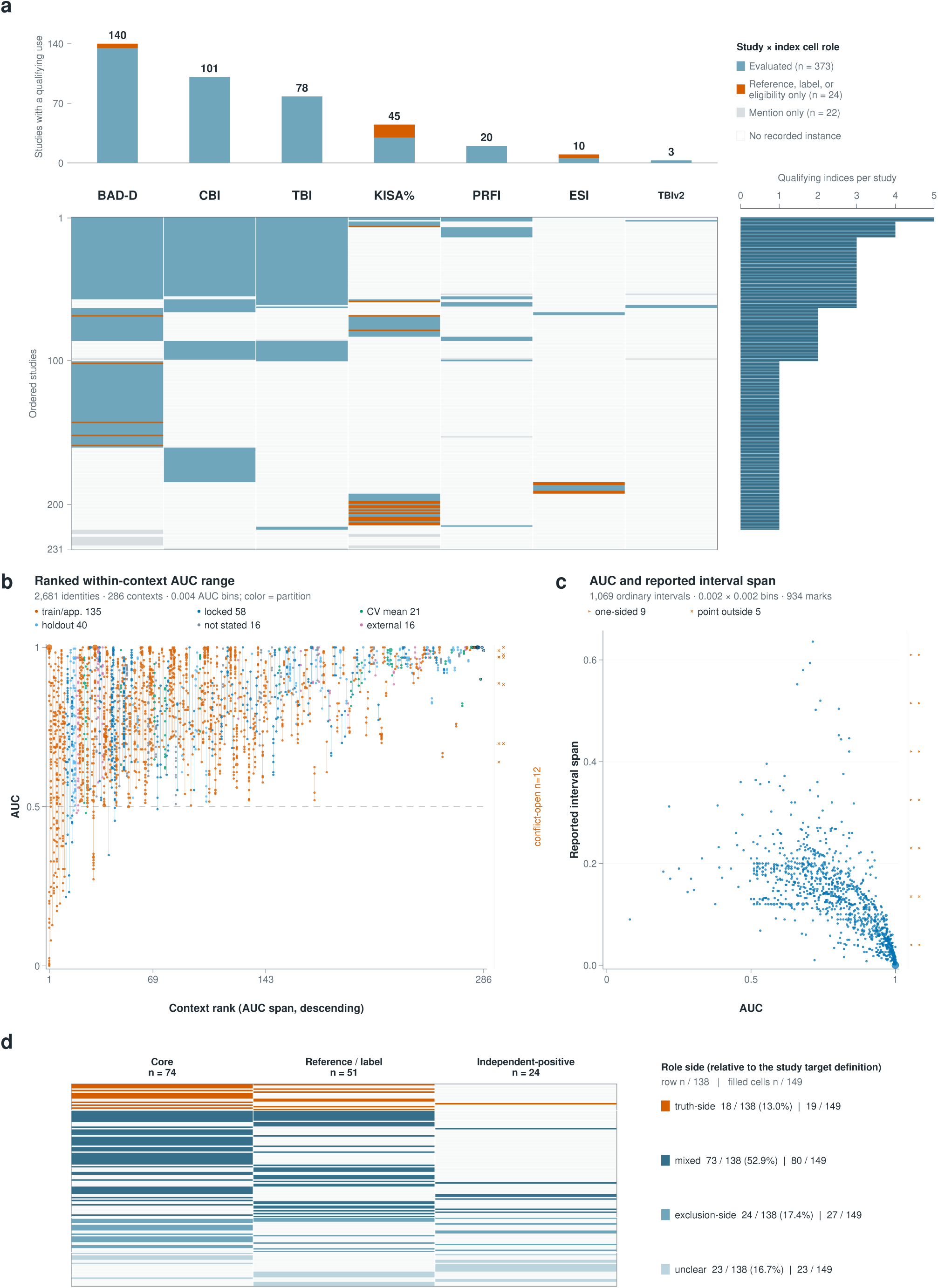
Composite-index usage roles, within-context AUC dispersion, reported-interval structure and membership pools. **a**, Roles of seven composite indices (419 study × index cells): 231 studies (rows) by index (columns; qualifying-use study counts: BAD-D (Belin/ Ambrósio Enhanced Ectasia Display deviation index) 140, CBI (Corvis Biomechanical Index) 101, TBI (Tomographic and Biomechanical Index) 78, KISA% (keratoconus percentage index) 45, PRFI (Pentacam Random Forest Index) 20, ESI (Ectasia Screening Index) 10, TBIv2 3). Cell roles are evaluated (*n* = 373); reference, label or eligibility only (*n* = 24); mention only (*n* = 22); or no recorded instance. Rows include 14 mention-only studies; cells with both roles are plotted as evaluated; the right margin counts qualifying indices per study. **b**, AUC within fixed contexts (a context fixes study, target, contrast, dataset, partition and endpoint). The 286 contexts comparing distinct models or indices are ordered by AUC span; vertical segments span each context’s AUC range; identities are binned at 0.004 AUC, marker area encoding the number per bin, and color marks the partition (CV, cross-validation; train/app., training or apparent-performance partition; locked, pre-existing locked partition). Median within-context span is 0.158; 69 contexts exceed 0.30. **c**, Reported interval span against reported AUC for the 1,069 identities whose interval limits were ordered and bracketed the reported AUC (0.002 × 0.002 bins). Median span falls from 0.184 at AUC 0–0.70 to 0.033 at 0.95–1.00 (Pearson *r* = −0.634). The mark at AUC 1.000 with span 0 carries 20 identities from six studies; nine one-sided and five point-outside identities sit on a separate rail. Intervals are spans as recorded, not verified 95% confidence intervals. In **b,c**, identities are reported measurements; studies are inference units. **d**, Membership of 138 studies across three predefined pools on a shared row axis: core (index-gated subclinical, early or forme fruste target; *n* = 74), reference/label (*n* = 51) and independent-positive (index-independent clinical target; *n* = 24). Color gives role side relative to each study’s target definition (truth-side 18, mixed 73, exclusion-side 24, unclear 23); role coding was invariant across pools within each study.

The five identifiable derivation studies contributed 32 claim units to the same claim-by-stratum audit; none supported VAE-NTT discrimination, and 26 of 32 (81.3%) exited at gate 1 [22,2,23,24,25] (Supplementary Table S5). The Corvis Biomechanical Index (CBI), for example, was derived by comparing keratoconic and normal eyes and provided no evidence for VAE-NTT discrimination [22].

The TBI derivation quantifies the mismatch between derivation task and target stratum: of 94 fellow eyes described as normal-appearing in its cohort, 40 (42.6%) were already flagged by BAD-D [2]. The cohort therefore did not represent a VAE-NTT stratum with dual certification: its target eyes met topographic normality criteria but were not certified as normal on tomography. One of its four adjudicated units exited at gate 1 and the other three at gate 2, so the reported derivation performance did not establish VAE-NTT performance. A direct evaluation of TBI in a stratum that excluded such flagged eyes produced an area under the receiver operating characteristic curve (AUC) of 0.35 [26].

### Deployed BAD-D and Ectasia Screening Index rules lacked a reconstructable public derivation chain

Where a derivation was identifiable, it was task-bound. BAD-D and the Ectasia Screening Index (ESI) instead had deployed rules that became reusable labels with no reconstructable public derivation chain. For BAD-D v2/v3, the deployed rule could be traced historically, but no public, peer-reviewed derivation sufficient to reproduce the algorithm was identifiable. The two nearest candidate sources were a 2012 monograph that could not be retrieved [27] and a 2009 trade publication retrieved from the manufacturer’s website [28]. The trade publication describes the display and its color thresholds at 1.6 and 2.6 standard deviations (s.d.) without a derivation (Supplementary Table S6). A paper on BAD-D v4 appeared in 2024, and none of its 12 claim units in this audit was supported [29]. BAD-D therefore left two distinct gaps: the deployed v2/v3 chain was not publicly reconstructable, and the later v4 paper did not establish VAE-NTT discrimination.

ESI showed a related supply-chain gap: a 2012 conference precursor is the leading candidate source, although it does not use the ESI name [30], and the paper that released the public source dataset states that its authors did not know the label-generating rule [31,32]. Ten studies applied ESI directly, and seven inherited its labels as ground truth through the public dataset; two studies belong to both groups (Supplementary Table S6). Once the labels entered a benchmark, unresolved provenance propagated: downstream studies repeatedly validated agreement with the inherited label and never independently re-established the clinical construct.

Herber et al. kept BAD-D among the tomographic exclusion criteria and took the detector from the biomechanical CBI of the Corvis ST [33], so BAD-D restricted the target stratum without generating the label. No unit from that study supplied a supported VAE-NTT result.

### Reported AUC varied within fixed contexts and carried its narrowest intervals where it was highest

Reported AUC varied within fixed measurement contexts: fixing the study, target, target contrast, evaluation dataset, partition and endpoint still left 286 measurement contexts in which two or more models or indices were reported. Within those contexts the median AUC span was 0.158, and 69 spanned more than 0.30 (Fig. 2b; all 3,084 AUC records are shown by target label in Fig. 3b and by resolution state in Supplementary Fig. 1). Reported interval width moved in the opposite direction to reported performance. Across the 1,069 records whose interval limits were ordered and bracketed the reported AUC, the median interval span fell from 0.184 at AUC 0–0.70 to 0.033 at AUC 0.95–1.00 (Pearson *r* = −0.634; Fig. 2c). Twenty records from six studies reported an AUC of 1.000 with [1.00, 1.00] intervals; 17 arose from pre-existing single-cohort evaluations, predominantly of established keratoconus. Extractable point AUCs accompanied 140 of 363 subclinical-target studies, and complete interval pairs were available for 1,074 of 2,785 resolvable AUC values (38.6%) in 68 of 154 studies. These patterns establish incomplete reporting and scope for selective emphasis.

**Fig. 3.**
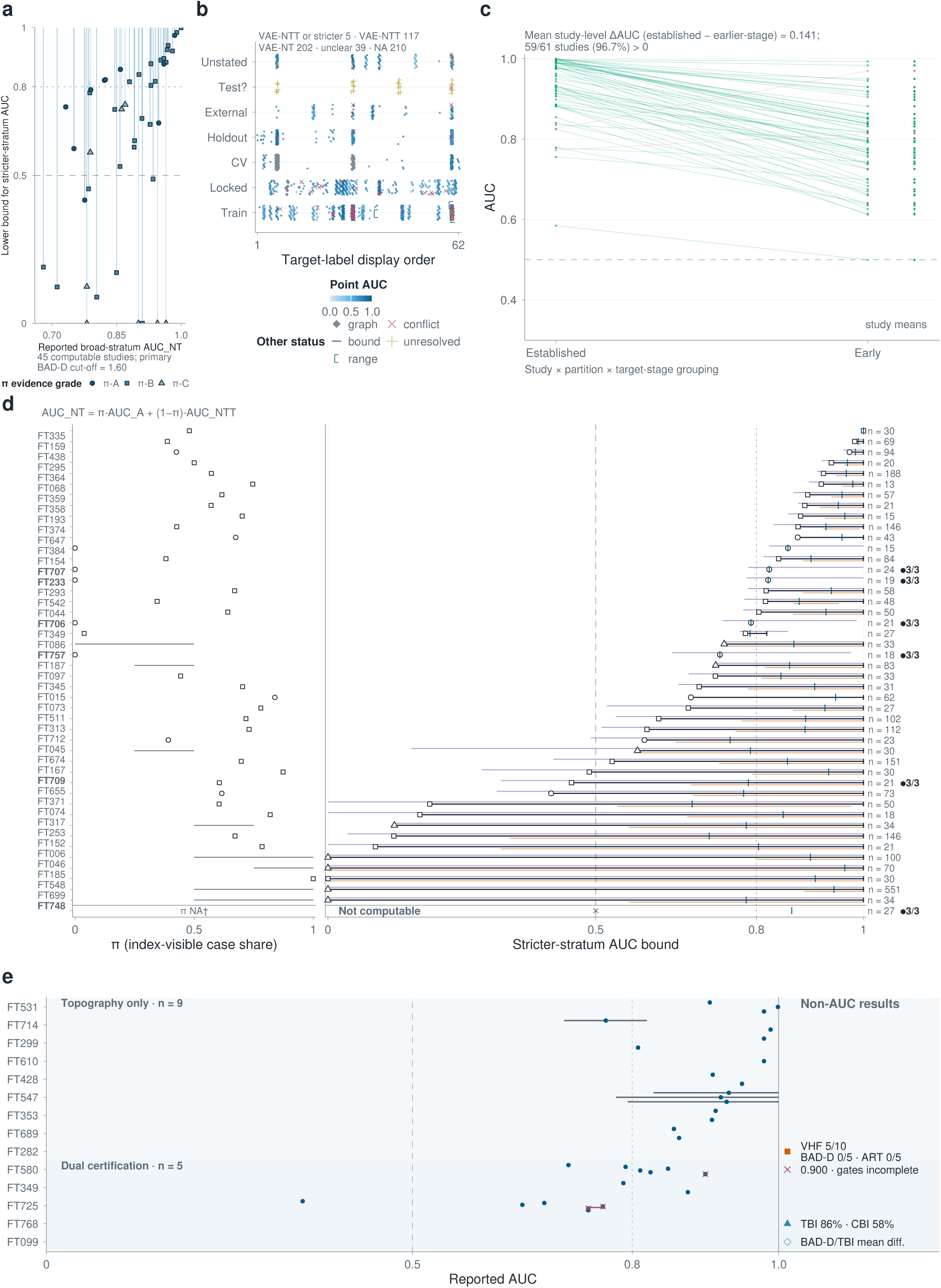
Partial identification bounds broad-stratum AUC away from stricter-stratum discrimination. AUC_NT = π·AUC_A + (1 − π)·AUC_NTT links broad-stratum (AUC_NT) and stricter-stratum (AUC_NTT) AUC; the index-visible case share π determines bound width. **a**, Reported AUC_NT versus the lower bound for stricter-stratum AUC in the 45 computable studies (primary BAD-D cut-off 1.60); markers sit at the lower bound, vertical lines span each study’s primary-threshold interval and marker shape gives the π evidence grade (π-A, π-B, π-C; Methods). **b**, 3,084 study-level AUC records, from 157 studies, across target labels and partitions (mutually exclusive study-level target categories, 573 studies: VAE-NTT or stricter 5, VAE-NTT 117, VAE-NT 202, unclear 39, NA 210); symbols mark graph-derived, conflicting, bound-only, unresolved and range-only records; target labels are in alphabetical order; Locked, locked pre-existing partitions; Test?, test partitions of unclear origin. **c**, 65 paired established-versus-earlier-stage partitions from 61 studies; each line is one partition pair, green where established-stage AUC exceeds earlier-stage AUC and pink otherwise; right-hand points are study means. Mean study-level ΔAUC (established − earlier-stage) is 0.141 (95% CI 0.120– 0.161, 2,000 study-level bootstrap resamples); 59 of 61 studies (96.7%) are above zero. **d**, Per-study bounds for the 46 qualifying studies (45 computable), ordered by descending primary lower bound. Left, π with grade-specific marks (circle, π-A; square, π-B; triangle, π-C); right, the stricter-stratum AUC bound at the primary 1.60 cut-off (black) with threshold-sensitivity bounds at 1.45 (purple) and 2.11 (orange), the reported AUC_NT (blue tick) and per-study *n*. Of the 45 lower bounds, 32 exceeded 0.50 and 18 exceeded 0.80; five intervals spanned [0, 1]. A filled circle with 3/3 marks the six studies for which the three Methods conditions were checked at full-text extraction. A dagger marks FT748 (FT identifiers denote corpus studies): outcome on a non-BAD-D scale, bounds not computable. **e**, Recognition evidence among the 14 index-exempt studies (9 topography-only, 5 dual certification): 31 AUC points with 4 reported 95% CIs; 3 non-AUC results and 1 gate note carry distinct markers (VHF, very high-frequency digital ultrasound; ART, Ambrósio Relational Thickness maximum).

Certainty, graded with the rubric described in Methods, was moderate for label–input independence, validation strictness, composite-index coverage and the controlled target-substitution estimate; low for normality verification, interval structure and subclinical point-AUC availability; and very low for pooled positive-class pre-selection, because 43 of its 54 contributing studies had mixed status (the Zenodo archive).

### Broad-stratum AUC did not identify VAE-NTT performance

Broad-stratum AUC is a composition-weighted mixture of index-visible and VAE-NTT performance, so it did not identify VAE-NTT performance without information on case composition, even when the reported AUC was treated as exact. A broad VAE-NT case group mixes two kinds of eyes, those already visible to the tomographic index and those normal on both topography and tomography. Let A denote the index-visible component and π its fraction among cases. For a fixed score and a common normal comparator, the broad-stratum AUC is the case-mixture average of the two components, AUC_NT = π·AUC_A + (1 − π)·AUC_NTT, which makes it a property of the case composition as much as of the score.

Nominally similar broad-stratum AUCs produced bounds ranging from informative floors to the entire [0, 1] interval (Fig. 3a, d; Supplementary Table S7). Forty-six of 573 studies met the partial-identification eligibility criteria, and 45 of them reported BAD-D-scale distributions sufficient to compute the bound at the prespecified threshold of 1.60; the remaining study reported the required distribution only on a tomographic scale incompatible with the BAD-D thresholds. At the prespecified tomographic threshold, 32 of 45 studies yielded an informative lower bound above 0.5 and 18 yielded a lower bound above 0.8. At the other extreme, 13 of 45 had conservative lower bounds at or below 0.5, and 5 had intervals spanning the entire [0, 1] range. For example, a reported AUC of 0.965 supported only by a distribution-free composition interval yielded [0.000, 1.000], whereas a reported AUC of 0.985 with direct eye-level counts yielded [0.974, 1.000].

The bottleneck was π, not the reported AUC: bound width tracked the evidence available on case composition. The median bound width was 0.013 in the 10 studies with direct eye-level counts (π-A), 0.195 in the 27 studies for which π was reconstructed from reported mean ± s.d. (π-B) and 0.938 in the 8 for which π was bounded by distribution-free intervals from reported quantiles (π-C). The three evidence grades for π were analyzed separately, in order of strength (Fig. 3). Results within the strongest evidence grade were heterogeneous: among the 10 π-A studies, the TBI derivation cohort had a stricter-stratum floor of 0.974 [2], the lowest floor was 0.417 [34], and 9 of the 10 had a floor above 0.5.

Twelve direct strict-stratum evaluations were available, 2 adverse and 10 indeterminate, with no supported VAE-NTT unit, so their AUCs are descriptive. Separately, the 14 index-exempt studies of the core pool (Fig. 2d), in which the index appeared only in normality certification and never in the positive-class definition, and discrimination was reported in certified-normal fellow-eye strata, entered a study-level comparison by certification depth (Fig. 3e). Among the nine topography-only studies, eight reported AUCs spanning 0.764–0.999; among the five studies with dual certification, three reported AUCs spanning 0.35–0.90, and the study contributing the upper value of 0.90 did not meet all audit gates. In this unadjusted contrast, both ends of the reported AUC range were lower under dual certification.

An AUC of 0.35 in a stratum defined by excluding eyes already flagged through the measurement pathway [26] measures discrimination in that stricter target. High broad-stratum AUC and low strict-stratum AUC can both be correct because they refer to different case compositions. Treating a broad-stratum AUC as the AUC for VAE-NTT is a performance-transfer error. The algebra established non-transferability but could not identify the source of the signal supporting the broad-stratum AUC, which required substituting the evaluation target while holding a frozen out-of-fold score and its comparator fixed.

### A fixed score lost discrimination as the target moved away from the label-generating pathway

Established-target AUC exceeded earlier-stage AUC in 59 of 61 published studies (96.7%) contributing 65 eligible partitions, with a study-weighted mean difference of 0.141 (95% confidence interval (CI): 0.120–0.161; Fig. 3c), across both development and evaluation designs; cohort, device and model also varied across these comparisons. A re-evaluation of the consensus criteria for subclinical keratoconus compared metric discrimination across 29 published studies without relabeling any eye [35]. Two reviewers checked 666 of 1,021 candidate pairs (40 of 59 studies); 24 pairs from 7 studies shared all eight registered identity fields, and in each both targets had been ascertained in the same tier (21 by device index, alone or with clinical findings, and 3 by clinical diagnosis), so none compared an index-defined with an independently ascertained target under one fixed model. A proxy-defined sensitivity analysis of 418 pairs from 43 studies estimated a target-stage AUC difference of 0.169 (95% CI: 0.147–0.192), with the established-stage AUC higher in every study (Methods). Fixed-score target substitution supplied the missing test.

With the source cohort, feature space, folds, model specification and comparator fixed, the frozen out-of-fold score fell from AUC 0.996 (95% CI: 0.994–0.998) for the operational ESI-early target to 0.682 (95% CI: 0.630–0.734) for FFearly and 0.549 (95% CI: 0.479–0.618) for the forme fruste keratoconus (FFKC) proxy [36] (Fig. 4a, b; Supplementary Table S8). The operational score had been trained only on the same-eye ESI-derived early label before target substitution. By design, ESI in the operational arm defined both the positive label and the exclusion rule for comparator eyes, so the case definition and the comparator selection shared one reference pathway. The first substitute, FFearly, kept the target eye ESI-normal and required the contralateral eye to be ESI-early; the second, the FFKC proxy, required the contralateral eye to be graded keratoconus by ESI, which placed it farther from the same-eye label-generating pathway. This 0.447 loss arose without refitting, site shift or model selection.

**Fig. 4.**
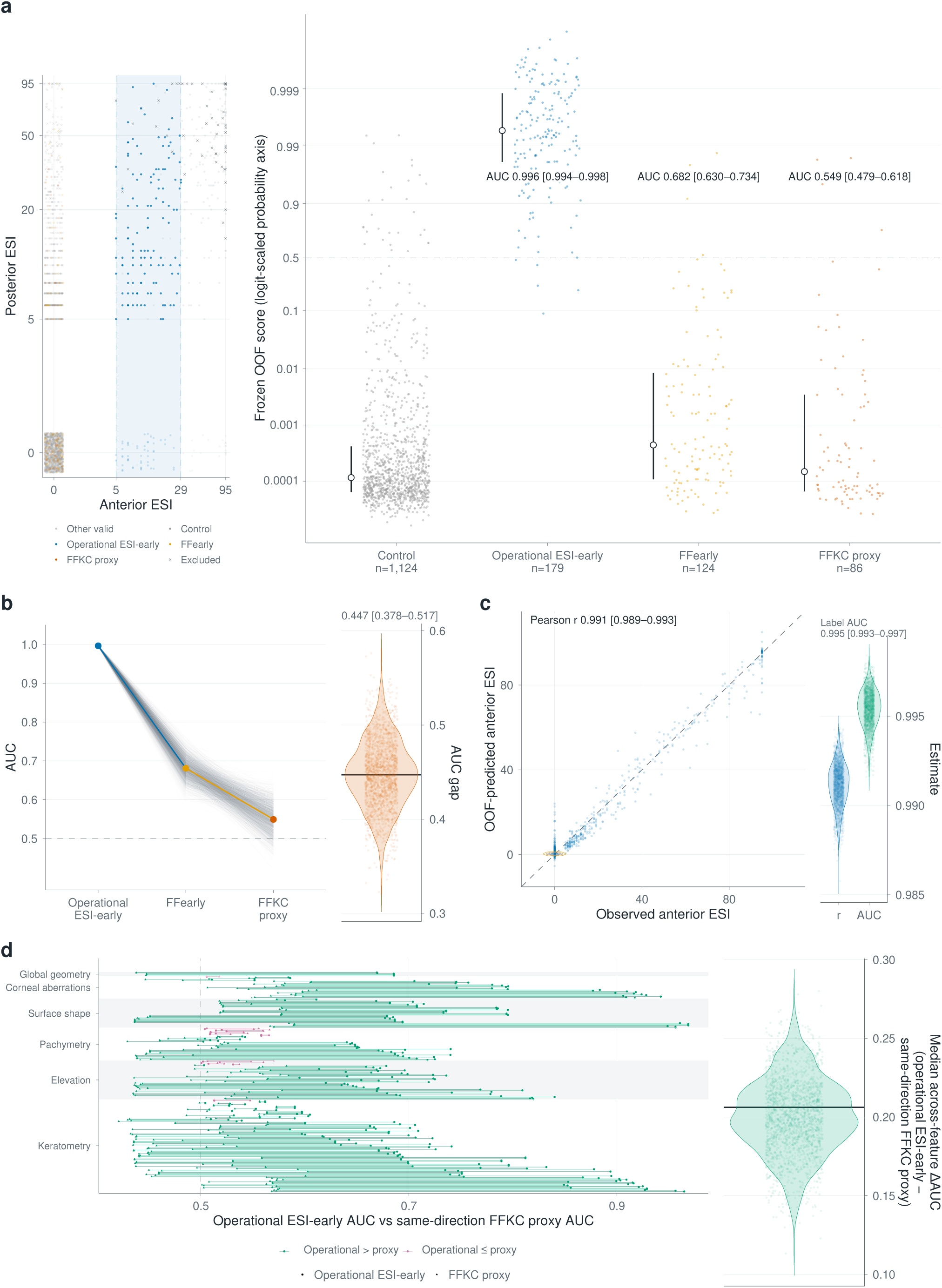
A frozen operational score loses discrimination as the target moves away from the label-generating pathway. **a**, Left, the label-generating ESI (Ectasia Screening Index) space of all 3,162 eyes (anterior versus posterior ESI, pseudo-log10 axes, sigma = 1; exact-zero values receive display-only jitter). The operational window spans anterior ESI 5–29. All three target comparisons use the same strict-control group. Control, FFearly and FFKC-proxy target eyes all have anterior ESI = 0 (control 1,124, FFearly 124, FFKC proxy 86); membership in the latter two groups is determined by the contralateral eye. Other valid eyes passing quality control (QC) outside the selected groups (*n* = 1,547); excluded, QC-failing eyes (*n* = 102); plotted points require finite anterior and posterior ESI. Right, the frozen out-of-fold (OOF) operational-label score for all 1,513 comparison eyes on a logit-scaled probability axis, by target group (hollow circle, median; whisker, interquartile range). AUC is 0.996 (95% CI 0.994–0.998) for operational ESI-early, 0.682 (0.630–0.734) for FFearly (defined by the contralateral eye’s vendor output) and 0.549 (0.479–0.618) for the FFKC (forme fruste keratoconus) proxy; dashed lines at 0.5 are visual references. **b**, The three AUCs and the Target-Swap Gap (operational − FFKC proxy = 0.447, 95% CI 0.378–0.517) across 2,000 joint patient-level bootstrap replicates. **c**, A model using the 298 retained native features (Native-298) reconstructs anterior ESI across 3,060 eyes: out-of-fold-predicted versus observed anterior ESI (Pearson *r* = 0.991, 95% CI 0.989–0.993); the reconstructed index recovers the operational label with AUC 0.995 (0.993–0.997); violins show the 2,000 bootstrap replicates. Contour lines trace two-dimensional point density, colored from orange (low) to blue (high); the dashed diagonal is the identity line. **d**, Per-feature target-swap gaps: 278 of 298 features (93.3%) show a positive operational-to-proxy AUC gap (feature direction fixed on the operational comparison; ties excluded), grouped into six descriptive feature families; the right violin gives the across-feature median ΔAUC over 2,000 joint bootstrap replicates.

Fixed-threshold performance showed the same target dependence. At the threshold selected for empirical specificity of at least 95%, operational sensitivity was 0.994 and specificity 0.953 (178 of 179 target eyes; 1,044 of 1,095 strict-control eyes). Applied unchanged, that threshold detected 0.132 (95% CI: 0.056–0.195) of FFearly eyes and 0.136 (95% CI: 0.046–0.222) of FFKC-proxy eyes. At the threshold selected for at least 99% specificity, operational sensitivity was 0.855 and specificity 0.991, and transported sensitivity was 0.018 (95% CI: 0.000–0.070) for FFearly and 0.000 (95% CI: 0.000– 0.043) for the FFKC proxy. Operating rules selected on the same-eye label retained little sensitivity for the substituted targets.

Reconstruction of the label-generating index was sufficient to reproduce near-perfect discrimination of the operational target. Although vendor-output columns were excluded from the inputs by name, a model using the 298 retained features reconstructed anterior ESI with *r* = 0.991 (95% CI: 0.989– 0.993), and the reconstructed index recovered the operational label with AUC 0.995 (95% CI: 0.993– 0.997) (Fig. 4c). At the feature level, 278 of 298 individual features (93.3%) had positive operational-to-proxy target-swap gaps (Fig. 4d). This input–label configuration is reference-pathway leakage: when the remaining measurements can reconstruct the label-generating rule, deleting the named index column does not remove the information that generates the label.

Direct learnability and score transport answered different questions: refitting asked whether the feature space could learn the FFKC proxy directly, whereas transport asked whether the frozen operational score already discriminated it. Refitting yielded AUC 0.529 (95% CI: 0.461–0.600), within the permutation null distribution (two-sided add-one *P* = 0.551, 1,000 fold-preserving permutations), while transport of the frozen operational score yielded AUC 0.549 (95% CI: 0.479–0.618). The Target-Swap Gap, operational AUC minus transport AUC, was 0.447 (95% CI: 0.378–0.517). Independent-proxy Target Retention, the transported above-chance discrimination as a fraction of the operational above-chance discrimination, was 0.099 (95% CI: −0.042 to +0.238). Neither refitting nor transport recovered discrimination against the FFKC proxy.

### Positive controls preserved manifest-disease performance, while demographics provided an alternative route to discrimination

On the eight features shared by the two devices, manifest keratoconus remained separable across them (AUC 0.945–0.993), whereas early and subclinical targets yielded AUCs of 0.677–0.759 (Fig. 5a; Supplementary Table S9). The independent Almería Pentacam cohort provided a second positive control: 40 non-composite same-device measurements from the BAD-D measurement family discriminated the pre-labeled subclinical grade with AUC 0.940 (95% CI: 0.880–0.983), and BAD-D alone reached 0.933 (Fig. 5b). Adding BAD-D to the 40 measurements did not measurably change discrimination (−0.003; 95% CI: −0.031 to +0.019). The same cross-fitted specifications separated manifest keratoconus from controls with AUC 0.994 from the 40 measurements alone and 1.000 after adding BAD-D. The same 40 measurements predicted BAD-D with an out-of-fold Pearson correlation of 0.956 (2,000-resample percentile interval 0.937–0.981), root mean squared error 1.303 (0.798–1.856), mean absolute error 0.691 (0.551–0.862) and R² 0.911 (0.863–0.961) across 205 eyes. In this cohort as well, the composite index was recoverable from its own measurement family.

**Fig. 5.**
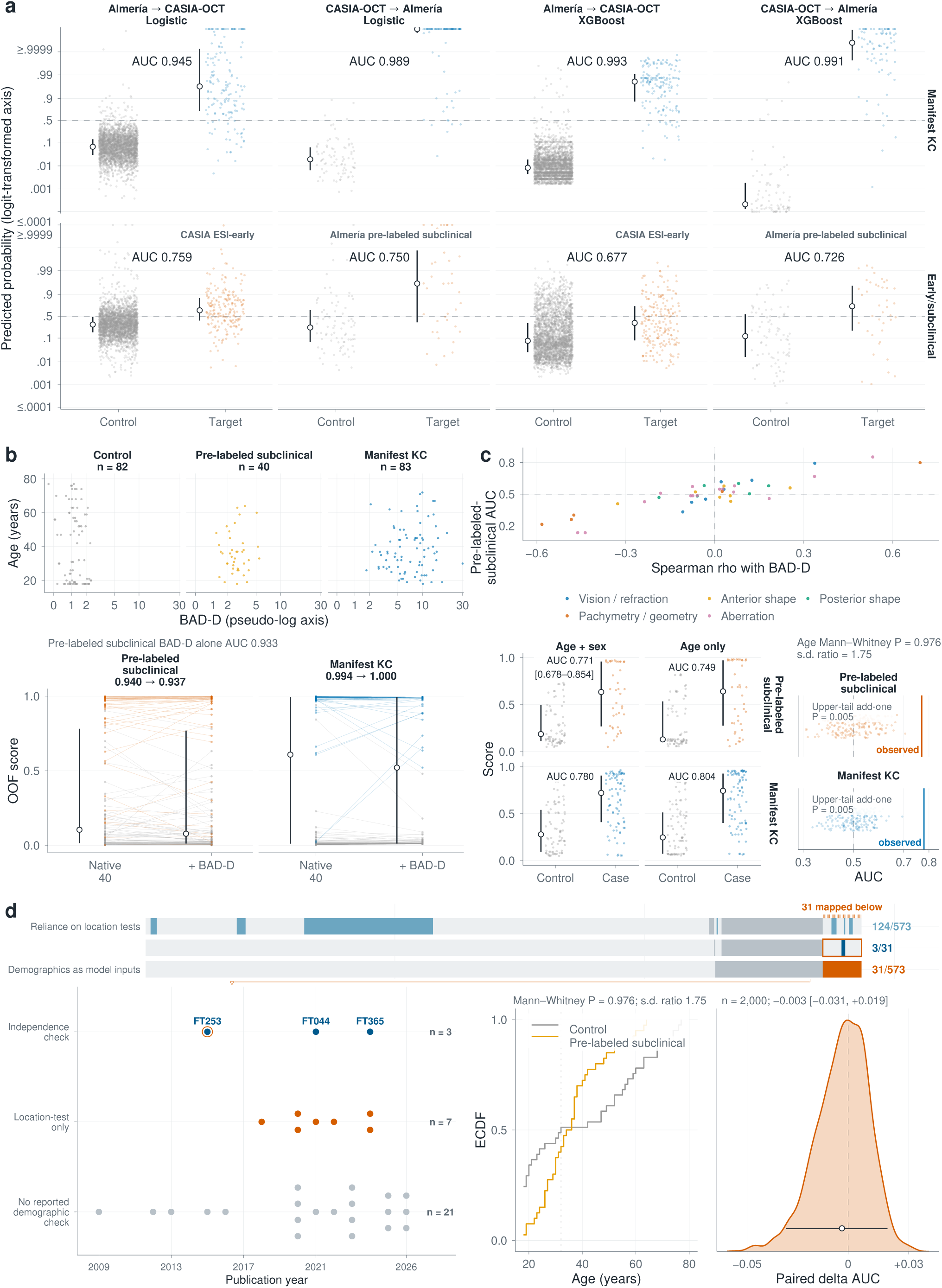
Manifest-disease performance and demographic structure provide discrimination routes that do not require the subclinical target. **a**, Almería (Pentacam)–CASIA-OCT transport using logistic and XGBoost models: predicted probability (logit-transformed axis; extremes display-clipped at 0.0001/0.9999, marked at end ticks) for control versus target eyes (CASIA: 162 manifest, 199 ESI-early, 2,699 controls; Almería: 83 manifest, 40 pre-labeled subclinical, 82 controls). Manifest keratoconus (KC) remains separable in both directions and models (AUC 0.945, 0.989, 0.993, 0.991), whereas early/subclinical targets (CASIA ESI-early; Almería pre-labeled subclinical) yield AUC 0.759, 0.750, 0.677, 0.726. Hollow circle, median; vertical line, interquartile range; dashed line at 0.5. **b**, Top, 205 Almería eyes by age and BAD-D (BAD-D on a pseudo-log axis, asinh(x/2)/log(10), ticks at original values). Bottom, same-device cross-fitted out-of-fold (OOF) scores from 40 non-composite measurements with and without BAD-D; lines connect paired scores: pre-labeled subclinical AUC 0.940 → 0.937 (BAD-D alone 0.933); manifest 0.994 → 1.000 (82 controls versus 83 manifest eyes). **c**, Top, Spearman correlations with BAD-D versus pre-labeled-subclinical AUC, by feature family. Bottom, cross-fitted scores using age + sex (pre-labeled subclinical AUC 0.771, 95% CI 0.678– 0.854 from 2,000 eye-level bootstrap resamples; manifest 0.780) and age only (0.749; manifest 0.804), with permutation nulls (200 label permutations; upper-tail add-one *P* = 0.005 for both targets; two-sided Mann–Whitney age *P* = 0.976; control-to-case age s.d. ratio 1.75). **d**, Demographic inputs and checks across 573 studies (top, one column per study; dark gray, unclear; light gray, other/none; lexicographic order by check field): 124 used location tests, 31 used demographic inputs, and 3 of 31 tested discrimination independent of demographics (one also reported a location test). Below, 31 demographic-input studies by year and check; Almería control and pre-labeled subclinical age empirical cumulative distribution functions (ECDF) (dotted lines, medians); and paired ΔAUC from adding BAD-D (−0.003, 95% CI −0.031 to +0.019; 2,000 eye-level resamples; white point, estimate; black segment, 95% percentile interval).

Age and sex alone separated the pre-labeled subclinical grade from controls in the second cohort (AUC 0.771, 95% CI: 0.678–0.854; upper-tail add-one permutation *P* = 0.005 from 200 label permutations; Fig. 5c). The same model also separated manifest keratoconus from controls (AUC 0.780; permutation *P* = 0.005). The univariate age comparison did not detect a distributional shift (two-sided Mann–Whitney U *P* = 0.976), whereas the control-to-case age s.d. ratio was 1.75. The age-plus-sex model separated groups that the rank comparison could not distinguish; an undetected shift did not imply similar distributions. Across the 573-study corpus, 31 studies (5.4%) entered age or sex directly into the model inputs, but only 3 of these 31 tested whether model discrimination was independent of demographics (Fig. 5d). Separately, 124 of 573 relied on location tests, which do not assess dispersion or joint structure; eight overlapped the 31 demographic-input studies, including seven with location tests but no independence check.

Across the two datasets, two routes to discrimination did not require detection of the intended subclinical corneal target: reconstruction of the label-generating index in one cohort and demographic group structure in the other. In CASIA, discrimination against the most separated proxy remained indistinguishable from chance. Both routes are compatible with the benchmark label, so testing clinical detection requires a target outside the benchmark’s label-generating reference pathway.

## Discussion

Published keratoconus AI has not established the capability for which it is cited, discrimination in fellow eyes that are normal on both topography and tomography: no VAE-NTT unit among 2,035 units from 212 adjudicated studies was supported. Index tracing showed that identifiable derivations addressed other target populations and that the deployed BAD-D and ESI rules lacked reconstructable public derivation chains, and partial identification showed that broad-stratum performance did not identify performance in the stricter stratum. In the experiment, a frozen score fell from AUC 0.996 for the same-eye ESI label to 0.549 for the most separated proxy as the target moved away from the label-generating pathway within one public cohort, and index reconstruction reproduced near-perfect discrimination of the operational label. In a second cohort, age and sex alone separated the pre-labeled subclinical grade from controls. Diagnostic accuracy thus behaved as a quantity jointly determined by the target stratum and the reference pathway that defined the label, not as a portable property of a model.

In the three-center test set of one study, BAD-D already flagged 29 of the 43 case eyes that the study labeled preclinical keratoconus, so the AUC of 0.96 for these cases versus normal eyes was measured in a mixed stratum [37]. The study was one of the 10 π-A studies with eye-level counts complete enough to apply the mixture identity. Under the mixture identity, this broad-stratum AUC bounds stricter-stratum performance without establishing it, as do the other broad-stratum AUCs in Fig. 3.

Reference-pathway leakage is governed by index role, not index presence. Used as an exclusion ruler, an index defines the question; used as the source of the positive label, it answers the question in advance. When the model’s inputs can also reconstruct it, the claim is capped at recovery of the reference rule. The practical rule is to define positive-class identity using evidence outside the model-input measurement pathway. Herber et al. sourced that identity from clinical keratoconus in the contralateral eye and paired tomographic exclusion criteria with a biomechanical detector [33].

Site generalization and target–measurement separation are separate axes. The former asks whether performance transports beyond the development data; the latter describes the relation between the model inputs and the evidence defining the clinical construct. Of 54 externally validated studies, 17 were extracted as constructing VAE-NTT or a stricter target, and 6 of those 17 as defining the positive class clinically; three of the six met that criterion in full on cell-level review (Table 1). Generalization across sites that share one reference pathway leaves target–measurement separation untested, which is how appraisal systems credited external validation while leaving that separation unscored.

The anchor gap was visible in the secondary literature, but the appraisal instruments as applied did not score it. Five systematic reviews and meta-analyses appraised this literature between 2022 and 2024 with QUADAS-2, the Quality Assessment of Prognostic Accuracy Studies (QUAPAS), the TRIPOD statement or Critical Appraisal Skills Programme (CASP) checklists, covering 19 to 93 primary studies each. None reported an item assessing whether the reference label was computed from the measurements read by the model [38,10,39,40,41], and a sixth review that applied a tailored QUADAS-2 rated reference-standard bias as unclear for every included study because no gold standard for subclinical keratoconus exists [42]. The Cochrane review came closest to the mechanism of reference-pathway leakage. It removed the QUADAS-2 timing question because the reference and index tests were, in its words, “performed on the same corneal images or parameters,” recording the structural overlap as a procedural exemption rather than as an object of assessment [38]. Its remaining reference-standard questions asked who interpreted the images and whether interpretation was blinded, not where the label came from. Another review found no significant association between its quality scores and reported accuracy in meta-regression (all *P* > 0.05) and still concluded that the models improve discrimination [41]. Appraisal instruments treat the reference standard as a per-study quality attribute, while quantitative synthesis pools unstratified AUCs, leaving the design flag disconnected from the performance estimate.

Anchor-gap evaluation changes where accuracy is measured and against what. Positive-class identity comes from evidence outside the model-input measurement pathway, in keratoconus the clinical diagnosis of the contralateral eye, which is already in the chart and requires no additional device or follow-up visit; the target eye is certified normal on both topography and tomography; and the frozen score and its operating threshold are carried unchanged onto those eyes. The fall from headline to transported accuracy, the Target-Swap Gap, then shows how much of the headline was agreement with the reference index, and Independent-proxy Target Retention shows how much discrimination survives. The experiment made this change within one cohort: moving the label from the scored eye’s own ESI grade to its fellow eye’s took the frozen score from AUC 0.996 to 0.549 and sensitivity at the 99%-specificity threshold from 0.855 to 0.000. Three disclosures record whether a study made the change: the source of positive-class identity; the measurements used to certify the target eye; and any measurement pathway still shared by the label, the exclusion criteria and the model inputs. Where target substitution is not feasible, these disclosures still show whether reference-pathway leakage remains possible. Orthogonal measurement breaks the shared reference pathway and strengthens the claim when it is available.

Table 2 converts those disclosures into six reporting items: three mandatory items that define the claim (the claim stratum, a clinically sourced positive-class identity outside the model inputs, and per-eye reporting of the exclusion rulers) and three extension modules (orthogonal measurement, prespecified target transport and demographic separation). The ESI-early→FFearly→FFKC-proxy result provides the transport example, with refit as a separate estimand; its most separated proxy remained within the vendor ESI lineage, whereas the audit treated a later clinical outcome as the most separated anchor. Each unmet item narrows the claim a performance estimate supports, which is why anchor-gap evaluation operates at the level of the claim rather than the study. Across the seven studies in Table 1, six external-validation candidates and one single-site architecture example, two designs met all three core items: Chan et al. with external transport but a shared measurement pathway, and Herber et al. with an orthogonal detector in a single-site design [33] (Table 3; cell-level evidence in Supplementary Table S10). No corpus study demonstrated fixed-score target transport across cohorts; the nearest equivalent was the fixed CBI of Herber et al. evaluated against the same normal eyes in the VAE-NT and VAE-NTT strata (AUC 0.992 and 0.950), a within-cohort gradient in the direction the experiment reproduces, and pathway orthogonality appeared only in that single-site architecture example.

**Table 2.**
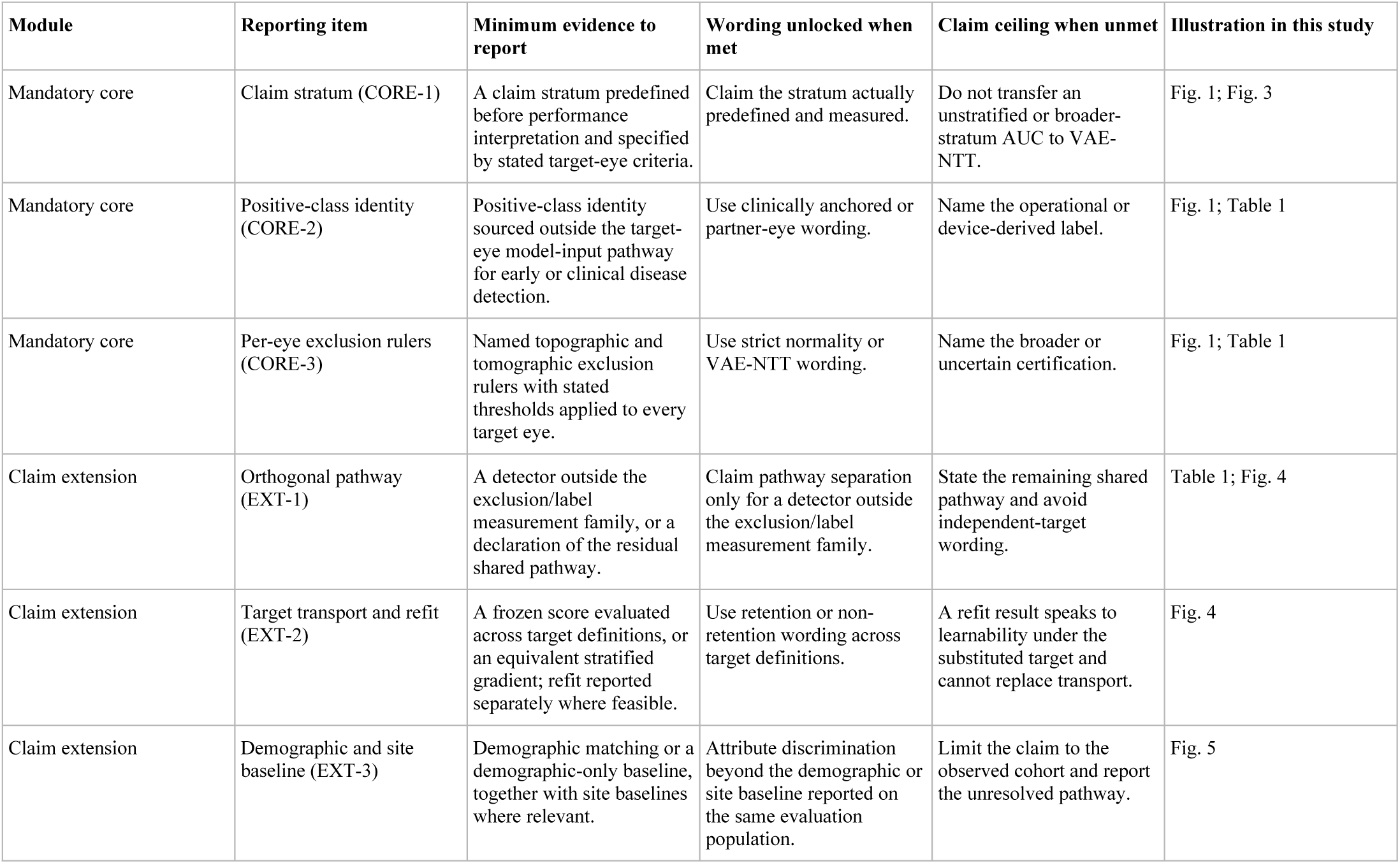
Anchor-gap evaluation: six reporting items. Abbreviations: AUC, area under the receiver operating characteristic curve; VAE-NTT, very asymmetric ectasia with normal topography and tomography.

| Module | Reporting item | Minimum evidence to report | Wording unlocked when met | Claim ceiling when unmet | Illustration in this study |
| --- | --- | --- | --- | --- | --- |
| Mandatory core | Claim stratum (CORE-1) | A claim stratum predefined before performance interpretation and specified by stated target-eye criteria. | Claim the stratum actually predefined and measured. | Do not transfer an unstratified or broader-stratum AUC to VAE-NTT. | Fig. 1; Fig. 3 |
| Mandatory core | Positive-class identity (CORE-2) | Positive-class identity sourced outside the target-eye model-input pathway for early or clinical disease detection. | Use clinically anchored or partner-eye wording. | Name the operational or device-derived label. | Fig. 1; Table 1 |
| Mandatory core | Per-eye exclusion rulers (CORE-3) | Named topographic and tomographic exclusion rulers with stated thresholds applied to every target eye. | Use strict normality or VAE-NTT wording. | Name the broader or uncertain certification. | Fig. 1; Table 1 |
| Claim extension | Orthogonal pathway (EXT-1) | A detector outside the exclusion/label measurement family, or a declaration of the residual shared pathway. | Claim pathway separation only for a detector outside the exclusion/label measurement family. | State the remaining shared pathway and avoid independent-target wording. | Table 1; Fig. 4 |
| Claim extension | Target transport and refit (EXT-2) | A frozen score evaluated across target definitions, or an equivalent stratified gradient; refit reported separately where feasible. | Use retention or non-retention wording across target definitions. | A refit result speaks to learnability under the substituted target and cannot replace transport. | Fig. 4 |
| Claim extension | Demographic and site baseline (EXT-3) | Demographic matching or a demographic-only baseline, together with site baselines where relevant. | Attribute discrimination beyond the demographic or site baseline reported on the same evaluation population. | Limit the claim to the observed cohort and report the unresolved pathway. | Fig. 5 |

**Table 3.** Application of the six reporting items to seven studies. For CORE-1/2/3, EXT-2, and EXT-3, states are Met, Conditional, Not demonstrated, and Not reported. EXT-1 uses Orthogonal, Shared, and Not reported. Herber et al., 2022 is the single-site architecture example from Table 1, not one of the six true-external candidates. Per-cell evidence statements with source excerpts are given in Supplementary Table S10.

| Module | Reporting item | Karimi et al., 2018 (FT126) | Issarti et al., 2020 (FT512) | Wahba et al., 2018 (FT666) | Saad & Gatinel, 2012 (FT686) | Ruiz Hidalgo et al., 2017 (FT687) | Chan et al., 2015 (FT688) | Herber et al., 2022 (FT378) |
| --- | --- | --- | --- | --- | --- | --- | --- | --- |
| Mandatory core | Claim stratum (CORE-1) | Conditional | Met | Met | Conditional | Conditional | Met | Met |
| Mandatory core | Positive-class identity (CORE-2) | Conditional | Met | Met | Not demonstrated | Conditional | Met | Met |
| Mandatory core | Per-eye exclusion rulers (CORE-3) | Not reported | Not demonstrated | Conditional | Not demonstrated | Not reported | Met | Met |
| Claim extension | Orthogonal pathway (EXT-1) | Shared | Shared | Shared | Shared | Not reported | Shared | Orthogonal |
| Claim extension | Target transport and refit (EXT-2) | Not demonstrated | Not demonstrated | Not reported | Not demonstrated | Not demonstrated | Not demonstrated | Met |
| Claim extension | Demographic and site baseline (EXT-3) | Not demonstrated | Conditional | Not demonstrated | Conditional | Conditional | Not demonstrated | Conditional |

In keratoconus, reference-pathway leakage operated through a benchmark supply chain spanning vendor development, deployment thresholds, dataset release, label inheritance and downstream revalidation. In this corpus, one vendor index entered a public dataset, the paper releasing that dataset states that the generation rule was not known to its authors [32], and seven downstream studies inherited those labels as ground truth. Imaging models trained on public datasets that carry device-derived labels are exposed to the same chain wherever one exists, and lineage and target-transport analyses can test it in other domains. Reference recoverability can be audited without the original derivation. In the CASIA cohort, the observed index values supplied the reconstruction target, and the reconstructed index recovered the operational label (Fig. 4c). Wherever reference outputs and candidate model inputs are both observable, the same test applies to a closed reference.

A label record should preserve the role of each measurement: the clinical or device source of positive-class identity, the eye from which identity was assigned, the reference version and thresholds, and the measurements used to certify the target eye. Reuse then carries the label’s operational meaning, so downstream accuracy claims can be read against the construct that the record documents. Patient-level linkage between the target and contralateral eyes should be preserved as well, so that a target definition can be rebuilt from its recorded evidence and evaluated with patient-grouped partitions.

Anchor-gap evaluation defines a performance unit that a downstream decision-maker can verify. Because accuracy is jointly determined by target stratum and reference pathway, regulators, guideline developers and purchasers should match each performance estimate to the stratum and pathway in which it was measured. An unstratified AUC cannot, by itself, support a claim for a stricter clinical target. This reporting unit can be recorded in a regulatory label claim, guideline table or procurement checklist and requires no access to code or data. Because changing the label-generating algorithm, its threshold or the target-eye eligibility rules changes the evaluated target even when the model is unchanged, the reference rule and its version belong in the claim alongside the model and the intended population.

A trial that claims detection beyond routine topography and tomography evaluates that claim in the prespecified instrument-negative stratum, in which the contralateral clinical diagnosis establishes cross-sectional membership in the high-risk group. Sensitivity at the intended operating threshold addresses identification of that high-risk group; overall AUC describes discrimination across the enrolled case mixture.

For a fellow eye that is normal on both topography and tomography, the relevant screening evidence is performance in that same stratum, and the published literature has not yet supplied it, so no published keratoconus AI model can be used to clear such an eye. The evidence that would support clearing it is sensitivity at a fixed operating threshold in that stratum, against the contralateral clinical diagnosis already in the chart, and two published designs already take positive-class identity from a clinical source outside the model inputs. A study reported that way either supplies the missing evidence or shows, as the target swap did here, how much of a headline AUC was agreement with the device index that defined its label.

## Methods

### Study design and registration

This study combined a registered systematic audit of the published literature with a fixed-score target-substitution experiment on a public dataset and a demographic stress test on a second independent public cohort. The review was registered on PROSPERO (CRD420261441197), and protocol amendments and deviations from the registered search are reported in the Zenodo archive (Data availability) and the Preferred Reporting Items for Systematic Reviews and Meta-Analyses (PRISMA) flow (Supplementary Fig. 2). We assigned an adverse finding only when the underlying design or reporting choice was avoidable at zero or low cost at the time of publication; conditions requiring a second device, multicenter resources or longitudinal follow-up were coded as future claim-strengthening standards. The study analyzed published literature and two publicly released datasets and collected no new data from human participants, so ethics approval was not required.

One included study was co-authored by members of this team [37]; it entered through the registered search and was coded under the same workflow. Two raters who had not participated in that study (F.G. and L.Y.) independently readjudicated all 10 of its claim units, with blinded resolution by a third rater (Ying Jie); none was supported. This recusal exercise remained outside the 212-study adjudicated set and is reported in the Zenodo archive.

### Search and corpus construction

Six sources, PubMed/MEDLINE, Embase, Scopus, the Web of Science Core Collection, Europe PMC and OpenAlex, were searched from inception within the registered search boundary of 27 June 2026, without language restriction, using the registered two-arm strategy. Source-level yields are provided in Supplementary Table S1; the Zenodo archive holds the executed queries for PubMed, Embase, Scopus and Web of Science and the query components for Europe PMC and OpenAlex, whose exact query strings were not retained. The PubMed search is documented by its recorded execution date (4 July 2026) and the retained list of 2,090 PMIDs; exports from the other five sources carry no execution date, a deviation recorded in the registration. Eligible studies addressed keratoconus screening by developing or validating classifiers based on artificial intelligence (AI), machine learning or statistical methods, or by evaluating a named fixed diagnostic index. Full text was obtained for all 573 included studies; unretrieved candidate reports are listed separately in the PRISMA flow [43]. Deduplication used digital object identifier (DOI) matching supplemented by full-text content hashing, which identified four duplicates whose full texts contained no DOI.

### Claim-by-stratum extraction and adjudication

Unit-level adjudication covered the 212 studies whose extracted claims asserted discrimination in the review’s target population; the 361 studies making no such claim formed the verification pool described below. Each adjudicated study was decomposed into claim-by-stratum units (one unit for each pairing of a performance claim with a target stratum), yielding 2,035 units across the 212 studies. Six gates were applied at the unit level; this framework replaced the registered study-level two-field extraction and is reported in the registration record as an analysis added after version 2.0. Gate 0 (task) assessed claim existence and scope. Gate 1 (population) assessed target-population construction and normality certification. Gate 2 (identity and reference) assessed the source of positive-class identity, target-eye ground truth and class-wise verification. Gate 3 (inputs) assessed eligibility records, headline inputs and data partitions, including participant or eye overlap across partitions. Gate 4 (evidence strength) assessed the adequacy of data sources, splits and scoring procedures for the headline estimate. Gate 5 (alignment) assessed agreement between the stated claim, the measured estimand and the implied clinical action. Full orthogonality of the measurement pathway was recorded descriptively as a condition for future designs; adverse verdicts depended on gates 0–5.

For every unit, the source of positive-class identity and the ground truth applied to the target eye were coded separately. Reference-pathway overlap between the label and model inputs was assigned one of five pathway-overlap grades (exact index, deterministic transform, same-device raw ancestor, independent-device feature and disjoint), ranging from exact overlap with a same-eye index pathway to complete disjointness, and this grade determined what evidence a study was required to provide.

The original and supplemental extraction sets were independently double-coded by two reviewers (H.J. and F.G.), with disagreements adjudicated by a third (Yang Jiang). Agreement after code normalization was 90.6% and 75.8%, respectively; the supplemental set consisted predominantly of citation-recovered studies with sparser reporting (Supplementary Fig. 3), and the two sets were analyzed separately.

### Verification subsample for non-adjudicated studies

The non-adjudicated studies formed the target-stratum verification pool and were assessed by stratified blinded sampling rather than unit-level adjudication. At the time of sampling, this pool comprised a random sampling frame of 394 studies (209 previously uncoded, 158 VAE-NT and 27 unclear-stratum studies) together with 30 studies verified earlier. From the frame, 122 studies were sampled and independently recoded by two blinded raters under a prespecified protocol, with disagreements resolved by a third rater. The random verification sample comprised 60 previously uncoded studies, 35 VAE-NT studies and all 27 unclear-stratum studies; incorporating 25 VAE-NT and 5 unclear-stratum studies verified earlier yielded stratum totals of 60, 60 and 32. Using only the prespecified random samples, we obtained a one-sided joint 95% upper confidence limit of 24 for the number of VAE-NTT studies among the 272 frame studies remaining unchecked, by finite-population hypergeometric inversion with Bonferroni adjustment across the non-census layers. Previously verified nonrandom studies were treated only as known negatives. Adjudication continued after sampling, so the final non-adjudicated pool holds 361 studies, 110 of which belong to the verification sample; the limit is reported for the frame in which the sample was drawn. The layer-wise inputs are reported in Supplementary Table S4, together with a binomial (Clopper–Pearson) projection of the same data and the earlier rule-of-three calculation.

### Index provenance audit

We audited each mention of seven prespecified composite indices across the corpus, distinguishing evaluation of an index from its use as a reference standard, label source or eligibility criterion (420 evidence rows, Zenodo archive). Indices with at least 10 qualifying uses were designated load-bearing, and six met this threshold. For each index, derivation records were traced to primary sources and classified as verified against the original document, candidate precursor with unconfirmed identity, or unverified. Unverified sources did not inform inference; all provenance grades appear in Supplementary Table S6.

### Partial identification across target strata

The mixture identity was applied only under three conditions: the same fixed score, the same normal comparator, and a broad case stratum that could be partitioned into an index-visible component (A) and a VAE-NTT component. Under these conditions, AUC_NT = π·AUC_A + (1 − π)·AUC_NTT bounds the stricter-stratum AUC within that study, following the logic of partial identification [44]. π was graded by evidence: direct eye-level counts (π-A); a normal approximation from reported mean ± s.d. (π-B); and distribution-free intervals from reported quantiles (π-C). Each qualifying study’s reported AUC and distributional summaries were taken at face value. The tomographic suspicion threshold was frozen in advance at BAD-D 1.60, with 1.45 and 2.11 as sensitivity values. Forty-six of 573 studies met the eligibility criteria; 45 provided BAD-D-scale distributions for bound calculation, and the remaining study reported the required distribution only on an incompatible tomographic scale (Supplementary Table S7). When π was interval-valued, we reported the widest stricter-stratum interval consistent with that range.

### Target-stage directionality comparison

For target-stage directionality, we included only AUC estimates reported as numerical point values from study–partition combinations containing both an established-keratoconus target and an earlier-stage or subclinical target within one of six explicit partition classes: train/apparent, internal-holdout, cross-validation-mean, locked-preexisting/single-cohort-evaluation, test/external and test/unclear-origin. Non-point estimates, target labels outside the fixed established/earlier-stage mapping and partitions labeled “not stated” or otherwise ambiguous were excluded. Within each study × partition × stage, eligible AUC estimates were averaged equally, and each partition contrast was the established-stage mean minus the earlier-stage mean; multiple partition contrasts were then averaged equally within study. The headline mean weighted studies equally, and a study was counted as positive when its study-level contrast exceeded zero. The 95% CI was estimated from 2,000 bootstrap resamples of studies.

### Registered natural target-swap adjudication

The registered synthesis of published natural target swaps required candidate AUC pairs to share study, evaluation dataset, partition, model or index, model configuration, endpoint, input set and comparator, to differ only in target, and to draw the two targets from different ascertainment tiers: an operational target whose positive class was assigned by a target-eye device index, alone or combined with clinical findings, and a substituted target assigned by independent clinical or longitudinal diagnosis or by partner-eye status without a target-eye index gate. We assembled 1,021 candidate pairs from 59 studies. For 666 pairs from 40 of these studies, two reviewers independently checked the eight identity fields against the full text, with disagreements resolved by a third; 24 pairs from 7 studies shared all eight fields, and the input set could not be determined for 641. The same procedure assigned the ascertainment tier of both targets in these 24 pairs: 21 were operational at both ends and 3 were clinical at both ends, so none of the checked pairs was eligible and the Target-Swap Gap and Independent-proxy Target Retention could not be computed from them. The 355 pairs from the remaining 19 studies were not checked. In a proxy sensitivity analysis, 418 pairs from 43 studies that shared a resolvable comparator and mapped to an established-stage and an earlier-stage target term were retained; pair differences (established-stage minus earlier-stage AUC) were averaged within study and then equally across the 43 studies, with a 2,000-replicate study-level bootstrap interval. Pair-level records are in the Zenodo archive.

### Controlled target-substitution experiment

The experiment used the public CASIA anterior-segment optical coherence tomography cohort from Harvard Dataverse (CC0), which includes patient-level linkage identifiers [31,32]. Eye grades in this cohort follow the vendor anterior ESI (ESI-normal, 0–4, with every observed normal value equal to 0; ESI-early, 5–29; ESI-KC, 30 or above), and the CASIA manifest-keratoconus label used below is the ESI-KC grade. We retained 298 native morphological features per eye and excluded the named vendor-output columns, ESI, the keratoconus index (KCI), the keratoconus prediction index (KPI) and any label-derived score. The retained morphological measurements remained available to the model and supplied the inputs for the index-reconstruction analysis.

The classifier was an XGBoost gradient-boosted tree model (XGBoost 3.3.0; 300 estimators, maximum depth 3, learning rate 0.05, subsample 0.9, column subsample 0.8 and seed 42), with median imputation fitted inside each training fold. Under patient-level five-fold cross-validation, we trained a separate model in each fold against the operational same-eye ESI-early target; all eyes from one patient remained in the same fold, and one eye per patient was selected within each comparison group. The resulting predictions formed a frozen out-of-fold score, and no eye from a substituted target group was used to fit, tune or select the operational models.

The frozen score was then evaluated against three target definitions ordered by their separation from the same-eye ESI label-generating pathway, each with the same fixed comparator population: operational ESI-early; FFearly, with the target eye ESI-normal and the contralateral eye ESI-early; and the FFKC proxy, with the target eye ESI-normal and the contralateral eye graded keratoconus by ESI. The ESI thresholds and group counts are given in Supplementary Table S8. Group membership was defined in two orders relative to morphology quality control. The primary specification defined the bilateral relationship before quality control and gives the Fig. 4 comparison groups (1,124 strict-control, 179 operational, 124 FFearly and 86 FFKC-proxy eyes); a QC-first specification applied quality control before defining the relationship (1,095, 179, 114 and 66 eyes) and was used for the fixed-threshold transport and target-specific refit analyses. The Target-Swap Gap was 0.447 under the primary and 0.437 under the QC-first specification. We computed and reported two estimands separately: fixed-score transport evaluated the frozen operational score on the substituted target, whereas refit retrained the full pipeline directly against the substituted target, with significance assessed against a permutation null. The Target-Swap Gap is the operational AUC minus the transport AUC, and Independent-proxy Target Retention is the transported above-chance discrimination as a fraction of the operational above-chance discrimination. For index reconstruction, we fitted an XGBoost regressor that predicted anterior ESI from the same 298 features (squared-error objective; 300 estimators, maximum depth 3, learning rate 0.05, subsample 0.9, column subsample 0.8, seed 42, fold-local median imputation). Patient-grouped five-fold out-of-fold predictions were generated across all 3,060 eyes, and the association between observed and out-of-fold-predicted anterior ESI was quantified by Pearson correlation. Label-recovery AUC was calculated for ESI-early eyes versus controls meeting the bilateral ESI-defined true-normal rule. For each feature, we fixed direction using the operational ESI-early comparison and applied it unchanged to the FFKC-proxy comparison, counted features with a positive operational-minus-proxy AUC gap with ties excluded, and summarized operational AUC minus same-direction proxy AUC by its across-feature median and a 2,000-replicate joint patient-bootstrap interval. The six feature families were descriptive groupings based solely on source column names.

### Registered fixed-threshold target transport

Two decision thresholds were selected on the operational-target versus strict-control out-of-fold scores (179 operational-target eyes versus 1,095 strict-control eyes, one eye per patient). The first maximized operational sensitivity at empirical specificity of at least 95%; the second met the corresponding 99% constraint. Each threshold was then applied unchanged to the FFearly (114 eyes) and FFKC-proxy (66 eyes) targets against the same strict controls, with the threshold reselected within every bootstrap replicate so that the intervals include threshold-selection uncertainty. Missed cases per 1,000 people at 1–10% prevalence follow the registered formula and are tabulated in the Zenodo archive, with 2,000 joint patient-cluster bootstrap intervals throughout. The estimand is fixed-threshold decision-analytic transport under the registered prevalence scenarios.

### Cross-device analysis

Cross-device analysis used eight shared features: steep K, flat K, mean K, maximum K, thinnest corneal thickness, apex corneal thickness, higher-order aberration root mean square and coma magnitude. Each model was fitted on the full training-device cohort and scored directly in the other device cohort. Logistic regression used full-training-device median imputation, standardization and class-balanced fitting; the XGBoost model used median imputation, 300 estimators, maximum depth 3, learning rate 0.05, subsample 0.9, column subsample 0.8, class weighting and seed 42. The test-set positive/negative counts were 162/2,699 for CASIA manifest keratoconus, 199/2,699 for CASIA ESI-early, 83/82 for Almería manifest keratoconus and 40/82 for Almería pre-labeled subclinical; this analysis reports point AUCs without confidence intervals.

### Registered continuous BAD-D reconstruction

BAD-D was predicted from the 40 non-composite Almería measurements by five-fold out-of-fold XGBoost regression with seed 42, fold-local median imputation, squared-error loss, 300 trees, maximum depth 3, learning rate 0.05, subsample 0.9, column subsample 0.8 and L2 regularization 1.0. Out-of-fold Pearson correlation, root mean squared error, mean absolute error and R² were computed on the 205 eyes, with 2,000-resample percentile intervals.

### Same-device increment and demographic pathway analyses

The Almería analyses used a public Pentacam cohort of 205 eyes (Mendeley Data, https://doi.org/10.17632/fcb47rngbd.1), for which no patient-level linkage identifier was available, so all analyses used stratified eye-level folds and eye-level bootstrap intervals, and no fellow-eye pairing or target substitution was attempted. This cohort supported two analyses: whether BAD-D added discrimination beyond 40 non-composite measurements from the same device, and whether age and sex alone separated the groups. Within the 122-eye normal-versus-pre-labeled-subclinical subset, each of 40 non-composite measurements was analyzed separately; Spearman correlation with BAD-D and raw-direction univariable AUC for the subclinical label used pairwise complete observations, and five descriptive feature families followed a fixed mapping of source columns provided in the Zenodo archive. The three same-device specifications were an XGBoost model using the 40 non-composite measurements, raw BAD-D alone evaluated as a univariable score on pairwise-complete observations, and an XGBoost model using the 40 measurements plus BAD-D. Both XGBoost models used 300 estimators, maximum depth 3, learning rate 0.05, subsample 0.9, column subsample 0.8, L2 regularization 1.0, training-fold class weighting and seed 42. Median imputation was fitted within each training fold, and identical stratified five-fold eye-level splits were reused. The same cross-fitted specifications were applied to the manifest-keratoconus contrast (83 manifest versus 82 control eyes). ΔAUC was the AUC of the 40-measurement-plus-BAD-D cross-fitted predictions minus that of the 40-measurement cross-fitted predictions, with its 95% CI taken from the same 2,000 paired eye-level bootstrap resamples.

For the demographic analysis, every corneal measurement was discarded, and age-plus-sex and age-only models were fitted separately against the pre-labeled subclinical and manifest keratoconus targets using the same XGBoost specification and stratified five-fold eye-level splits as the same-device 40-feature analysis, with 2,000-replicate eye bootstrap intervals. For each target, the age-plus-sex model was compared with 200 label permutations using an upper-tail add-one Monte Carlo *P* value. Group comparability was also assessed using the field’s conventional location tests, alongside dispersion diagnostics. Corpus-level counting of demographic safeguards (independence testing, demographic model inputs, reliance on location tests) followed prespecified text-based criteria given in the Zenodo archive.

### Reporting-bias and certainty assessment

Reporting and selection biases were assessed narratively, because no pooled meta-analysis was performed and funnel-plot and small-study methods did not apply. Three structural patterns were prespecified for recording: within-context AUC dispersion among fixed measurement contexts reporting two or more models or indices, the availability of extractable point AUCs for subclinical-target studies, and the structure of reported intervals, including zero-width intervals at AUC 1.000.

Certainty in each principal finding was graded with a rubric informed by the Grading of Recommendations Assessment, Development and Evaluation (GRADE) approach, as registered. The rubric scored contributing-study count and consistency, the determinable versus unclear proportion, risk of bias and extraction reproducibility, each from 0 to 2, with mandatory downgrades when unclear or mixed status exceeded 25% and 50% of contributing studies; the full grading table is provided in the Zenodo archive.

### Statistics and reproducibility

Each inferential analysis is reported separately; multiplicity adjustment was limited to the Bonferroni correction within the verification confidence limit. Confidence intervals are 95% percentile bootstrap intervals from 2,000 replicates, resampled at the patient level for CASIA analyses (patients as clusters, so that fellow eyes were never split across replicates) and at the eye level for Almería analyses (no patient identifier available). Bootstrap intervals were used for AUCs and their differences because the sampling distribution of an AUC under patient clustering has no closed form at these group sizes. Analysis sample sizes were 1,513 comparison eyes for the frozen-score target-substitution AUCs, with group counts by target in Supplementary Table S8, and 3,060 eyes for anterior ESI reconstruction. The target-stage directionality contrast used 61 studies (65 partitions), resampled at the study level; the Almería same-device, BAD-D-increment and demographic models used 40 pre-labeled subclinical versus 82 control eyes and 83 manifest versus 82 control eyes; and continuous BAD-D reconstruction used 205 eyes. Permutation *P* values are add-one Monte Carlo estimates: the refit comparison used 1,000 fold-preserving label permutations with a two-sided alternative on the statistic |AUC − 0.5|, and the demographic models used 200 label permutations with an upper-tail alternative, chosen because the hypothesis of interest was discrimination above chance. The prespecified univariate age comparisons used the two-sided Mann–Whitney U test (40 pre-labeled subclinical versus 82 control eyes, and 83 manifest versus 82 control eyes), chosen because the age distributions were skewed and differed in dispersion. The verification upper confidence limit is one-sided at the 95% level. The threshold for interpreting a *P* value was 0.05, and exact *P* values are reported. Pearson and Spearman correlations are reported as descriptive measures without hypothesis tests; ± denotes s.d. throughout. Analyses used Python 3.12.13 with NumPy 2.4.6, pandas 3.0.3, SciPy 1.18.0, scikit-learn 1.9.0 and XGBoost 3.3.0. The Zenodo archive (Data availability) provides the adjudication tables, extraction codebook, analysis scripts and unit-level assessments used for the corpus-level results.

### Use of large language models

Large language model tools were used to improve the language of the manuscript and to assist with writing analysis code. They were not used for the registered searches, screening, study selection, data extraction, adjudication or the interpretation of results. The authors reviewed, edited and verified all output and take full responsibility for the content of this article.

## Supporting information

PRISMA 2020 checklists

## Data availability

All data derive from the published literature and two public datasets: the CASIA cohort (Harvard Dataverse, CC0) and the Almería Pentacam cohort (Mendeley Data, CC BY 4.0, https://doi.org/10.17632/fcb47rngbd.1). The study roster, unit-level assessments, extraction tables, codebook, index-mention evidence and protocol deviations are archived at Zenodo under https://doi.org/10.5281/zenodo.22304145.

## Code availability

Analysis scripts for the corpus-level and experimental results are archived at Zenodo under https://doi.org/10.5281/zenodo.22304145, with software and package versions listed in Methods.

## Acknowledgements

This work was supported by the Beijing Hospitals Authority’s Ascent Plan (No. DFL20240202), the “Yicheng Outstanding Talents” program of the Beijing Economic-Technological Development Area and the National High-Level Talents Special Support Program (Leading Scientific and Technological Innovation Talent). The funders had no role in study design, data collection, analysis and interpretation, or writing of the manuscript.

## Author contributions

H.J. conceived and designed the study, performed data extraction and the analyses and wrote the manuscript. F.G. contributed to the study design, performed data extraction, interpreted the results and revised the manuscript. F.G. and L.Y. independently readjudicated the author-linked study, with disagreements resolved by Ying Jie. Yang Jiang adjudicated disagreements in the original and supplemental extraction sets. Ying Jie, L.Y., Y.L. and Yang Jiang supervised the study and revised the manuscript. All authors read and approved the final manuscript.

