## Supplementary material for "Systematic review finds keratoconus AI unproven in fellow eyes with normal topography and tomography": PRISMA 2020 checklists

### PRISMA 2020 checklist

PRISMA 2020 checklist for the manuscript 'Systematic review finds keratoconus AI unproven in fellow eyes with normal topography and tomography'. Locations refer to section headings of the main text, Supplementary Information and the Zenodo archive.

#### PRISMA 2020 checklist

| Item | Location where item is reported | Note |
|---|---|---|
| 1 — Identify the report as a systematic review. | Title | |
| 2 — See the PRISMA 2020 for Abstracts checklist. | Abstract; PRISMA 2020 for Abstracts checklist below | |
| 3 — Describe the rationale for the review in the context of existing knowledge. | Introduction | |
| 4 — Provide an explicit statement of the objective(s) or question(s) the review addresses. | Introduction | |
| 5 — Specify the inclusion and exclusion criteria for the review and how studies were grouped for the syntheses. | Methods, Search and corpus construction; Methods, Claim-by-stratum extraction and adjudication; PROSPERO CRD420261441197 | |
| 6 — Specify all databases, registers, websites, organisations, reference lists and other sources searched or consulted to identify studies. Specify the date when each source was last searched or consulted. | Methods, Search and corpus construction; Supplementary Table S1; Zenodo archive, 08_search_strategy/; PROSPERO CRD420261441197 | Registered search boundary 27 June 2026; the PubMed search has a recorded execution date (4 July 2026) and a retained list of 2,090 PMIDs, the other five source exports carry no execution date (deviation recorded in the registration) |
| 7 — Present the full search strategies for all databases, registers and websites, including any filters and limits used. | Methods, Search and corpus construction; Supplementary Table S1; Zenodo archive, 08_search_strategy/ | Executed queries are given for PubMed, Embase, Scopus and Web of Science; for Europe PMC and OpenAlex the query components are given, as the exact executed strings were not retained |
| 8 — Specify the methods used to decide whether a study met the inclusion criteria of the review, including how many reviewers screened each record and each report retrieved, whether they worked independently, and if applicable, details of automation tools used in the process. | Methods, Search and corpus construction; Zenodo archive, 09_registration_and_deviations/; PROSPERO CRD420261441197 | |
| 9 — Specify the methods used to collect data from reports, including how many reviewers collected data from each report, whether they worked independently, any processes for obtaining or confirming data from study investigators, and if applicable, details of automation tools used in the process. | Methods, Claim-by-stratum extraction and adjudication; Methods, Verification subsample for non-adjudicated studies; Zenodo archive, 02_claim_adjudication/; PROSPERO CRD420261441197 | |
| 10a — List and define all outcomes for which data were sought. Specify whether all results that were compatible with each outcome domain in each study were sought (e.g. for all measures, time points, analyses), and if not, the methods used to decide which results to collect. | Methods, Claim-by-stratum extraction and adjudication; Methods, Target-stage directionality comparison; Methods, Registered natural target-swap adjudication; Methods, Partial identification across target strata; PROSPERO CRD420261441197 | |
| 10b — List and define all other variables for which data were sought (e.g. participant and intervention characteristics, funding sources). Describe any assumptions made about any missing or unclear information. | Methods, Claim-by-stratum extraction and adjudication; Zenodo archive, 01_corpus_roster/ and 02_claim_adjudication/; PROSPERO CRD420261441197 | |
| 11 — Specify the methods used to assess risk of bias in the included studies, including details of the tool(s) used, how many reviewers assessed each study and whether they worked independently, and if applicable, details of automation tools used in the process. | Zenodo archive, 01_corpus_roster/ and 10_prisma/; PROSPERO CRD420261441197 | |

| Item | Location where item is reported | Note |
|---|---|---|
| 12 — Specify for each outcome the effect measure(s) (e.g. risk ratio, mean difference) used in the synthesis or presentation of results. | Methods, Target-stage directionality comparison; Methods, Partial identification across target strata; Methods, Controlled target-substitution experiment; Methods, Registered fixed-threshold target transport; Methods, Statistics and reproducibility | |
| 13a — Describe the processes used to decide which studies were eligible for each synthesis (e.g. tabulating the study intervention characteristics and comparing against the planned groups for each synthesis (item #5)). | Methods, Target-stage directionality comparison; Methods, Registered natural target-swap adjudication; Methods, Partial identification across target strata | |
| 13b — Describe any methods required to prepare the data for presentation or synthesis, such as handling of missing summary statistics, or data conversions. | Methods, Target-stage directionality comparison; Methods, Partial identification across target strata; Methods, Controlled target-substitution experiment; Methods, Cross-device analysis; Methods, Same-device increment and demographic pathway analyses | |
| 13c — Describe any methods used to tabulate or visually display results of individual studies and syntheses. | Results; Figs. 1–5; Supplementary Figs. 1–3; Supplementary Tables S1–S10 | |
| 13d — Describe any methods used to synthesize results and provide a rationale for the choice(s). If meta-analysis was performed, describe the model(s), method(s) to identify the presence and extent of statistical heterogeneity, and software package(s) used. | Methods, Target-stage directionality comparison; Methods, Registered natural target-swap adjudication; Methods, Partial identification across target strata; Methods, Reporting-bias and certainty assessment; Methods, Statistics and reproducibility | Meta-analysis-specific elements are not applicable; non-meta-analytic synthesis methods and results are reported at the listed locations |
| 13e — Describe any methods used to explore possible causes of heterogeneity among study results (e.g. subgroup analysis, meta-regression). | Methods, Target-stage directionality comparison; Methods, Partial identification across target strata; Methods, Cross-device analysis; Methods, Same-device increment and demographic pathway analyses | |
| 13f — Describe any sensitivity analyses conducted to assess robustness of the synthesized results. | Methods, Registered natural target-swap adjudication; Methods, Partial identification across target strata; Supplementary Tables S7 and S8; PROSPERO CRD420261441197 | |
| 14 — Describe any methods used to assess risk of bias due to missing results in a synthesis (arising from reporting biases). | Methods, Reporting-bias and certainty assessment; Zenodo archive, 10_prisma/ | |
| 15 — Describe any methods used to assess certainty (or confidence) in the body of evidence for an outcome. | Methods, Reporting-bias and certainty assessment; Zenodo archive, 10_prisma/; PROSPERO CRD420261441197 | |
| 16a — Describe the results of the search and selection process, from the number of records identified in the search to the number of studies included in the review, ideally using a flow diagram. | Results, VAE-NTT discrimination was not established in the corpus; Supplementary Table S1; Supplementary Fig. 2 | |
| 16b — Cite studies that might appear to meet the inclusion criteria, but which were excluded, and explain why they were excluded. | Zenodo archive, 10_prisma/ | |
| 17 — Cite each included study and present its characteristics. | Results, VAE-NTT discrimination was not established in the corpus; Supplementary Table S2; Zenodo archive, 01_corpus_roster/ | |
| 18 — Present assessments of risk of bias for each included study. | Zenodo archive, 01_corpus_roster/ and 10_prisma/ | |
| 19 — For all outcomes, present, for each study: (a) summary statistics for each group (where appropriate) and (b) an effect estimate and its precision (e.g. confidence/credible interval), ideally using structured tables or plots. | Results; Supplementary Tables S3–S9; Zenodo archive, 01_corpus_roster/, 02_claim_adjudication/, 03_index_provenance/ and 04_partial_identification/ | |

| Item | Location where item is reported | Note |
| --- | --- | --- |
| 20a — For each synthesis, briefly summarise the characteristics and risk of bias among contributing studies. | Results; Supplementary Tables S2, S3 and S5–S7; Zenodo archive, 01_corpus_roster/, 02_claim_adjudication/, 04_partial_identification/ and 10_prisma/ | |
| 20b — Present results of all statistical syntheses conducted. If meta-analysis was done, present for each the summary estimate and its precision (e.g. confidence/credible interval) and measures of statistical heterogeneity. If comparing groups, describe the direction of the effect. | Results; Supplementary Tables S3, S4 and S7–S9 | Meta-analysis-specific elements are not applicable; non-meta-analytic synthesis methods and results are reported at the listed locations |
| 20c — Present results of all investigations of possible causes of heterogeneity among study results. | Results, Broad-stratum AUC did not identify VAE-NTT performance; Results, A fixed score lost discrimination as the target moved away from the label-generating pathway; Results, Positive controls preserved manifest-disease performance, while demographics provided an alternative route to discrimination; Supplementary Tables S7–S9 | |
| 20d — Present results of all sensitivity analyses conducted to assess the robustness of the synthesized results. | Results, A fixed score lost discrimination as the target moved away from the label-generating pathway; Results, Broad-stratum AUC did not identify VAE-NTT performance; Supplementary Tables S7 and S8; PROSPERO CRD420261441197 | |
| 21 — Present assessments of risk of bias due to missing results (arising from reporting biases) for each synthesis assessed. | Results, Reported AUC varied within fixed contexts and carried its narrowest intervals where it was highest; Zenodo archive, 10_prisma/ | |
| 22 — Present assessments of certainty (or confidence) in the body of evidence for each outcome assessed. | Results, Reported AUC varied within fixed contexts and carried its narrowest intervals where it was highest; Zenodo archive, 10_prisma/ | |
| 23a — Provide a general interpretation of the results in the context of other evidence. | Discussion | |
| 23b — Discuss any limitations of the evidence included in the review. | Discussion | |
| 23c — Discuss any limitations of the review processes used. | Discussion; Methods, Study design and registration; Zenodo archive, 09_registration_and_deviations/; PROSPERO CRD420261441197 | |
| 23d — Discuss implications of the results for practice, policy, and future research. | Discussion | |
| 24a — Provide registration information for the review, including register name and registration number, or state that the review was not registered. | Methods, Study design and registration; PROSPERO CRD420261441197 | |
| 24b — Indicate where the review protocol can be accessed, or state that a protocol was not prepared. | Methods, Study design and registration; Zenodo archive, 09_registration_and_deviations/; PROSPERO CRD420261441197 | |
| 24c — Describe and explain any amendments to information provided at registration or in the protocol. | Methods, Study design and registration; Zenodo archive, 09_registration_and_deviations/; PROSPERO CRD420261441197 | |
| 25 — Describe sources of financial or non-financial support for the review, and the role of the funders or sponsors in the review. | Acknowledgements | |
| 26 — Declare any competing interests of review authors. | Competing interests | |

| Item | Location where item is reported | Note |
|---|---|---|
| 27 — Report which of the following are publicly available and where they can be found: template data collection forms; data extracted from included studies; data used for all analyses; analytic code; any other materials used in the review. | Data availability; Code availability; Methods, Statistics and reproducibility; Zenodo archive, 01_corpus_roster/, 02_claim_adjudication/, 03_index_provenance/, 04_partial_identification/, 05_target_substitution_experiment/, 06_cross_device_and_demographic/, 07_verification_subsample/, 08_search_strategy/, 09_registration_and_deviations/, 10_prisma/ and 11_supplementary_tables/ | |

#### PRISMA 2020 for Abstracts checklist

| Item | Location where item is reported | Note |
|---|---|---|
| 1 — Identify the report as a systematic review. | Title | |
| 2 — Provide an explicit statement of the main objective(s) or question(s) the review addresses. | | Not reported in the abstract |
| 3 — Specify the inclusion and exclusion criteria for the review. | | Not reported in the abstract |
| 4 — Specify the information sources (e.g. databases, registers) used to identify studies and the date when each was last searched. | | Not reported in the abstract |
| 5 — Specify the methods used to assess risk of bias in the included studies. | | Not reported in the abstract |
| 6 — Specify the methods used to present and synthesise results. | Abstract | |
| 7 — Give the total number of included studies and participants and summarise relevant characteristics of studies. | Abstract | |
| 8 — Present results for main outcomes, preferably indicating the number of included studies and participants for each. If meta-analysis was done, report the summary estimate and confidence/credible interval. If comparing groups, indicate the direction of the effect (i.e. which group is favoured). | Abstract | Meta-analysis-specific elements are not applicable; non-meta-analytic synthesis methods and results are reported at the listed locations |
| 9 — Provide a brief summary of the limitations of the evidence included in the review (e.g. study risk of bias, inconsistency and imprecision). | Abstract | |
| 10 — Provide a general interpretation of the results and important implications. | Abstract | |
| 11 — Specify the primary source of funding for the review. | | Not reported in the abstract |
| 12 — Provide the register name and registration number. | Abstract | |
