## Supplementary Information for "Systematic review finds keratoconus AI unproven in fellow eyes with normal topography and tomography"

Contents: Supplementary Figs. 1–3 and Supplementary Tables S1–S10.

Abbreviations used in the Supplementary Figures and Tables: AI, artificial intelligence; ANN, artificial neural network; ARTmax, Ambrósio Relational Thickness maximum; AUC, area under the receiver operating characteristic curve; BAD-D, Belin/Ambrósio Enhanced Ectasia Display deviation index; CART, classification and regression tree; CBI, Corvis Biomechanical Index; CH, corneal hysteresis; CHAID, chi-squared automatic interaction detection; CI, confidence interval; CRF, corneal resistance factor; CV, cross-validation; DOI, digital object identifier; ESI, Ectasia Screening Index; FF, forme fruste; FFKC, forme fruste keratoconus; FT, corpus study identifier; HOA, higher-order aberrations; I-S, inferior –superior asymmetry value; IHD, index of height decentration; KA, Keratoconus Assistant; KC, keratoconus; KCI, keratoconus index; KISA%, keratoconus percentage index; Kmax, maximum keratometry; KSI, keratoconus severity index; LASIK, laser-assisted in situ keratomileusis; OCT, optical coherence tomography; ORA, Ocular Response Analyzer; PRFI, Pentacam Random Forest Index; RF, random forest; RMS, root mean square; ROC, receiver operating characteristic; s.d., standard deviation; SD-OCT, spectral-domain optical coherence tomography; SS, combined forme fruste and suspect keratoconus group; TBI, Tomographic and Biomechanical Index; TBIv2, second version of the Tomographic and Biomechanical Index; UZG, Ghent University Hospital; VAE-NT, very asymmetric ectasia with normal topography; VAE-NTT, very asymmetric ectasia with normal topography and tomography.

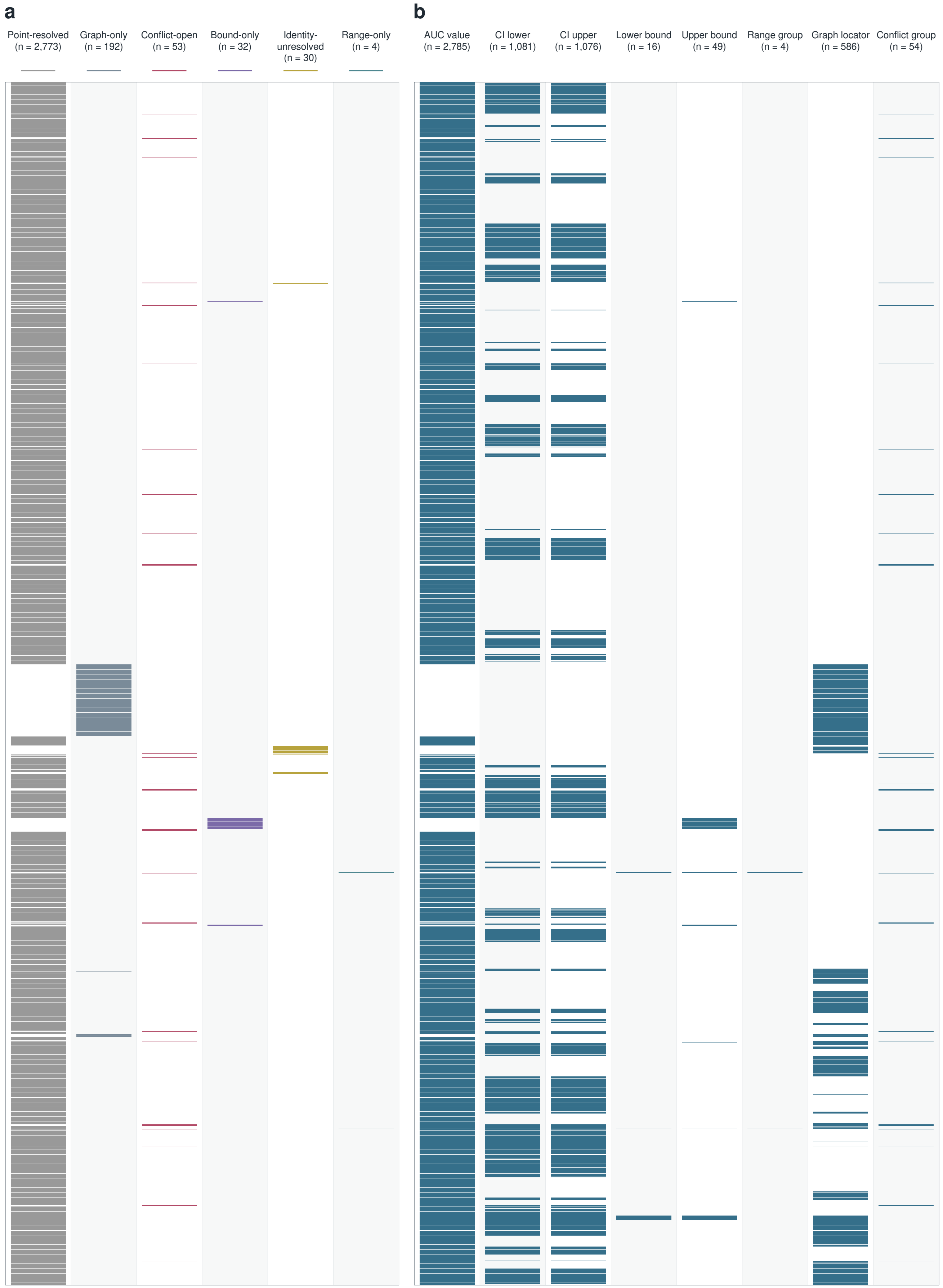
a
Point-resolved (n = 2,773)
Graph-only (n = 192)
Conflict-open (n = 53)
Bound-only (n = 32)
Identity-unresolved (n = 30)
Range-only (n = 4)
b
AUC value (n = 2,785)
CI lower (n = 1,081)
CI upper (n = 1,076)
Lower bound (n = 16)
Upper bound (n = 49)
Range group (n = 4)
Graph locator (n = 586)
Conflict group (n = 54)

**Supplementary Fig. 1 | Resolution states and field completeness of all 3,084 AUC records.** **a**, Each of the 3,084 AUC records extracted from 157 studies receives one mark in exactly one of six resolution states: point-resolved (n = 2,773), graph-only (n = 192), conflict-open (n = 53), bound-only (n = 32), identity-unresolved (n = 30) and range-only (n = 4). Rows are ordered by study, extraction set (original or supplementary), partition and resolution state, and the row order is identical in both panels. **b**, Presence of the eight source fields for the same 3,084 records (8 × 3,084 = 24,672 cells; a filled cell denotes a non-empty field; 5,651 cells are non-empty). Column headers give per-field counts. Resolution states describe how each AUC value is documented in its source report.

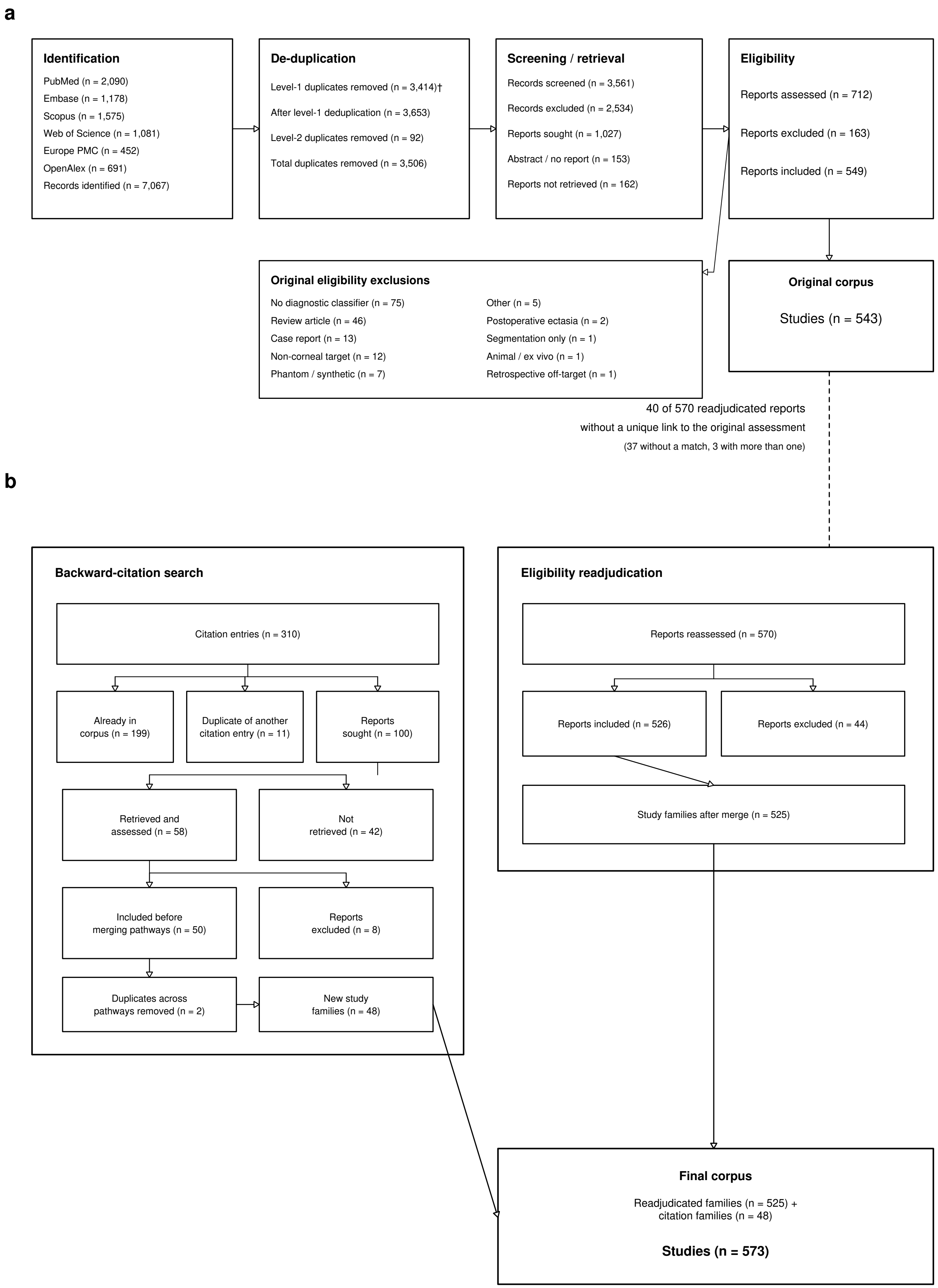
a
Identification
PubMed (n = 2,090)
Embase (n = 1,178)
Scopus (n = 1,575)
Web of Science (n = 1,081)
Europe PMC (n = 452)
OpenAlex (n = 691)
Records identified (n = 7,067)
De-duplication
Level-1 duplicates removed (n = 3,414)†
After level-1 deduplication (n = 3,653)
Level-2 duplicates removed (n = 92)
Total duplicates removed (n = 3,506)
Screening / retrieval
Records screened (n = 3,561)
Records excluded (n = 2,534)
Reports sought (n = 1,027)
Abstract / no report (n = 153)
Reports not retrieved (n = 162)
Eligibility
Reports assessed (n = 712)
Reports excluded (n = 163)
Reports included (n = 549)
Original eligibility exclusions
No diagnostic classifier (n = 75)
Review article (n = 46)
Case report (n = 13)
Non-corneal target (n = 12)
Phantom / synthetic (n = 7)
Other (n = 5)
Postoperative ectasia (n = 2)
Segmentation only (n = 1)
Animal / ex vivo (n = 1)
Retrospective off-target (n = 1)
Original corpus
Studies (n = 543)
40 of 570 readjudicated reports
without a unique link to the original assessment
(37 without a match, 3 with more than one)
b
Backward-citation search
Citation entries (n = 310)
Already in corpus (n = 199)
Duplicate of another citation entry (n = 11)
Reports sought (n = 100)
Retrieved and assessed (n = 58)
Not retrieved (n = 42)
Included before merging pathways (n = 50)
Reports excluded (n = 8)
Duplicates across pathways removed (n = 2)
New study families (n = 48)
Eligibility readjudication
Reports reassessed (n = 570)
Reports included (n = 526)
Reports excluded (n = 44)
Study families after merge (n = 525)
Final corpus
Readjudicated families (n = 525) + citation families (n = 48)
Studies (n = 573)

**Supplementary Fig. 2 | Original search and eligibility readjudication leading to the 573-study corpus. a**, The original six-database search: 7,067 records identified; 3,561 screened; 1,027 reports sought; 712 assessed; 549 reports included; 543 studies. The dagger marks level-1 deduplication, performed under a rule that was later revised (Supplementary Table S1, note 1). **b**, The final corpus combines two pathways that remain separate until study families are merged: eligibility readjudication of the original reports (570 reports reassessed; 526 included; 525 study families) and the backward-citation search (310 citation entries; 100 reports sought; 58 assessed; 50 included; 48 new study families after removal of 2 duplicates across pathways), giving 525 + 48 = 573 studies. The dashed connection between panels marks that 40 of the 570 readjudicated reports lack a unique link to the original assessment (37 without a match, 3 with more than one).

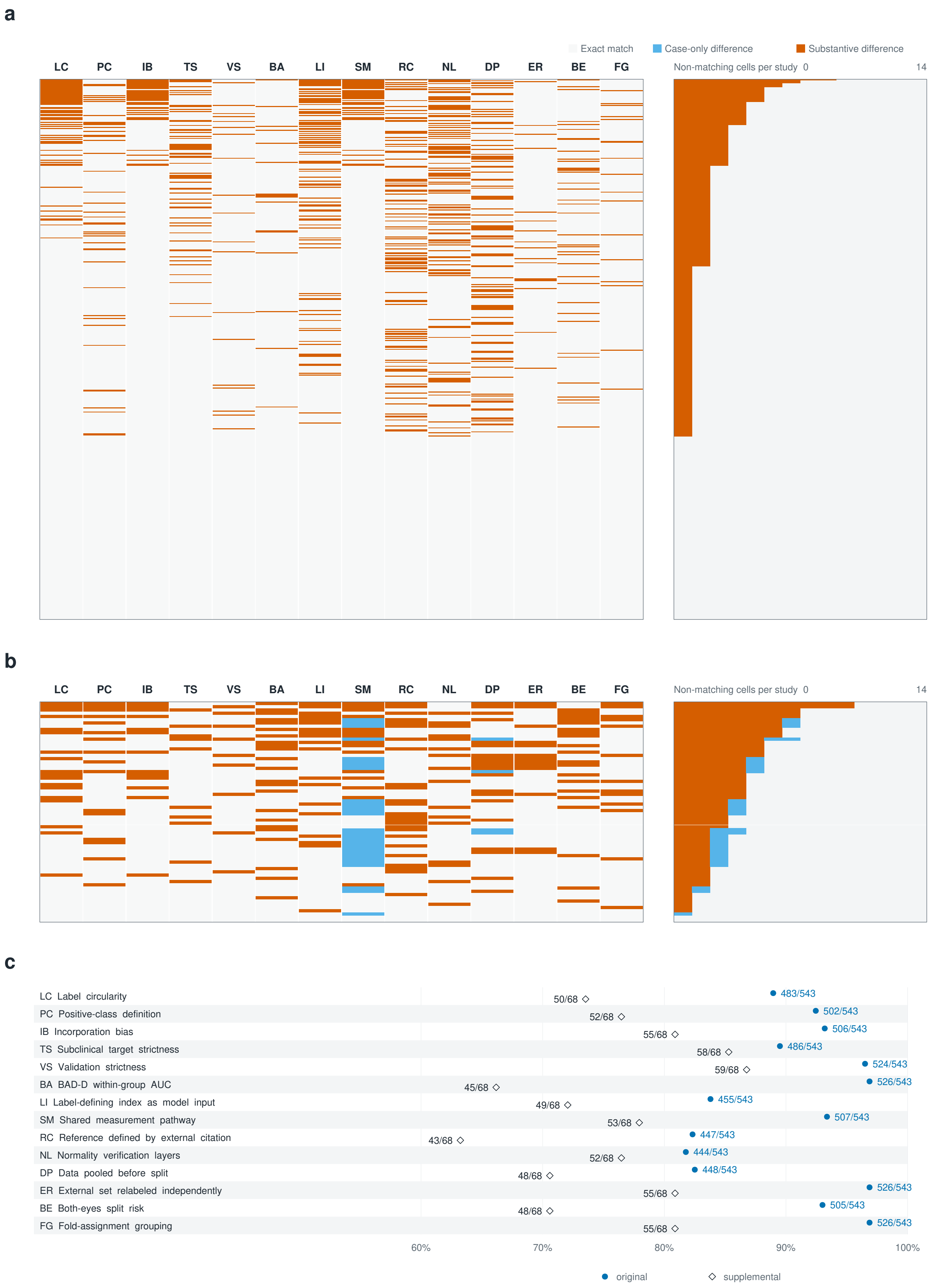
a
Exact match
Case-only difference
Substantive difference
LC PC IB TS VS BA LI SM RC NL DP ER BE FG
Non-matching cells per study 0 14
b
LC PC IB TS VS BA LI SM RC NL DP ER BE FG
Non-matching cells per study 0 14
c
LC Label circularity 50/68 483/543
PC Positive-class definition 52/68 502/543
IB Incorporation bias 55/68 506/543
TS Subclinical target strictness 58/68 486/543
VS Validation strictness 59/68 524/543
BA BAD-D within-group AUC 45/68 526/543
LI Label-defining index as model input 49/68 455/543
SM Shared measurement pathway 53/68 507/543
RC Reference defined by external citation 43/68 447/543
NL Normality verification layers 52/68 444/543
DP Data pooled before split 48/68 448/543
ER External set relabeled independently 55/68 526/543
BE Both-eyes split risk 48/68 505/543
FG Fold-assignment grouping 55/68 526/543
60%
70%
80%
90%
100%
original
supplemental

**Supplementary Fig. 3 | Cell-level agreement of independent extraction across the original and supplementary extraction sets. a**, Original set: all 7,602 paired cells from 543 double-coded studies × 14 CORE fields; normalized agreement 6,885/7,602 (90.6%). Column codes give the 14 CORE fields: LC, label circularity; PC, positive-class definition; IB, incorporation bias; TS, subclinical target strictness; VS, validation strictness; BA, BAD-D within-group AUC; LI, label-defining index as model input; SM, shared measurement pathway; RC, reference defined by external citation; NL, normality verification layers; DP, data pooled before split; ER, external set relabeled independently; BE, both-eyes split risk; FG, fold-assignment grouping. **b**, Supplementary set: all 952 paired cells from 68 studies × 14 CORE fields; exact agreement 690/952 (72.5%), normalized agreement 722/952 (75.8%). Cells are exact matches, case-only differences (resolved by lowercasing after whitespace trimming; all 32 occur in the supplementary set) or substantive differences. Rows are sorted by substantive-mismatch burden and the right-hand bars give per-study nonexact cell counts. **c**, Field-level normalized agreement per set; each label gives agreeing cells over the set denominator. Agreement is pre-adjudication coder concordance. The original-set denominator is all 543 studies double-coded at extraction.

### Supplementary Table S1. Identification, screening, retrieval, and eligibility accounting

This table accounts for the original six-database search, the subsequent eligibility readjudication and backward-citation search, and the resulting 573-study corpus.

| Phase | Path | Item | Count | Unit | Note |
|---|---|---|---|---|---|
| Identification | Original six-database search | PubMed | 2,090 | records | |
| Identification | Original six-database search | Embase | 1,178 | records | |
| Identification | Original six-database search | Scopus | 1,575 | records | |
| Identification | Original six-database search | Web of Science | 1,081 | records | |
| Identification | Original six-database search | Europe PMC | 452 | records | |
| Identification | Original six-database search | OpenAlex | 691 | records | |
| Identification | Original six-database search | Records identified | 7,067 | records | |
| Deduplication | Original six-database search | Level-1 duplicates removed | 3,414 | records | Deduplication rule revised afterwards (note 1) |
| Deduplication | Original six-database search | Records after level-1 deduplication | 3,653 | records | |
| Deduplication | Original six-database search | Level-2 duplicates removed | 92 | records | |
| Deduplication | Original six-database search | Total duplicates removed | 3,506 | records | |
| Screening | Original six-database search | Records screened | 3,561 | records | |
| Screening | Original six-database search | Records excluded | 2,534 | records | |
| Retrieval | Original six-database search | Reports sought | 1,027 | reports | |
| Retrieval | Original six-database search | Conference abstract or no full report | 153 | reports | |
| Retrieval | Original six-database search | Reports not retrieved | 162 | reports | |
| Eligibility | Original six-database search | Reports assessed | 712 | reports | |
| Eligibility | Original six-database search | Reports excluded | 163 | reports | |
| Exclusion reason | Original eligibility assessment | No diagnostic classifier | 75 | reports | |
| Exclusion reason | Original eligibility assessment | Review article | 46 | reports | |
| Exclusion reason | Original eligibility assessment | Case report | 13 | reports | |
| Exclusion reason | Original eligibility assessment | Non-corneal target | 12 | reports | |
| Exclusion reason | Original eligibility assessment | Phantom or synthetic study | 7 | reports | |
| Exclusion reason | Original eligibility assessment | Other | 5 | reports | |

| Phase | Path | Item | Count | Unit | Note |
|---|---|---|---|---|---|
| Exclusion reason | Original eligibility assessment | Postoperative ectasia | 2 | reports | |
| Exclusion reason | Original eligibility assessment | Segmentation only | 1 | reports | |
| Exclusion reason | Original eligibility assessment | Animal or ex vivo | 1 | reports | |
| Exclusion reason | Original eligibility assessment | Retrospective off-target | 1 | reports | |
| Inclusion | Original six-database search | Reports included | 549 | reports | |
| Inclusion | Original six-database search | Integrated analytic studies | 543 | studies | Superseded by the 573-study corpus |
| Readjudication | Eligibility readjudication of the original reports | Reports reassessed | 570 | reports | |
| Readjudication | Eligibility readjudication of the original reports | Reports excluded | 44 | reports | |
| Readjudication | Eligibility readjudication of the original reports | Reports included | 526 | reports | |
| Readjudication | Eligibility readjudication of the original reports | Study families after report-family merge | 525 | studies | |
| Backward citation | Backward-citation search | Backward-citation target entries | 310 | entries | |
| Backward citation | Backward-citation search | Already in corpus | 199 | entries | |
| Backward citation | Backward-citation search | Duplicate of another backward-citation entry | 11 | entries | |
| Backward citation | Backward-citation search | Reports sought | 100 | reports | |
| Backward citation | Backward-citation search | Reports retrieved and assessed | 58 | reports | |
| Backward citation | Backward-citation search | Reports not retrieved | 42 | reports | |
| Backward citation | Backward-citation search | Reports excluded | 8 | reports | |
| Backward citation | Backward-citation search | Reports included before cross-stream merge | 50 | reports | |
| Backward citation | Backward-citation search | Cross-stream duplicate reports removed | 2 | reports | |
| Backward citation | Backward-citation search | Net-new study families | 48 | studies | |
| Final corpus | Eligibility readjudication and backward-citation search | Final corpus | 573 | studies | |
| Record linkage | Eligibility readjudication of the original reports | Readjudicated reports without a unique link to the original 712-report assessment | 40 | reports | 37 reports without a matching record and 3 with more than one |
| Exclusion reasons | Eligibility readjudication and backward-citation search | Exclusion reasons after readjudication | Recorded separately for each pathway | reports | |
| Search boundary | Original six-database search | Incremental search after 27 June 2026 | Not run | records | Records published after the registered search boundary are not represented |

1. Counts from the original six-database execution are reported as executed. The level-1 deduplication rule was revised afterwards, and these counts were not recomputed.
2. The final corpus comprises 525 readjudicated study families and 48 study families added by the backward-citation search, 573 studies in total.

Machine-readable companion: table_s1.csv in the Zenodo archive (11_supplementary_tables/; 48 data rows, same headers).

### Supplementary Table S2. Characteristics of the 573 included studies

This table summarizes the fixed 573-study denominator by publication period, study design, target layer, validation approach, index role, model category, and demographic safeguards.

| Dimension | Category | n | Denominator | Percent | Missing or unclear (n) |
|---|---|---|---|---|---|
| Publication period | 1990–1999 | 14 | 573 | 2.4 | 1 |
| Publication period | 2000–2009 | 30 | 573 | 5.2 | 1 |
| Publication period | 2010–2019 | 193 | 573 | 33.7 | 1 |
| Publication period | 2020–2026 | 335 | 573 | 58.5 | 1 |
| Publication period | Missing/unreported | 1 | 573 | 0.2 | 1 |
| Study design | Case–control, multi-arm | 307 | 573 | 53.6 | 7 |
| Study design | Case–control, two-arm | 205 | 573 | 35.8 | 7 |
| Study design | Consecutive clinic spectrum | 30 | 573 | 5.2 | 7 |
| Study design | Population screening | 13 | 573 | 2.3 | 7 |
| Study design | Enriched referral | 11 | 573 | 1.9 | 7 |
| Study design | Unclear | 7 | 573 | 1.2 | 7 |
| Target layer | Established keratoconus | 199 | 573 | 34.7 | 1 |
| Target layer | Subclinical, early or forme fruste | 99 | 573 | 17.3 | 1 |
| Target layer | Mixed | 274 | 573 | 47.8 | 1 |
| Target layer | Unclear | 1 | 573 | 0.2 | 1 |
| Validation approach | True external | 54 | 573 | 9.4 | 5 |
| Validation approach | Pseudo-external | 4 | 573 | 0.7 | 5 |
| Validation approach | Internal hold-out | 106 | 573 | 18.5 | 5 |
| Validation approach | k-fold cross-validation | 65 | 573 | 11.3 | 5 |
| Validation approach | Leave-one-out cross-validation | 10 | 573 | 1.7 | 5 |
| Validation approach | None | 329 | 573 | 57.4 | 5 |
| Validation approach | Unclear | 5 | 573 | 0.9 | 5 |
| Model category | Named index | 254 | 573 | 44.3 | 0 |
| Model category | Classical machine learning | 173 | 573 | 30.2 | 0 |
| Model category | Deep learning | 92 | 573 | 16.1 | 0 |
| Model category | Hybrid | 1 | 573 | 0.2 | 0 |
| Model category | Other | 52 | 573 | 9.1 | 0 |
| Model category | Single diagnostic parameter | 1 | 573 | 0.2 | 0 |

| Dimension | Category | n | Denominator | Percent | Missing or unclear (n) |
|---|---|---|---|---|---|
| Demographic variables in model inputs | Yes | 31 | 573 | 5.4 | 86 |
| Demographic variables in model inputs | No | 456 | 573 | 79.6 | 86 |
| Demographic variables in model inputs | Unclear | 86 | 573 | 15.0 | 86 |
| Demographic location/balance assessment | Mean or proportion test | 124 | 573 | 21.6 | 85 |
| Demographic location/balance assessment | Matched design | 66 | 573 | 11.5 | 85 |
| Demographic location/balance assessment | Imbalance acknowledged | 65 | 573 | 11.3 | 85 |
| Demographic location/balance assessment | No claim | 233 | 573 | 40.7 | 85 |
| Demographic location/balance assessment | Unclear | 85 | 573 | 14.8 | 85 |
| Index role | Qualifying index-role study | 217 | 573 | 37.9 | 0 |
| Index role | Index appearance only | 14 | 573 | 2.4 | 0 |
| Index role | No recorded index appearance | 342 | 573 | 59.7 | 0 |

1. Each dimension is exhaustive over the same 573-study corpus; unclear or unreported entries remain explicit categories.
2. Index role follows the study-by-index incidence matrix: 217 qualifying studies, 14 appearance-only studies, and 342 studies with no recorded appearance.
3. Percentages are calculated as n/573 and may not sum to exactly 100.0 because of rounding.
4. Model category classifies each study's primary model family. The 212 studies described in the main text as evaluating fixed, named indices against established diagnoses are counted on that evaluation criterion and are not the same set as the 254 named-index studies.

Machine-readable companion: table_s2.csv in the Zenodo archive (11_supplementary_tables/; 39 data rows, same headers).

### Supplementary Table S3. Six-gate accounting for 2,035 claim units

This table gives the entered, exited, reason-specific, and final-verdict accounting for all 2,035 claim units evaluated through the six-gate framework. The six units with unresolved claim–stratum pairing are removed before Gate 0 in this table (2,029 entered), whereas Fig. 1a and the main text carry them through Gates 0 and 1 (2,035 entered; 1,763 reached) and set them aside before Gate 2; the exit counts are identical under both conventions, and the entered values differ by six.

| Stage | Gate | Entered (n) | Exit reason | Exited (n) | Final verdict | Remaining after stage (n) |
|---|---|---|---|---|---|---|
| Pre-gate resolution | Pairing resolution | 2,035 | Pairing disputed | 6 | Pairing disputed | 2,029 |
| Gate 0 | Existence | 2,029 | Cause mismatch | 247 | Out of scope | 1,757 |
| Gate 0 | Existence | 2,029 | No positive claim | 1 | Out of scope | 1,757 |
| Gate 0 | Existence | 2,029 | Reporting indeterminate | 24 | Indeterminate | 1,757 |
| Gate 1 | Population normality | 1,757 | Claim overreach | 97 | Adverse | 197 |
| Gate 1 | Population normality | 1,757 | Evidence incomplete | 5 | Adverse | 197 |
| Gate 1 | Population normality | 1,757 | Not evidence for VAE-NTT | 1,292 | Out of scope | 197 |
| Gate 1 | Population normality | 1,757 | Reporting indeterminate | 166 | Indeterminate | 197 |
| Gate 2 | Identity truth | 197 | Evidence incomplete | 133 | Adverse | 55 |
| Gate 2 | Identity truth | 197 | Positive-class index circularity | 4 | Adverse | 55 |
| Gate 2 | Identity truth | 197 | Reporting indeterminate | 5 | Indeterminate | 55 |
| Gate 3 | Headline input | 55 | Cross-partition observation leakage | 1 | Adverse | 3 |
| Gate 3 | Headline input | 55 | Evidence incomplete | 3 | Adverse | 3 |
| Gate 3 | Headline input | 55 | Reporting indeterminate | 38 | Indeterminate | 3 |
| Gate 3 | Headline input | 55 | Target–input isolation failure | 5 | Adverse | 3 |
| Gate 3 | Headline input | 55 | Threshold-asymmetry answer leakage | 5 | Adverse | 3 |
| Gate 4 | Evidence strength | 3 | Evidence incomplete | 2 | Adverse | 0 |
| Gate 4 | Evidence strength | 3 | Reporting indeterminate | 1 | Indeterminate | 0 |
| Gate 5 | Claim alignment | 0 | No units reached or exited | 0 | NA | 0 |
| Final verdict summary | NA | 2,035 | NA | 0 | Supported | NA |
| Final verdict summary | NA | 2,035 | NA | 1,540 | Out of scope | NA |
| Final verdict summary | NA | 2,035 | NA | 234 | Indeterminate | NA |
| Final verdict summary | NA | 2,035 | NA | 255 | Adverse | NA |
| Final verdict summary | NA | 2,035 | NA | 6 | Pairing disputed | NA |
| Ancillary normality audit | Population normality | 487 | No normality judgment record | 75 | Not a gate exit | 412 |
| Ancillary verification audit | NA | 394 | Studies blind-recoded in verification sample | 122 | 0 recoded as VAE-NTT | 272 |

1. Gate 0 and Gate 1 account for 1,832 exits; 197 units entered Gate 2 after the six units with unresolved claim–stratum pairing were set aside outside the gate chain.

2. The final verdict distribution is 1,540 out of scope, 234 indeterminate, 255 adverse, six pairing disputed (shown as "unresolved" in Fig. 1), and zero supported. The related 487/75 normality audit and 394/122 verification audit (sampling frame at the time of sampling; Supplementary Table S4) are shown as ancillary rows, not gate exits.
3. Repeated entered and remaining values identify the common gate denominator for each reason-specific row and must not be summed across reasons.

Machine-readable companion: table_s3.csv in the Zenodo archive (11_supplementary_tables/; 26 data rows, same headers).

### Supplementary Table S4. Finite-population verification parameters and joint upper limits

This table reports the composition of the final 361-study non-adjudicated pool and the finite-population parameters behind the 24-study main limit, the 22-study Clopper–Pearson sensitivity limit, and the earlier 14-study rule-of-three calculation. Uncoded denotes studies with no previously coded target stratum.

| Panel | Method | Stratum | Population (N) | Sample (n) | Probability denominator (n) | Existing nonrandom negatives (n) | Observed VAE-NTT (k) | Unchecked (N) | Per-stratum limit or projection | Joint rounded limit | Interpretation |
|---|---|---|---|---|---|---|---|---|---|---|---|
| Final 361-study non-adjudicated pool | Composition only | Uncoded | 189 | 51 | 51 | NA | 0 | 138 | Not calculated | NA | Pool composition; no limit calculated |
| Final 361-study non-adjudicated pool | Composition only | VAE-NT | 146 | 33 | 33 | NA | 0 | 113 | Not calculated | NA | Pool composition; no limit calculated |
| Final 361-study non-adjudicated pool | Composition only | Unclear | 26 | 26 | 26 | NA | 0 | 0 | Not calculated | NA | Pool composition; no limit calculated |
| Final 361-study non-adjudicated pool | Composition only | Joint | 361 | 110 | 110 | NA | 0 | 251 | NA | Not calculated | Not a substitute for the 24-study main result |
| Final verification analysis | A: finite-population hypergeometric (main) | Uncoded | 209 | 60 | 60 | 0 | 0 | 149 | 10 | NA | Primary per-stratum limit |
| Final verification analysis | A: finite-population hypergeometric (main) | VAE-NT | 158 | 35 | 35 | 25 | 0 | 123 | 14 | NA | Primary per-stratum limit |
| Final verification analysis | A: finite-population hypergeometric (main) | Unclear | 27 | 27 | 27 | 5 | 0 | 0 | 0 | NA | Primary per-stratum limit |
| Final verification analysis | A: finite-population hypergeometric (main) | Joint | 394 | 122 | 122 | 30 | 0 | 272 | NA | 24 | Primary result |
| Final verification analysis | C: Clopper–Pearson (sensitivity) | Uncoded | 209 | 60 | 60 | 0 | 0 | 149 | 8.88 | NA | Sensitivity per-stratum limit |
| Final verification analysis | C: Clopper–Pearson (sensitivity) | VAE-NT | 158 | 35 | 35 | 25 | 0 | 123 | 12.30 | NA | Sensitivity per-stratum limit |
| Final verification analysis | C: Clopper–Pearson (sensitivity) | Unclear | 27 | 27 | 27 | 5 | 0 | 0 | 0.00 | NA | Sensitivity per-stratum limit |
| Final verification analysis | C: Clopper–Pearson (sensitivity) | Joint | 394 | 122 | 122 | 30 | 0 | 272 | 21.19 | 22 | Sensitivity result |
| Final verification analysis | B: rule-of-three (earlier calculation) | Uncoded | 209 | 60 | 60 | 0 | 0 | 149 | 7.45 | NA | Comparator only; joint coverage is below 95% |
| Final verification analysis | B: rule-of-three (earlier calculation) | VAE-NT | 183 | 35 | 60 | 25 | 0 | 123 | 6.15 | NA | Comparator only; joint coverage is below 95% |
| Final verification analysis | B: rule-of-three (earlier calculation) | Unclear | 32 | 27 | 32 | 5 | 0 | 0 | 0.00 | NA | Comparator only; joint coverage is below 95% |
| Final verification analysis | B: rule-of-three (earlier calculation) | Joint | 424 | 122 | 152 | 30 | 0 | 272 | 13.60 | 14 | Earlier calculation; not a joint 95% limit |

1. The final pool contains 361 non-adjudicated studies (189 uncoded, 146 VAE-NT, and 26 unclear). Of the 122 verified studies, 110 are in the final pool; 10 are not in the 573-study corpus and two were adjudicated after sampling.
2. Method A is the primary finite-population result. Method C projects the binomial Clopper–Pearson upper limit of each stratum onto its unchecked count and is shown as a sensitivity analysis.
3. Method B reproduces the earlier 14-study calculation from per-stratum 95% bounds.
4. Existing nonrandom negatives contribute deterministic exclusions but do not enlarge the random-sample probability denominator under Method A.

Machine-readable companion: table_s4.csv in the Zenodo archive (11_supplementary_tables/; 16 data rows, same headers).

### Supplementary Table S5. Unit-level six-gate dispositions for index-derivation studies

This table lists all 44 claim units for the index-derivation studies: 32 units from the five identifiable index-derivation studies and 12 units from the BAD-D v4 development-validation study (FT365).

| Index | Study | Claim unit | Exit gate | Exit reason | Final verdict |
|---|---|---|---|---|---|
| CBI | FT285 | C1/S1 | 1 | Not evidence for VAE-NTT | Out of scope |
| CBI | FT285 | C2/S2 | 1 | Not evidence for VAE-NTT | Out of scope |
| CBI | FT285 | C3/S1 | 1 | Claim overreach | Adverse |
| CBI | FT285 | C4/S1 | 1 | Not evidence for VAE-NTT | Out of scope |
| TBI | FT438 | C1/S1 | 2 | Evidence incomplete | Adverse |
| TBI | FT438 | C2/S1 | 2 | Evidence incomplete | Adverse |
| TBI | FT438 | C3/S1 | 1 | Not evidence for VAE-NTT | Out of scope |
| TBI | FT438 | C4/S1 | 2 | Evidence incomplete | Adverse |
| TBIv2 | FT548 | C1/S1 | 1 | Not evidence for VAE-NTT | Out of scope |
| TBIv2 | FT548 | C2/S2 | 1 | Not evidence for VAE-NTT | Out of scope |
| TBIv2 | FT548 | C3/S1 | 1 | Reporting indeterminate | Indeterminate |
| TBIv2 | FT548 | C4/S1 | 1 | Not evidence for VAE-NTT | Out of scope |
| TBIv2 | FT548 | C5/S1 | 1 | Reporting indeterminate | Indeterminate |
| KISA% | FT509 | C1/S1 | 1 | Not evidence for VAE-NTT | Out of scope |
| KISA% | FT509 | C2/S1 | 1 | Claim overreach | Adverse |
| KISA% | FT509 | C3/S2 | 1 | Claim overreach | Adverse |
| KISA% | FT509 | C4/S3 | 1 | Claim overreach | Adverse |
| KISA% | FT509 | C5/S4 | 1 | Claim overreach | Adverse |
| KISA% | FT509 | C6/S5 | 1 | Claim overreach | Adverse |
| KISA% | FT509 | C7/S1 | 1 | Not evidence for VAE-NTT | Out of scope |
| KISA% | FT509 | C8/S1 | 1 | Not evidence for VAE-NTT | Out of scope |
| KISA% | FT509 | C9/S1 | 0 | Cause mismatch | Out of scope |
| KISA% | FT509 | C10/S1 | 1 | Claim overreach | Adverse |
| KISA% | FT509 | C11/S1 | 1 | Claim overreach | Adverse |
| KISA% | FT509 | C12/S1 | 1 | Not evidence for VAE-NTT | Out of scope |
| KISA% | FT509 | C13/S2 | 1 | Not evidence for VAE-NTT | Out of scope |
| PRFI | FT364 | C1/S1 | 3 | Reporting indeterminate | Indeterminate |
| PRFI | FT364 | C2/S2 | 2 | Evidence incomplete | Adverse |

| Index | Study | Claim unit | Exit gate | Exit reason | Final verdict |
|---|---|---|---|---|---|
| PRFI | FT364 | C3/S1 | 1 | Not evidence for VAE-NTT | Out of scope |
| PRFI | FT364 | C4/S1 | 1 | Not evidence for VAE-NTT | Out of scope |
| PRFI | FT364 | C5/S2 | 1 | Not evidence for VAE-NTT | Out of scope |
| PRFI | FT364 | C6/S3 | 1 | Not evidence for VAE-NTT | Out of scope |
| BAD-D | FT365 | C1/S1 | 1 | Not evidence for VAE-NTT | Out of scope |
| BAD-D | FT365 | C2/S2 | 1 | Not evidence for VAE-NTT | Out of scope |
| BAD-D | FT365 | C3/S1 | 1 | Not evidence for VAE-NTT | Out of scope |
| BAD-D | FT365 | C4/S2 | 1 | Not evidence for VAE-NTT | Out of scope |
| BAD-D | FT365 | C5/S1 | 1 | Not evidence for VAE-NTT | Out of scope |
| BAD-D | FT365 | C6/S2 | 1 | Not evidence for VAE-NTT | Out of scope |
| BAD-D | FT365 | C7/S1 | 1 | Reporting indeterminate | Indeterminate |
| BAD-D | FT365 | C8/S2 | 1 | Reporting indeterminate | Indeterminate |
| BAD-D | FT365 | C9/S1 | 1 | Reporting indeterminate | Indeterminate |
| BAD-D | FT365 | C10/S2 | 1 | Reporting indeterminate | Indeterminate |
| BAD-D | FT365 | C11/S1 | 2 | Evidence incomplete | Adverse |
| BAD-D | FT365 | C12/S2 | 2 | Evidence incomplete | Adverse |

1. Among the 26 Gate 1 exits in the 32-unit five-index derivation set, 16 exited because the study answered a weaker question than VAE-NTT discrimination; the attribution failure lies in the downstream cross-layer use of the index, not in the derivation study itself.
2. The deployed BAD-D v2/v3 lineage and the named ESI lineage have no identifiable public derivation study and therefore appear in the public-lineage register in Supplementary Table S6 rather than as derivation-task rows here; FT365 is the separate BAD-D v4 development-validation study.
3. Exit gate is the first terminal gate (0–5); exit reasons and final verdicts are transcribed from the adjudication records.

Machine-readable companion: table_s5.csv in the Zenodo archive (11_supplementary_tables/; 44 data rows, same headers).

### Supplementary Table S6. Public lineage register for BAD-D and ESI

This register lists all 24 public-lineage nodes (Part A) and all 25 typed relations between them (Part B) for BAD-D and ESI. Relations refer to nodes by their Node ID.

#### A. Nodes (24)

| Index | Node ID | Node type | Label | Year | Verification status | Source |
|---|---|---|---|---|---|---|
| BAD-D | badd_src_belin2009 | Source document | Belin/Ambrósio/Steinmueller 2009 trade publication (candidate source for BAD II architecture and 1.6/2.6 color rules) | 2009 | Retrieved (manufacturer-hosted, not peer reviewed; describes the display and color thresholds, no derivation) | Belin et al. 2009, September 2009 |
| BAD-D | badd_src_lyra2012 | Source document | Lyra et al. 2012 monograph chapter (candidate BAD II/III construction account) | 2012 | Unverified | Lyra et al. 2012, Ch. 10, pp. 127–129 |
| BAD-D | badd_v2 | Index version | BAD II (BAD-D v2 deployed rule; derivation not publicly reconstructable) | 2010 | Verified | Pentacam interpretation guide 2015, printed p. 89 |
| BAD-D | badd_v3 | Index version | BAD III (BAD-D v3 deployed rule; derivation not publicly reconstructable) | 2012 | Verified | Lineage synthesis in this study |
| BAD-D | badd_study_lopes2024 | Study | Lopes et al. 2024 BAD-D v4 development-validation study (FT365) | 2024 | Verified | Lopes et al. 2024 (FT365) |
| BAD-D | badd_v4 | Index version | BAD-D v4 | 2024 | Verified | Lopes et al. 2024 (FT365) |
| ESI | esi_src_langenbucher2012 | Source document | Langenbucher et al. 2012 CASIA-1000 automatic-screening conference abstract T085 (candidate precursor; ESI name absent) | 2012 | Candidate | Langenbucher et al. 2012, abstract T085 |
| ESI | esi_index | Index version | ESI deployed named rule (published by 2015; version unspecified) | 2015 | Verified | FT189; CASIA2 Application Manual p. 57 |
| ESI | esi_study_spira2015 | Study | Spira et al. 2015 | 2015 | Verified | FT189 |
| ESI | esi_study_kojima2020 | Study | Kojima et al. 2020 | 2020 | Verified | FT493 |
| ESI | esi_study_lavric2020 | Study | Lavric et al. 2020 | 2020 | Verified | FT269 |
| ESI | esi_study_alyasseri2022 | Study | Alyasseri et al. 2022 | 2022 | Verified | FT029 |
| ESI | esi_study_paul2024 | Study | Paul et al. 2024 (KeratoEL) | 2024 | Verified | FT504 |
| ESI | esi_study_mourgues2024 | Study | Mourgues et al. 2024 | 2024 | Verified | FT410 |
| ESI | esi_study_mirsalehi2025 | Study | Mirsalehi et al. 2025 (preprint/final pair deduplicated as one study) | 2025 | Verified | FT563 + FT564 |
| ESI | esi_study_mirsalehi_langenbucher 2026 | Study | Mirsalehi and Langenbucher 2026 | 2026 | Verified | FT022 |
| ESI | esi_study_yousefi2018 | Study | Yousefi et al. 2018 | 2018 | Verified | FT497; Yousefi et al. 2018 |
| ESI | esi_study_itoi2020 | Study | Itoi et al. 2020 | 2020 | Verified | FT068 |

| Index | Node ID | Node type | Label | Year | Verification status | Source |
|---|---|---|---|---|---|---|
| ESI | esi_dataset_yousefi2018 | Dataset | Yousefi/Harvard Dataverse CASIA SS-1000 public dataset with ESI-derived labels (DVN/G2CRMO) | 2018 | Verified | Yousefi et al. 2018 dataset, DOI 10.7910/DVN/G2CRMO |
| ESI | esi_study_aatila2021 | Study | Aatila et al. 2021 | 2021 | Verified | FT495 |
| ESI | esi_study_priya2022 | Study | Priya et al. 2022 | 2022 | Verified | FT506 |
| ESI | esi_study_aiiot2024 | Study | 2024 AIIOT study | 2024 | Verified | FT472 |
| ESI | esi_study_chaari2025 | Study | Chaari et al. 2025 | 2025 | Verified | FT107 |
| ESI | esi_study_oconnell | Study | O'Connell et al. conference abstract | Year unknown | Verified | O'Connell et al., abstract (DOI 10.1111/ceo.14486) |

#### B. Typed relations (25)

| Index | From | To | Relation | Evidence |
|---|---|---|---|---|
| BAD-D | badd_src_belin2009 | badd_v2 | Candidate precursor | Belin et al. 2009 retrieved; contains no derivation |
| BAD-D | badd_src_lyra2012 | badd_v2 | Candidate precursor | Lyra et al. 2012 remains unverified |
| BAD-D | badd_src_lyra2012 | badd_v3 | Candidate precursor | Candidate BAD III account is developmental, not a deployed equation |
| BAD-D | badd_v2 | badd_v3 | Version succession | BAD II (2010) to BAD III (2012) |
| BAD-D | badd_v3 | badd_v4 | Version succession | Lineage synthesis in this study |
| BAD-D | badd_study_lopes2024 | badd_v4 | Derivation | FT365 |
| ESI | esi_src_langenbucher2012 | esi_index | Candidate precursor | Candidate only because T085 does not use the ESI name or prove equivalence |
| ESI | esi_index | esi_study_spira2015 | Direct use | Spira et al. 2015 (FT189) |
| ESI | esi_index | esi_study_kojima2020 | Direct use | Kojima et al. 2020 (FT493) |
| ESI | esi_index | esi_study_lavric2020 | Direct use | Lavric et al. 2020 (FT269) |
| ESI | esi_index | esi_study_alyasseri2022 | Direct use | Alyasseri et al. 2022 (FT029) |
| ESI | esi_index | esi_study_paul2024 | Direct use | Paul et al. 2024 (FT504) |
| ESI | esi_index | esi_study_mourgues2024 | Direct use | Mourgues et al. 2024 (FT410) |
| ESI | esi_index | esi_study_mirsalehi2025 | Direct use | Mirsalehi et al. 2025 (FT563 + FT564) |
| ESI | esi_index | esi_study_mirsalehi_langenbucher2026 | Direct use | Mirsalehi and Langenbucher 2026 (FT022) |
| ESI | esi_index | esi_study_yousefi2018 | Direct use | Yousefi et al. 2018 (FT497) |
| ESI | esi_index | esi_study_itoi2020 | Direct use | Itoi et al. 2020 (FT068) |
| ESI | esi_study_yousefi2018 | esi_dataset_yousefi2018 | Derivation | FT497 and DOI 10.7910/DVN/G2CRMO |
| ESI | esi_dataset_yousefi2018 | esi_study_lavric2020 | Label inheritance | Lavric et al. 2020 (FT269) |
| ESI | esi_dataset_yousefi2018 | esi_study_aatila2021 | Label inheritance | Aatila et al. 2021 (FT495) |
| ESI | esi_dataset_yousefi2018 | esi_study_priya2022 | Label inheritance | Priya et al. 2022 (FT506) |
| ESI | esi_dataset_yousefi2018 | esi_study_paul2024 | Label inheritance | Paul et al. 2024 (FT504) |

| Index | From | To | Relation | Evidence |
|---|---|---|---|---|
| ESI | esi_dataset_yousefi2018 | esi_study_aiiot2024 | Label inheritance | 2024 AIIOT study (FT472) |
| ESI | esi_dataset_yousefi2018 | esi_study_chaari2025 | Label inheritance | Chaari et al. 2025 (FT107) |
| ESI | esi_dataset_yousefi2018 | esi_study_oconnell | Label inheritance | O'Connell et al., abstract (DOI 10.1111/ceo.14486) |

1. The deployed BAD-D v2/v3 rules have no publicly reproducible peer-reviewed derivation. The named ESI likewise has no peer-reviewed derivation; the report by Yousefi et al. (2018) states that the ESI formula is unclear.
2. Belin 2009 was retrieved from the manufacturer's website and contains no derivation; Lyra 2012 remains unverified, and T085 remains a candidate. Candidate precursor is a distinct relation type and is not derivation.
3. The register contains 10 direct-use relations and 7 label-inheritance relations.
4. The O'Connell et al. entry has no verified publication year in the source register and is therefore shown as year unknown.
5. Lineage assertions were compiled in the index-provenance audit described in Methods; the public sources supporting each node and relation are given in the Source and Evidence columns.

Machine-readable companion: table_s6.csv in the Zenodo archive (11_supplementary_tables/; 49 data rows, same headers).

### Supplementary Table S7. Study-level partial-identification bounds

This table reports all 46 eligible studies, including the source and evidence grade for π, threshold-specific sample information, AUC (NT), the main bound, and both sensitivity bounds.

| Study | Year | Target eyes (n) | Model | AUC (NT) | π source and grade (BAD-D 1.60) | π at 1.60 | AUC (NTT) main bound, 1.60 | AUC (NTT) sensitivity bound, 1.45 | AUC (NTT) sensitivity bound, 2.11 | Location in source report |
|---|---|---|---|---|---|---|---|---|---|---|
| FT006 | 2019 | 100 | TBI | 0.901 | π-C: Distribution-free quantile/range interval | [0.500, 1.000] | [0.000, 1.000] | [0.000, 1.000] | [0.802, 1.000] | Table 5; Table 1 |
| FT015 | 2020 | 62 | CH+BAD-D model | 0.948 | π-A: 52/62 | 0.839 | [0.678, 1.000] | Not estimable; threshold-specific BAD-D distribution not reported | Not estimable; threshold-specific BAD-D distribution not reported | Abstract/Table 3; Results |
| FT044 | 2021 | 50 | Pentacam+OPD-Scan model | 0.930 | π-B: Normal-tail approximation | 0.641 | [0.805, 1.000] | [0.775, 1.000] | [0.868, 1.000] | Figure 1/Table 3; Table 1 |
| FT045 | 2024 | 30 | RMS HOA | 0.789 | π-C: Distribution-free quantile/range interval | [0.250, 0.500] | [0.578, 1.000] | [0.156, 1.000] | [0.578, 1.000] | Table 2; Table 1 |
| FT046 | 2023 | 70 | BAD-D | 0.965 | π-C: Distribution-free quantile/range interval | [0.750, 1.000] | [0.000, 1.000] | [0.000, 1.000] | [0.000, 1.000] | Table 6; Table 4 |
| FT068 | 2020 | 13 | As/Ps | 0.980 | π-B: Normal-tail approximation | 0.746 | [0.921, 1.000] | [0.898, 1.000] | [0.960, 1.000] | Table 2; Table 1 |
| FT073 | 2021 | 27 | TBI | 0.928 | π-B: Normal-tail approximation | 0.780 | [0.673, 1.000] | [0.521, 1.000] | [0.869, 1.000] | Table 3a; Table 2 |
| FT074 | 2017 | 18 | BAD-D+Bowman roughness | 0.850 | π-B: Normal-tail approximation | 0.819 | [0.171, 1.000] | [0.000, 1.000] | [0.671, 1.000] | Abstract/Table 2; Table 1 |
| FT086 | 2026 | 33 | mean above-threshold correlation of dynamics | 0.870 | π-C: Distribution-free quantile/range interval | [0.000, 0.500] | [0.740, 1.000] | [0.740, 1.000] | [0.740, 1.000] | External validation VAE-NT vs N ROC, Figure S8; External validation Table 2 |
| FT097 | 2019 | 33 | 4-variable model | 0.846 | π-B: Normal-tail approximation | 0.443 | [0.723, 1.000] | [0.673, 1.000] | [0.809, 1.000] | Abstract/results; Table 1 |
| FT152 | 2019 | 21 | TBI | 0.804 | π-B: Normal-tail approximation | 0.785 | [0.089, 1.000] | [0.000, 1.000] | [0.323, 1.000] | Table 3; Abstract/Table 1 |
| FT154 | 2023 | 84 | RF Model 6 (RTVue-XR+Corvis ST) | 0.902 | π-B: Normal-tail approximation | 0.381 | [0.842, 1.000] | [0.813, 1.000] | [0.887, 1.000] | Results, Table 3; Table 1 |
| FT159 | 2022 | 69 | RF (SD-OCT+Corvis) | 0.990 | π-B: Normal-tail approximation | 0.388 | [0.984, 1.000] | [0.981, 1.000] | [0.988, 1.000] | Results; Table 1 |
| FT167 | 2018 | 30 | anterior elevation (EA) | 0.935 | π-B: Normal-tail approximation | 0.873 | [0.488, 1.000] | [0.287, 1.000] | [0.792, 1.000] | ROC text/Table 2; Table 1 |
| FT185 | 2022 | 30 | TBI | 0.910 | π-B: Normal-tail approximation | 1.000 | [0.000, 1.000] | [0.000, 1.000] | [0.000, 1.000] | Table 4; Table 3 |
| FT187 | 2021 | 83 | AstigF | 0.862 | π-C: Distribution-free quantile/range interval | [0.250, 0.500] | [0.724, 1.000] | [0.724, 1.000] | [0.816, 1.000] | Table 3; Table 2 |
| FT193 | 2021 | 15 | SLR2 | 0.965 | π-B: Normal-tail approximation | 0.702 | [0.883, 1.000] | [0.860, 1.000] | [0.928, 1.000] | Table 3; Table 1 |
| FT253 | 2015 | 146 | BAD-D | 0.712 | π-B: Normal-tail approximation | 0.672 | [0.123, 1.000] | [0.036, 1.000] | [0.339, 1.000] | Table 2; Table 1 |
| FT293 | 2022 | 58 | CHAID/CART training model | 0.940 | π-B: Normal-tail approximation | 0.670 | [0.818, 1.000] | [0.784, 1.000] | [0.886, 1.000] | Results; Training-database Table 3 |

| Study | Year | Target eyes (n) | Model | AUC (NT) | π source and grade (BAD-D 1.60) | π at 1.60 | AUC (NTT) main bound, 1.60 | AUC (NTT) sensitivity bound, 1.45 | AUC (NTT) sensitivity bound, 2.11 | Location in source report |
|---|---|---|---|---|---|---|---|---|---|---|
| FT295 | 2021 | 20 | DDA mean Q4 alpha | 0.970 | π-B: Normal-tail approximation | 0.500 | [0.940, 1.000] | [0.922, 1.000] | [0.964, 1.000] | Abstract/Table 3; Table 1 |
| FT313 | 2023 | 112 | cCBI | 0.891 | π-B: Normal-tail approximation | 0.730 | [0.596, 1.000] | [0.511, 1.000] | [0.761, 1.000] | Results §2.2.2; Table 1 |
| FT317 | 2024 | 34 | BAD-D | 0.781 | π-C: Distribution-free quantile/range interval | [0.500, 0.750] | [0.124, 1.000] | [0.124, 1.000] | [0.562, 1.000] | Table 4; Table 1 |
| FT335 | 2018 | 30 | Scheimpflug+SD-OCT model | 1.000 | π-B: Normal-tail approximation | 0.480 | [1.000, 1.000] | [1.000, 1.000] | [1.000, 1.000] | Results, model table; Table 1 |
| FT345 | 2020 | 31 | CBI | 0.909 | π-B: Normal-tail approximation | 0.703 | [0.693, 1.000] | [0.654, 1.000] | [0.784, 1.000] | FFK vs normal ROC text; Baseline Table 1 |
| FT349 | 2018 | 27 | IHD (baseline) | 0.788 | π-B: Normal-tail approximation | 0.038 | [0.780, 0.819] | [0.769, 0.859] | [0.788, 0.789] | Baseline ROC, abstract/results; Baseline D-index text |
| FT358 | 2016 | 21 | FECTB combined model | 0.953 | π-B: Normal-tail approximation | 0.571 | [0.890, 1.000] | [0.878, 1.000] | [0.919, 1.000] | Abstract/results; Table 3 |
| FT359 | 2019 | 57 | TBI | 0.960 | π-B: Normal-tail approximation | 0.616 | [0.896, 1.000] | [0.869, 1.000] | [0.940, 1.000] | ROC table (VAE-NT vs normal); Group table |
| FT364 | 2018 | 188 | PRFI | 0.968 | π-B: Normal-tail approximation | 0.572 | [0.925, 1.000] | [0.913, 1.000] | [0.949, 1.000] | Table 7; VAE-NT group table |
| FT371 | 2025 | 50 | epithelial SN-IT | 0.680 | π-B: Normal-tail approximation | 0.605 | [0.190, 1.000] | [0.000, 1.000] | [0.540, 0.977] | Abstract; Table 1 |
| FT374 | 2025 | 146 | cTBI | 0.930 | π-B: Normal-tail approximation | 0.427 | [0.878, 1.000] | [0.832, 1.000] | [0.925, 0.993] | Table 3; Table 1 |
| FT384 | 2020 | 15 | BAD-D | 0.859 | π-A: 0/15 | 0.000 | [0.859, 0.859] | [0.824, 1.000] | [0.859, 0.859] | Abstract/ROC Table 3; Methods/Table 1 |
| FT438 | 2017 | 94 | TBI | 0.985 | π-A: 40/94 | 0.426 | [0.974, 1.000] | [0.963, 1.000] | [0.980, 1.000] | Table B; Table A |
| FT511 | 2023 | 102 | BAD-D | 0.892 | π-B: Normal-tail approximation | 0.718 | [0.618, 1.000] | [0.538, 1.000] | [0.773, 1.000] | Table 3; Table 1 |
| FT542 | 2017 | 48 | ARTmax | 0.880 | π-B: Normal-tail approximation | 0.345 | [0.817, 1.000] | [0.778, 1.000] | [0.870, 0.954] | Table 2; Table 1 |
| FT548 | 2023 | 551 | TBIv2 | 0.945 | π-C: Distribution-free quantile/range interval | [0.500, 1.000] | [0.000, 1.000] | [0.000, 1.000] | [0.890, 1.000] | Table 3; Table 4 |
| FT655 | 2024 | 73 | TBI | 0.776 | π-A: Reported 61.6% sensitivity of BAD-D = 1.60 for VAE-NT | 0.616 | [0.417, 1.000] | [0.315, 1.000] | [0.682, 1.000] | Table 4 / §3.2; Table 1; cutoff sensitivity |
| FT647 | 2024 | 43 | EfficientNet-B0 Model 2 | 0.960 | π-A: 29/43 | 0.674 | [0.877, 1.000] | Not estimable; threshold-specific BAD-D distribution not reported | Not estimable; threshold-specific BAD-D distribution not reported | Abstract/results; BAD-D rule/results |
| FT674 | 2020 | 151 | Final D | 0.858 | π-B: Normal-tail approximation | 0.698 | [0.531, 1.000] | [0.421, 1.000] | [0.731, 1.000] | Table 3; Table 2 |
| FT699 | 2022 | 34 | BAD-D | 0.781 | π-C: Distribution-free quantile/range interval | [0.500, 1.000] | [0.000, 1.000] | [0.000, 1.000] | [0.562, 1.000] | Results; Group median table |
| FT712 | 2019 | 23 | TBI | 0.751 | π-A: 9/23 | 0.391 | [0.591, 1.000] | [0.491, 1.000] | [0.649, 1.000] | Table 2 / text; Table A / text |
| FT233 | 2026 | 19 | CBI | 0.822 | π-A: 0/19 | 0.000 | [0.822, 0.822] | [0.782, 1.000] | [0.822, 0.822] | ROC analysis; Table 2/inclusion criteria |
| FT706 | 2019 | 21 | TBI | 0.790 | π-A: 0/21 | 0.000 | [0.790, 0.790] | [0.738, 0.987] | [0.790, 0.790] | Abstract/ROC analysis; Table 1 and Discussion |
| FT707 | 2022 | 24 | TBI | 0.824 | π-A: 0/24 | 0.000 | [0.824, 0.824] | [0.785, 1.000] | [0.824, 0.824] | Table 2; Table 1 |

| Study | Year | Target eyes (n) | Model | AUC (NT) | π source and grade (BAD-D 1.60) | π at 1.60 | AUC (NTT) main bound, 1.60 | AUC (NTT) sensitivity bound, 1.45 | AUC (NTT) sensitivity bound, 2.11 | Location in source report |
|---|---|---|---|---|---|---|---|---|---|---|
| FT709 | 2017 | 21 | CBI | 0.785 | π-B: Normal-tail approximation | 0.606 | [0.455, 1.000] | [0.323, 1.000] | [0.678, 1.000] | ROC results; Table 1 |
| FT748 | 2015 | 27 | CRF | 0.866 | Not estimable; source metric is BDE, not BAD-D | Not estimable; source metric is BDE, not BAD-D | Not estimable; source metric is BDE, not BAD-D | Not estimable; source metric is BDE, not BAD-D | Not estimable; source metric is BDE, not BAD-D | Table 3; Table 1 |
| FT757 | 2018 | 18 | TBI | 0.732 | π-A: 0/18 | 0.000 | [0.732, 0.732] | [0.643, 0.976] | [0.732, 0.732] | Table B/Discussion; Inclusion/Table A |

1. AUC (NT) is the reported discrimination for the normal-versus-target construction; AUC (NTT) is the partially identified discrimination after the π-based target correction. Bracketed values are partial-identification bounds computed from the reported AUC and π, not sampling confidence intervals.
2. The main threshold is 1.60; sensitivity thresholds are 1.45 and 2.11. Eyes at or above a threshold count as flagged; where a source reported its count with a strict inequality, that count was used as reported. π-A, π-B, and π-C denote descending evidence tiers defined in the source analysis.
3. FT015 and FT647 lack threshold-specific BAD-D distributions for the two sensitivity analyses and are not estimable. FT748 reports BDE rather than BAD-D, so its π and all three bounds are not estimable on the required threshold scale.

Machine-readable companion: table_s7.csv in the Zenodo archive (11_supplementary_tables/; 46 data rows, same headers).

### Supplementary Table S8. Target-swap, refit, and pathway-reconstruction results

This table reports the exact Fig. 4 target definitions, denominators, AUCs, confidence intervals, target-swap gaps, retention, target-specific refit, reconstruction, directional-field, and permutation results.

| Component | Estimand or target | Definition | Positives (n) | Negatives or total (n) | Estimate | 95% CI lower | 95% CI upper | Unit | Method |
|---|---|---|---|---|---|---|---|---|---|
| Fixed-score transport | Operational ESI-early | Target-eye anterior ESI 5–29 | 179 | 1,124 | 0.996 | 0.994 | 0.998 | AUC | Same cross-fitted operational axis; no transport-target refit |
| Fixed-score transport | FFearly (fellow-eye defined) | Target-eye ESI-normal; partner eye ESI-early | 124 | 1,124 | 0.682 | 0.630 | 0.734 | AUC | Same cross-fitted operational axis; no transport-target refit |
| Fixed-score transport | FFKC proxy | Target-eye ESI-normal; partner eye ESI-KC | 86 | 1,124 | 0.549 | 0.479 | 0.618 | AUC | Same cross-fitted operational axis; no transport-target refit |
| Target-swap gap | Operational minus FFearly | Paired AUC attenuation from operational target | 124 | 1,124 controls; 179 operational | 0.315 | 0.262 | 0.366 | Delta AUC | Joint 2,000-replicate patient-cluster bootstrap |
| Target-swap gap | Operational minus FFKC proxy | Paired AUC attenuation from operational target | 86 | 1,124 controls; 179 operational | 0.447 | 0.378 | 0.517 | Delta AUC | Joint 2,000-replicate patient-cluster bootstrap |
| Target retention | FFKC proxy | (AUC FFKC proxy − 0.5)/(AUC operational − 0.5) | 86 | 1,124 controls; 179 operational | 0.099 | −0.042 | 0.238 | Retention | Derived within each joint bootstrap replicate |
| Target-specific refit | Contralateral ESI-KC proxy | Native-298 model refit directly to proxy target | 66 | 1,095 | 0.529 | 0.461 | 0.600 | AUC | QC-first specification (quality control before relationship definition; note 4) |
| Target-specific refit | Refit permutation test | Two-sided add-one permutation P for \|AUC − 0.5\| | NA | 1,000 permutations (551 at or beyond the observed statistic) | 0.551 | NA | NA | P value | Test statistic \|AUC − 0.5\|; add-one P = (551 + 1)/(1,000 + 1) |
| Pathway reconstruction | Continuous anterior ESI | Out-of-fold reconstruction from 298 native features | NA | 3,060 eyes; 1,733 patients | 0.991 | 0.989 | 0.993 | Pearson r | Patient-grouped five-fold out-of-fold; 2,000 patient bootstraps |
| Pathway reconstruction | ESI-early label recovery | ESI-early eyes versus bilateral ESI-true-normal eyes; out-of-fold predicted continuous anterior ESI is the score | 199 | 2,190 | 0.995 | 0.993 | 0.997 | AUC | Out-of-fold reconstructed continuous ESI score |
| Directional field | Native-298 directions | With feature direction fixed on the operational comparison, operational-target AUC exceeds FFKC-proxy AUC | 278 | 298 | 278/298 | NA | NA | Features | One correlated descriptive feature field |

1. ESI grades follow the anterior ESI: ESI-normal 0–4 (every observed normal value is 0), ESI-early 5–29, ESI-KC 30 or above. Strict controls are bilateral ESI-normal eyes. Operational ESI-early, FFearly fellow-eye, and FFKC proxy targets contain 179, 124, and 86 eyes, respectively; all fixed-score analyses use 1,124 strict-control eyes.
2. The transport model is an XGBoost classifier fit only to operational ESI-early versus strict controls, with fold-local imputation and patient-locked folds. The 95% CI columns are the 2.5th and 97.5th percentiles of 2,000 patient-level bootstrap replicates.
3. Target retention of 0.099 means 9.9% of above-chance operational discrimination was retained under the FFKC proxy.
4. The target-specific refit uses the QC-first specification, in which morphology quality control preceded the bilateral relationship definition (1,095 strict-control and 66 FFKC-proxy eyes); the fixed-score rows use the primary specification (1,124 and 86). Under the QC-first specification the fixed-score Target-Swap Gap is 0.437 (95% CI 0.352–0.521) instead of 0.447 (Methods).

Machine-readable companion: table_s8.csv in the Zenodo archive (11_supplementary_tables/; 11 data rows, same headers).

### Supplementary Table S9. Cross-device, BAD-D-increment, and demographic-probe results

This table reports the exact Fig. 5 cross-device train-to-test results (Part A), the same-device BAD-D increment in the Almería cohort (Part B), the demographic-only models and group-balance statistics (Part C), and the corpus-level demographic safeguard counts (Part D).

#### A. Cross-device transport

| Train | Test or cohort | Model | Target | Feature set or statistic | Train positives (n) | Train negatives (n) | Test or cohort positives (n) | Test or cohort negatives (n) | Within-test-cohort CV AUC | Estimate | Note |
|---|---|---|---|---|---|---|---|---|---|---|---|
| Almería Pentacam | CASIA-OCT | Logistic regression | Manifest keratoconus | Eight harmonized native features | 83 | 82 | 162 | 2,699 | 0.997 | 0.945 | Point AUC only (note 1) |
| CASIA-OCT | Almería Pentacam | Logistic regression | Manifest keratoconus | Eight harmonized native features | 162 | 2,699 | 83 | 82 | 0.982 | 0.989 | Point AUC only (note 1) |
| Almería Pentacam | CASIA-OCT | XGBoost | Manifest keratoconus | Eight harmonized native features | 83 | 82 | 162 | 2,699 | 0.998 | 0.993 | Point AUC only (note 1) |
| CASIA-OCT | Almería Pentacam | XGBoost | Manifest keratoconus | Eight harmonized native features | 162 | 2,699 | 83 | 82 | 0.981 | 0.991 | Point AUC only (note 1) |
| Almería Pentacam | CASIA-OCT | Logistic regression | Early or subclinical (CASIA ESI-early; Almeria pre-labeled subclinical) | Eight harmonized native features | 40 | 82 | 199 | 2,699 | 0.958 | 0.759 | Point AUC only (note 1) |
| CASIA-OCT | Almería Pentacam | Logistic regression | Early or subclinical (CASIA ESI-early; Almeria pre-labeled subclinical) | Eight harmonized native features | 199 | 2,699 | 40 | 82 | 0.830 | 0.750 | Point AUC only (note 1) |
| Almería Pentacam | CASIA-OCT | XGBoost | Early or subclinical (CASIA ESI-early; Almeria pre-labeled subclinical) | Eight harmonized native features | 40 | 82 | 199 | 2,699 | 0.957 | 0.677 | Point AUC only (note 1) |
| CASIA-OCT | Almería Pentacam | XGBoost | Early or subclinical (CASIA ESI-early; Almeria pre-labeled subclinical) | Eight harmonized native features | 199 | 2,699 | 40 | 82 | 0.737 | 0.726 | Point AUC only (note 1) |

#### B. Same-device analyses in the Almería cohort

| Section | Model | Target | Feature set or statistic | Test or cohort positives (n) | Test or cohort negatives (n) | Estimate | 95% CI lower | 95% CI upper | Note |
|---|---|---|---|---|---|---|---|---|---|
| Same-device BAD-D increment | XGBoost | Pre-labeled subclinical | 40 non-composite measurements | 40 | 82 | 0.940 | 0.880 | 0.983 | 40 features |
| Same-device BAD-D increment | XGBoost | Pre-labeled subclinical | 40 measurements + BAD-D | 40 | 82 | 0.937 | 0.876 | 0.982 | 41 features |
| Same-device BAD-D increment | XGBoost | Manifest keratoconus | 40 non-composite measurements | 83 | 82 | 0.994 | 0.984 | 1.000 | 40 features |
| Same-device BAD-D increment | XGBoost | Manifest keratoconus | 40 measurements + BAD-D | 83 | 82 | 1.000 | 1.000 | 1.000 | 41 features |
| Same-device BAD-D alone | BAD-D | Pre-labeled subclinical | BAD-D only | 40 | 82 | 0.933 | NA | NA | Point AUC only (note 1) |
| Same-device BAD-D alone | BAD-D | Manifest keratoconus | BAD-D only | 83 | 82 | 0.996 | NA | NA | Point AUC only (note 1) |
| Same-device BAD-D increment | Paired eye bootstrap | Pre-labeled subclinical | ΔAUC (with BAD-D minus without) | 40 | 82 | −0.003 | −0.031 | 0.019 | Delta AUC |

| Section | Model | Target | Feature set or statistic | Test or cohort positives (n) | Test or cohort negatives (n) | Estimate | 95% CI lower | 95% CI upper | Note |
|---|---|---|---|---|---|---|---|---|---|
| Same-device BAD-D increment | Paired eye bootstrap | Manifest keratoconus | ΔAUC (with BAD-D minus without) | 83 | 82 | 0.006 | 0.000 | 0.016 | Delta AUC |

#### C. Demographic-only models and group balance in the Almería cohort

| Section | Model | Target | Feature set or statistic | Test or cohort positives (n) | Test or cohort negatives (n) | Estimate | 95% CI lower | 95% CI upper | P value | Note |
|---|---|---|---|---|---|---|---|---|---|---|
| Demographic-only model | XGBoost | Pre-labeled subclinical | Age + sex | 40 | 82 | 0.771 | 0.678 | 0.854 | NA | Five-fold cross-fitting; eye bootstrap |
| Demographic-only model | XGBoost | Pre-labeled subclinical | Age only | 40 | 82 | 0.749 | 0.652 | 0.840 | NA | Five-fold cross-fitting; eye bootstrap |
| Demographic-only model | XGBoost | Manifest keratoconus | Age + sex | 83 | 82 | 0.780 | 0.705 | 0.850 | NA | Five-fold cross-fitting; eye bootstrap |
| Demographic-only model | XGBoost | Manifest keratoconus | Age only | 83 | 82 | 0.804 | 0.736 | 0.866 | NA | Five-fold cross-fitting; eye bootstrap |
| Demographic-only model | Permutation test | Manifest keratoconus | Age + sex, permutation P | 83 | 82 | 0.780 | NA | NA | 0.005 | 200 permutations; null mean 0.499 |
| Demographic-only model | Permutation test | Pre-labeled subclinical | Age + sex, permutation P | 40 | 82 | 0.771 | NA | NA | 0.005 | 200 permutations; null mean 0.504 |
| Demographic balance | NA | Manifest keratoconus | Age, two-sided Mann–Whitney U P | 83 | 82 | NA | NA | NA | 0.665 | Two-sided test |
| Demographic balance | NA | Manifest keratoconus | Control-to-case age s.d. ratio | 83 | 82 | 1.540 | NA | NA | NA | Descriptive dispersion ratio |
| Demographic balance | NA | Pre-labeled subclinical | Age, two-sided Mann–Whitney U P | 40 | 82 | NA | NA | NA | 0.976 | Two-sided test |
| Demographic balance | NA | Pre-labeled subclinical | Control-to-case age s.d. ratio | 40 | 82 | 1.750 | NA | NA | NA | Descriptive dispersion ratio |

#### D. Demographic safeguards across the 573-study corpus

| Item | Studies (n) | Denominator | Note |
|---|---|---|---|
| Independence check beyond location testing | 3 | 573 | Three studies performed a stronger independence check |
| Demographic variables used as model inputs | 31 | 573 | Fixed 573-study denominator |
| Mean/proportion location test reported | 124 | 573 | Fixed 573-study denominator |
| Overlap: demographic inputs and location testing | 8 | 573 | Subset of the 31 and 124 study counts |
| Demographic-input studies with a location test but no independence check | 7 | 573 | Subset of the 8 overlap studies |

1. Cross-device rows pair each transported AUC with the within-test-cohort cross-validation AUC of the same model family; confidence intervals were not computed for the cross-device transports or for BAD-D alone. All other 95% CI values are the 2.5th and 97.5th percentiles of 2,000 eye-level bootstrap resamples.
2. The Almería subclinical grade was assigned in the source dataset (pre-labeled). BAD-D alone reaches AUC 0.933 for that grade, and adding BAD-D to the 40 non-composite measurements changes AUC by −0.003 (95% CI −0.031 to +0.019).
3. Part D uses the fixed denominator of 573 studies and reports study counts; 8 studies overlap the demographic-input and location-test groups, and 7 of those 8 report a location test without an independence check.

Machine-readable companion: table_s9.csv in the Zenodo archive (11_supplementary_tables/; 31 data rows, same headers).

### Supplementary Table S10. Cell-level evidence for Table 3

#### A. Evidence statements for the 42 application cells

| Module | Reporting item | Study | Table 3 status | Evidence statement | Source | Source excerpt(s) |
|---|---|---|---|---|---|---|
| Mandatory core | Claim stratum (CORE-1) | Karimi et al., 2018 (FT126) | Conditional | A suspect stratum is named, but it is defined by persistent diagnostic uncertainty rather than by stated eligibility criteria. | Evidence record karimi-et-al-2018-ft126 (Zenodo archive, 02_claim_adjudication) | “the suspect to keratoconus eyes refers to the eyes that even after a deep examination of the eye by ophthalmologists and different devices, still the doctors cannot make sure that whether the eye is healthy or keratoconus” |
| Mandatory core | Claim stratum (CORE-1) | Issarti et al., 2020 (FT512) | Met | Five strata are explicit; the suspect/FFKC target is the asymptomatic target eye with no clinical signs of ectasia or tomographic changes, while the contralateral eye has clinical keratoconus. | Evidence record issarti-et-al-2020-ft512 (Zenodo archive, 02_claim_adjudication) | “In this study five groups were considered: normal, suspect keratoconus, early, mild, and moderate to advanced keratoconus”; “the contralateral, asymptomatic eye of a subject with clinical keratoconus in one eye, showing no clinical signs of ectasia or tomographic changes.” |
| Mandatory core | Claim stratum (CORE-1) | Wahba et al., 2018 (FT666) | Met | FFKC is explicitly defined by target-eye normality across clinical, curvature, elevation, and pachymetry domains plus evident keratoconus in the fellow eye. | Evidence record wahba-et-al-2018-ft666 (Zenodo archive, 02_claim_adjudication) | “a cornea that has no abnormal finding by clinical examination, curvature, elevation, and pachymetry parameters with evident KC in the fellow eye” |
| Mandatory core | Claim stratum (CORE-1) | Saad & Gatinel, 2012 (FT686) | Conditional | The validation population is predefined as normal versus at risk for LASIK, but the at-risk stratum was assigned by one surgeon's subjective reading of the maps rather than by stated criteria. | Evidence record saad-gatinel-2012-ft686 (Zenodo archive, 02_claim_adjudication) | “retrospectively evaluated by one experimented refractive surgeon and classified as ‘normal’ or ‘at risk for LASIK’.” |
| Mandatory core | Claim stratum (CORE-1) | Ruiz Hidalgo et al., 2017 (FT687) | Conditional | Clinical groups are explicit, but the SS group combines forme fruste and suspect eyes under center-specific workflows. | Evidence record ruiz-hidalgo-et-al-2017-ft687 (Zenodo archive, 02_claim_adjudication) | “classified into one of the following groups: normal (N), keratoconus (KC), forme fruste and suspect keratoconus (SS), and postrefractive (PR) surgery for myopia” |
| Mandatory core | Claim stratum (CORE-1) | Chan et al., 2015 (FT688) | Met | Two groups are specified; target-eye normality is defined by clinical and topographic examination plus negative KCI/KSI, no steep keratometry (>47 D), no maximum posterior elevation >40 µm and no asymmetric anterior keratographic pattern, with frank keratoconus in the contralateral eye. | Evidence record chan-et-al-2015-ft688 (Zenodo archive, 02_claim_adjudication) | “The FFKC group involved clinically and topographically ‘normal’ eyes with the contralateral eye showing frank keratoconus”; “including a negative Tomey keratoconus screening report [using the Klyce/Maeda keratoconus index (KCI) and the Smolek/Klyce index (KSI)] … and without a steep keratometric curvature of >47 D, maximum posterior corneal elevation of >40 µm, and an asymmetric anterior keratographic pattern in the axial power map” |

| Module | Reporting item | Study | Table 3 status | Evidence statement | Source | Source excerpt(s) |
|---|---|---|---|---|---|---|
| Mandatory core | Claim stratum (CORE-1) | Herber et al., 2022 (FT378) | Met | VAE-NT and VAE-NTT are separated explicitly, with named topographic and tomographic thresholds. | Evidence record herber-et-al-2022-ft378 (Zenodo archive, 02_claim_adjudication) | "the fellow eye appeared topographically normal with a KISA% of less than 60, I-S value of less than 1.45 D, Kmax of less than 47.00 D (VAE-NT), or topographically and tomographically normal … and BAD-D of less than 1.6 (VAE-NTT)" |
| Mandatory core | Positive-class identity (CORE-2) | Karimi et al., 2018 (FT126) | Conditional | Three ophthalmologists contributed to identity assignment, but Corvis and Pentacam also participated in the identification pathway. | Evidence record karimi-et-al-2018-ft126 (Zenodo archive, 02_claim_adjudication) | "The healthy and keratoconus eyes were identified via the Corvis and Pentacam by help of three ophthalmologists." |
| Mandatory core | Positive-class identity (CORE-2) | Issarti et al., 2020 (FT512) | Met | Suspect identity is anchored by clinical keratoconus in the contralateral eye; the target eye is asymptomatic and has no clinical or tomographic changes. | Evidence record issarti-et-al-2020-ft512 (Zenodo archive, 02_claim_adjudication) | "the contralateral, asymptomatic eye of a subject with clinical keratoconus in one eye, showing no clinical signs of ectasia or tomographic changes." |
| Mandatory core | Positive-class identity (CORE-2) | Wahba et al., 2018 (FT666) | Met | FFKC identity uses evident keratoconus in the fellow eye while the target eye is normal across stated domains. | Evidence record wahba-et-al-2018-ft666 (Zenodo archive, 02_claim_adjudication) | "with evident KC in the fellow eye" |
| Mandatory core | Positive-class identity (CORE-2) | Saad & Gatinel, 2012 (FT686) | Not demonstrated | Positive identity is a target-eye classification by the authors based only on Placido and elevation maps, not an outside-pathway clinical or partner-eye anchor. | Evidence record saad-gatinel-2012-ft686 (Zenodo archive, 02_claim_adjudication) | "The topographies were classified by the authors as either 'suitable' or 'at risk' for LASIK surgery based only on the Placido and elevation maps." |
| Mandatory core | Positive-class identity (CORE-2) | Ruiz Hidalgo et al., 2017 (FT687) | Conditional | The forme fruste component has a partner-eye anchor, but the combined SS group also contains dubious suspect eyes. | Evidence record ruiz-hidalgo-et-al-2017-ft687 (Zenodo archive, 02_claim_adjudication) | "forme fruste was defined as the tomographically normal companion eyes of eyes with obvious keratoconus, and keratoconus suspect eyes were the dubious cases" |
| Mandatory core | Positive-class identity (CORE-2) | Chan et al., 2015 (FT688) | Met | Contralateral keratoconus was reconfirmed by a corneal subspecialist through clinical examination and topography. | Evidence record chan-et-al-2015-ft688 (Zenodo archive, 02_claim_adjudication) | "The diagnosis of keratoconus in the contralateral eye was reconfirmed by clinical examination and evaluation of topographies … by a corneal subspecialist" |
| Mandatory core | Positive-class identity (CORE-2) | Herber et al., 2022 (FT378) | Met | Positive identity is clinical keratoconus in the partner eye. | Evidence record herber-et-al-2022-ft378 (Zenodo archive, 02_claim_adjudication) | "one eye had clinical keratoconus (VAE-E)" |
| Mandatory core | Per-eye exclusion rulers (CORE-3) | Karimi et al., 2018 (FT126) | Not reported | A complete ophthalmic examination is reported, but the prespecified exclusion rulers and their per-eye results are not. | Evidence record karimi-et-al-2018-ft126 (Zenodo archive, 02_claim_adjudication) | "All participants underwent a complete ophthalmic examination" |
| Mandatory core | Per-eye exclusion rulers (CORE-3) | Issarti et al., 2020 (FT512) | Not demonstrated | Target-eye normality is narrative; no named numeric exclusion ruler is reported in the suspect definition. | Evidence record issarti-et-al-2020-ft512 (Zenodo archive, 02_claim_adjudication) | "showing no clinical signs of ectasia or tomographic changes." |
| Mandatory core | Per-eye exclusion rulers (CORE-3) | Wahba et al., 2018 (FT666) | Conditional | Clinical, curvature, elevation, and pachymetry domains are stated, but numeric ruler cut-offs are absent from the group definition. | Evidence record wahba-et-al-2018-ft666 (Zenodo archive, 02_claim_adjudication) | "no abnormal finding by clinical examination, curvature, elevation, and pachymetry parameters" |

| Module | Reporting item | Study | Table 3 status | Evidence statement | Source | Source excerpt(s) |
|---|---|---|---|---|---|---|
| Mandatory core | Per-eye exclusion rulers (CORE-3) | Saad & Gatinel, 2012 (FT686) | Not demonstrated | Target eyes were subjectively classified from Placido and elevation maps rather than certified by reported per-eye rulers. | Evidence record saad-gatinel-2012-ft686 (Zenodo archive, 02_claim_adjudication) | "classified by the authors … based only on the Placido and elevation maps." |
| Mandatory core | Per-eye exclusion rulers (CORE-3) | Ruiz Hidalgo et al., 2017 (FT687) | Not reported | Center standards and workflows are named, but the full prespecified ruler set and per-eye measurements are not reported. | Evidence record ruiz-hidalgo-et-al-2017-ft687 (Zenodo archive, 02_claim_adjudication) | "Each clinical diagnosis was performed according to the standards and workflow of the center that performed it." |
| Mandatory core | Per-eye exclusion rulers (CORE-3) | Chan et al., 2015 (FT688) | Met | Target-eye normality required negative KCI/KSI, no steep keratometry (>47 D), no maximum posterior elevation >40 µm and no asymmetric anterior keratographic pattern. | Evidence record chan-et-al-2015-ft688 (Zenodo archive, 02_claim_adjudication) | "including a negative Tomey keratoconus screening report [using the Klyce/Maeda keratoconus index (KCI) and the Smolek/Klyce index (KSI)] … and without a steep keratometric curvature of >47 D, maximum posterior corneal elevation of >40 µm, and an asymmetric anterior keratographic pattern in the axial power map" |
| Mandatory core | Per-eye exclusion rulers (CORE-3) | Herber et al., 2022 (FT378) | Met | VAE-NTT certification reports KISA%, I-S, Kmax, and BAD-D thresholds for the target eye. | Evidence record herber-et-al-2022-ft378 (Zenodo archive, 02_claim_adjudication) | "KISA% of less than 60, I-S value of less than 1.45 D, Kmax of less than 47.00 D, and BAD-D of less than 1.6" |
| Claim extension | Orthogonal pathway (EXT-1) | Karimi et al., 2018 (FT126) | Shared | Corvis ST supplies the model inputs, and the Corvis also entered the Corvis/Pentacam-assisted identity pathway, so the detector lies inside the label measurement family. | Evidence record karimi-et-al-2018-ft126 (Zenodo archive, 02_claim_adjudication) | "the inputs of the model … are all the clinical data provided by Corvis-ST"; "identified via the Corvis and Pentacam by help of three ophthalmologists" |
| Claim extension | Orthogonal pathway (EXT-1) | Issarti et al., 2020 (FT512) | Shared | Model inputs are Pentacam elevation and thickness measurements, while target normality is also defined tomographically. | Evidence record issarti-et-al-2020-ft512 (Zenodo archive, 02_claim_adjudication) | "showing no … tomographic changes"; "Anterior and posterior corneal elevation data were exported as Pentacam CSV files" |
| Claim extension | Orthogonal pathway (EXT-1) | Wahba et al., 2018 (FT666) | Shared | I-S is calculated from the axial curvature display and IHD from elevation data; both are derived from the same Oculyzer/Pentacam scan. | Evidence record wahba-et-al-2018-ft666 (Zenodo archive, 02_claim_adjudication) | "Every eye was scanned … by a rotating Scheimpflug camera (Allegro Oculyzer II equivalent to Pentacam HR)"; "I-S value … in the axial (sagittal) curvature display"; "IHD … decentration of elevation data … calculated automatically by the device" |
| Claim extension | Orthogonal pathway (EXT-1) | Saad & Gatinel, 2012 (FT686) | Shared | Both the subjective labels and the score use Placido, elevation, and tomography maps from the target-eye measurement pathway. | Evidence record saad-gatinel-2012-ft686 (Zenodo archive, 02_claim_adjudication) | "classified … based only on the Placido and elevation maps"; "uses a combination of Placido and tomography corneal indices obtained with the Orbscan system" |
| Claim extension | Orthogonal pathway (EXT-1) | Ruiz Hidalgo et al., 2017 (FT687) | Not reported | The two centers used different tomographers for diagnosis, but the report does not provide one detector–reference pairing that supports a single orthogonal or shared classification for the combined analysis. | Evidence record ruiz-hidalgo-et-al-2017-ft687 (Zenodo archive, 02_claim_adjudication) | "the diagnoses were based on 2 different tomographers (Placido and Orbscan in Rothschild, and Pentacam at UZA)" |

| Module | Reporting item | Study | Table 3 status | Evidence statement | Source | Source excerpt(s) |
|---|---|---|---|---|---|---|
| Claim extension | Orthogonal pathway (EXT-1) | Chan et al., 2015 (FT688) | Shared | SCORE and target certification both use Orbscan/Tomey topographic and tomographic information. | Evidence record chan-et-al-2015-ft688 (Zenodo archive, 02_claim_adjudication) | "Orbscan IIz and Tomey keratoconus screening system"; "It combines 12 Placido and tomographic indices" |
| Claim extension | Orthogonal pathway (EXT-1) | Herber et al., 2022 (FT378) | Orthogonal | Pentacam supplies the topographic/tomographic exclusion rulers, whereas Corvis ST and ORA supply biomechanical detector measurements. | Evidence record herber-et-al-2022-ft378 (Zenodo archive, 02_claim_adjudication) | "topographically and tomographically normal … BAD-D of less than 1.6"; "Corneal tomography was measured using … Pentacam"; "The Corvis ST … biomechanically related … parameters were analyzed"; "The ORA measures the dynamic corneal behavior" |
| Claim extension | Target transport and refit (EXT-2) | Karimi et al., 2018 (FT126) | Not demonstrated | The trained ANN was tested on independent eyes, but the same healthy/suspect/keratoconus stress ranges defined the test labels; no frozen score was evaluated across target definitions. | Evidence record karimi-et-al-2018-ft126 (Zenodo archive, 02_claim_adjudication) | "Three different ranges were defined"; "This stress was then compared to the ranges that we provided for different conditions of the eye"; "we used independent data to test our model" |
| Claim extension | Target transport and refit (EXT-2) | Issarti et al., 2020 (FT512) | Not demonstrated | UZG data excluded from training supplied external-center validation of the same five-stage Logik output; no frozen score was evaluated across target definitions. | Evidence record issarti-et-al-2020-ft512 (Zenodo archive, 02_claim_adjudication) | "five groups were considered: normal, suspect keratoconus, early, mild, and moderate to advanced keratoconus"; "used to perform an additional, external validation" |
| Claim extension | Target transport and refit (EXT-2) | Wahba et al., 2018 (FT666) | Not reported | A validation sample evaluated the deduced IHD formula, but fixed-score target transport or an equivalent stratified gradient was not reported. | Evidence record wahba-et-al-2018-ft666 (Zenodo archive, 02_claim_adjudication) | "A validation sample of 78 eyes … was then collected prospectively"; "Regarding the validation sample, the AUROC … of the absolute difference between the actual and the deduced IHD were calculated"; "the deduced IHD from the abovementioned formula" |
| Claim extension | Target transport and refit (EXT-2) | Saad & Gatinel, 2012 (FT686) | Not demonstrated | The existing score was applied to consecutive eyes excluded from score creation and compared with the same suitable/at-risk map classification; no frozen score was evaluated across target definitions. | Evidence record saad-gatinel-2012-ft686 (Zenodo archive, 02_claim_adjudication) | "consecutive cases not included in the previous analysis to create the artificial intelligence system"; "classified … as either 'suitable' or 'at risk'"; "None of the eyes of the tested group … was included in the original group … used for the score creation" |
| Claim extension | Target transport and refit (EXT-2) | Ruiz Hidalgo et al., 2017 (FT687) | Not demonstrated | KA (Keratoconus Assistant) was applied to a cohort not used in development but evaluated against the same clinical group scheme; no frozen score was evaluated across target definitions. | Evidence record ruiz-hidalgo-et-al-2017-ft687 (Zenodo archive, 02_claim_adjudication) | "applying it to a cohort that was not involved in its development"; "These eyes were not used in the previous analysis that defined KA"; "classified into the same groups as before" |
| Claim extension | Target transport and refit (EXT-2) | Chan et al., 2015 (FT688) | Not demonstrated | Masked investigators applied the existing SCORE Analyzer in a new Asian cohort under the same FFKC-versus-control definition; no frozen score was evaluated across target definitions. | Evidence record chan-et-al-2015-ft688 (Zenodo archive, 02_claim_adjudication) | "independently tested them with the SCORE Analyzer through masked investigators"; "We classified our study eyes into 2 groups"; "All topographies in our study were tested with the SCORE Analyzer" |

| Module | Reporting item | Study | Table 3 status | Evidence statement | Source | Source excerpt(s) |
|---|---|---|---|---|---|---|
| Claim extension | Target transport and refit (EXT-2) | Herber et al., 2022 (FT378) | Met | CBI, a fixed biomechanical score, was evaluated against the same normal eyes across the ordered VAE-NT and VAE-NTT strata within one monocentric cohort (AUC 0.992 and 0.950; sensitivity and specificity reported at the fixed cut-off of 0.2), an equivalent stratified gradient; cross-cohort transport and refit were not reported. | Evidence record herber-et-al-2022-ft378 (Zenodo archive, 02_claim_adjudication) | "The current prospective, observational, and monocentric study"; "NE vs VAE-NT … 0.992 … ≥ 0.2"; "NE vs VAE-NTT … 0.950 … ≥ 0.2"; "The AUC of CBI was above 0.95, if normal eyes were compared either with all ectatic subgroups, mild/moderate keratoconus, VAE-NTT, or VAE-NT" |
| Claim extension | Demographic and site baseline (EXT-3) | Karimi et al., 2018 (FT126) | Not demonstrated | Age and gender were recorded but explicitly excluded as analysis factors; no matching or demographic-only baseline was reported. | Evidence record karimi-et-al-2018-ft126 (Zenodo archive, 02_claim_adjudication) | "Although age and gender of the participants have been recorded, they have not been considered as effective factors in the analyses." |
| Claim extension | Demographic and site baseline (EXT-3) | Issarti et al., 2020 (FT512) | Conditional | Two centers and an external-center check are reported, but demographic information is aggregate and no demographic-only baseline is given. | Evidence record issarti-et-al-2020-ft512 (Zenodo archive, 02_claim_adjudication) | "These Scheimpflug measurements were retrospectively collected from two centres"; "used to perform an additional, external validation … at an external center" |
| Claim extension | Demographic and site baseline (EXT-3) | Wahba et al., 2018 (FT666) | Not demonstrated | Cohort age and sex counts are reported without matching, stratification, or a demographic-only baseline. | Evidence record wahba-et-al-2018-ft666 (Zenodo archive, 02_claim_adjudication) | "The mean (±SD) age of the patients was 28.5 ± 7.5 years; 41 eyes of males and 87 of females" |
| Claim extension | Demographic and site baseline (EXT-3) | Saad & Gatinel, 2012 (FT686) | Conditional | Age is reported by subjective classification group, but no demographic-only baseline is reported. | Evidence record saad-gatinel-2012-ft686 (Zenodo archive, 02_claim_adjudication) | "Patient's demographics … in each subgroup … are listed in Table 1"; "Patient's characteristics in the population and in each subgroup" |
| Claim extension | Demographic and site baseline (EXT-3) | Ruiz Hidalgo et al., 2017 (FT687) | Conditional | Group demographics and center-specific clinical classifications are reported, but no demographic-only baseline is provided. | Evidence record ruiz-hidalgo-et-al-2017-ft687 (Zenodo archive, 02_claim_adjudication) | "The demographic details of the groups can be found in Table 1"; "the same subjects were diagnosed independently by … UZA"; "Description of the Study Groups According to the UZA and Rothschild Classifications" |
| Claim extension | Demographic and site baseline (EXT-3) | Chan et al., 2015 (FT688) | Not demonstrated | Age, race, and sex are tabulated, but sex and racial composition differ between groups and no demographic-only baseline is reported. | Evidence record chan-et-al-2015-ft688 (Zenodo archive, 02_claim_adjudication) | "There were more men in the FFKC group"; "a disproportionately high percentage of patients were Chinese (95.2% vs. 70.8% in the FFKC group)" |
| Claim extension | Demographic and site baseline (EXT-3) | Herber et al., 2022 (FT378) | Conditional | Demographics are tabulated and age is compared across subgroups, but no demographic-only baseline is reported. | Evidence record herber-et-al-2022-ft378 (Zenodo archive, 02_claim_adjudication) | "The demographic data of the study cohort are presented in Table A"; "participants with normal eyes had a similar age compared to patients with moderate keratoconus, VAE-NTT, and VAE-NT" |

#### B. Study limitations and constructive roles

| Study | Remaining limitation | Constructive role |
|---|---|---|
| Karimi et al., 2018 (FT126) | Shared mechanical-property values were applied across healthy and keratoconic models, which may limit physiological specificity. | Demonstrates a Corvis-based modeling approach across healthy, suspect, and keratoconic eyes. |
| Issarti et al., 2020 (FT512) | Modest overlap remained between healthy and suspect keratoconus cases. | Adds suspect-keratoconus detection and an objective severity score to five-stratum classification. |

| Study | Remaining limitation | Constructive role |
|---|---|---|
| Wahba et al., 2018 (FT666) | The analysis selected right eyes except for FFKC because these cases were scarce. | Shows strong correlation between I-S and IHD but does not establish strict-target diagnostic performance. |
| Saad & Gatinel, 2012 (FT686) | The scoring method is tied to Orbscan topographers. | Shows reasonable concordance between objective and subjective classifications for early subclinical keratoconus detection. |
| Ruiz Hidalgo et al., 2017 (FT687) | Equipment and forme fruste definitions differed between centers. | Supports cross-sectional identification of forme fruste keratoconus framed as at risk of ectasia after refractive surgery; it does not establish prediction of post-LASIK ectasia. |
| Chan et al., 2015 (FT688) | No available system detected all FFKC cases or predicted post-LASIK ectasia unequivocally. | Supports cross-sectional identification of FFKC framed as at risk of post-LASIK keratectasia; it does not establish prediction of post-LASIK ectasia. |
| Herber et al., 2022 (FT378) | Generalizability is limited by a White study population and small keratoconus sample. | Architecture positive example: tomographic exclusion ruler plus an orthogonal biomechanical detector. |

Source excerpts are quoted from the published reports; ellipses mark gaps between noncontiguous passages. Part B gives the complete limitation and constructive-role statements.

#### Zenodo archive contents

https://doi.org/10.5281/zenodo.22304145

The archive is organized in eleven numbered directories: 01_corpus_roster/; 02_claim_adjudication/; 03_index_provenance/; 04_partial_identification/; 05_target_substitution_experiment/; 06_cross_device_and_demographic/; 07_verification_subsample/; 08_search_strategy/; 09_registration_and_deviations/; 10_prisma/; 11_supplementary_tables/. Each directory carries a README describing its files.
